# Visual Outcomes After Radiotherapy and Radiosurgery for Optic Pathway Hypothalamic Glioma: A Systematic Review and Meta-analysis

**DOI:** 10.64898/2026.07.25.26358933

**Authors:** Farzan Fahim, Amirmahdi Mojtahedzadeh, Farzin Mohammad Moradi, Zahra Zali, Zahra Darvishi, Vesal Abbasian, Mobina Puraminaie, Negin Aflaki, Shima Khalili, Parisa Fooladi, Seyyed Mohammad Hosseini Marvast, Parshana Farsandaj, Mohammadreza AmaniTehrani, Fatemeh Khazaei, Nastaran Sadat Mahdavi, Saeed Safari, Alireza Zali

## Abstract

**Background:** Optic pathway hypothalamic glioma (OPHG) can cause irreversible visual impairment. Radiotherapy and radiosurgery are used for progressive, recurrent, or treatment-refractory disease, but their effects on visual outcomes remain incompletely defined.

**Objective:** To synthesize visual outcomes following radiotherapy or radiosurgery for OPHG, with visual preservation as the primary outcome.

**Methods:** This prospectively registered systematic review and meta-analysis (PROSPERO CRD420261440136) followed the PRISMA 2020 statement. PubMed, Scopus, Web of Science, Embase, the Cochrane Library, Google Scholar, and ClinicalTrials.gov were searched from inception to 1 June 2026. Eligible studies included patients with OPHG or related optic pathway/hypothalamic gliomas treated with radiotherapy or radiosurgery and reporting extractable visual outcomes. Visual preservation was defined as stable or improved vision. Random-effects meta-analyses of logit-transformed proportions were performed.

**Results:** Forty-nine studies were included in the systematic review, of which 19 contributed 494 visual-outcome observations to the meta-analysis. The pooled visual preservation rate was 75.6% (95% CI 65.1–83.7%; I^2^ = 71.6%). Pooled rates of visual improvement, stability, and worsening were 24.7% (95% CI 19.5–30.8%), 46.7% (95% CI 37.0–56.6%), and 20.6% (95% CI 12.7–31.6%), respectively. Preservation was higher in pure radiotherapy/radiosurgery cohorts than in mixed-treatment cohorts (85% vs 66%; subgroup p = 0.0104), although differences in cohort composition and treatment attribution may have influenced this finding.

**Conclusion:** Radiotherapy and radiosurgery were associated with preservation of vision in approximately three-quarters of evaluable outcomes, predominantly through visual stability rather than recovery. Standardized visual-outcome definitions are needed to improve future comparisons and treatment evaluation.

## Introduction

Optic pathway hypothalamic glioma (OPHG) is a rare and clinically heterogeneous tumor involving the optic nerve, optic chiasm, optic tract, hypothalamus, or contiguous visual pathway structures. It occurs predominantly in children and may arise sporadically or in association with neurofibromatosis type 1 (NF1). Although some tumors follow an indolent course, others progress within the chiasmatic-hypothalamic region and may cause substantial visual, endocrine, and neurodevelopmental morbidity. Visual impairment is often the principal clinical concern and may manifest as reduced visual acuity, visual field loss, optic atrophy, strabismus, nystagmus, proptosis, or blindness. Hypothalamic involvement may additionally contribute to endocrine dysfunction, obesity, and long-term developmental consequences [1–11] .

Management is individualized according to age, NF1 status, tumor location, visual trajectory, radiological progression, and prior treatment. Observation may be appropriate for clinically and radiologically stable disease, whereas chemotherapy and, increasingly, targeted systemic therapies are used for progressive pediatric low-grade glioma. Surgery is generally limited to biopsy, decompression, cyst management, or selected exophytic components because of the risk of injury to the optic pathways and hypothalamus. Radiotherapy, including conventional photon radiotherapy, conformal radiotherapy, intensity-modulated radiotherapy, proton therapy, fractionated stereotactic radiotherapy, and stereotactic radiosurgery/Gamma Knife, remains a treatment option for selected patients with progressive, recurrent, or treatment-refractory disease when durable tumor control and preservation of remaining visual function are considered important therapeutic goals [12–22].

Despite several decades of clinical experience, the effect of radiotherapy and radiosurgery on visual outcomes in OPHG remains uncertain. The available literature is heterogeneous with respect to radiation technique, treatment timing, patient selection, baseline visual status, follow-up duration, outcome definitions, and whether radiotherapy-specific results can be distinguished from those of mixed-treatment cohorts [11, 23–33] . Moreover, prior reviews have generally emphasized overall management, radiological control, survival, or broad clinical outcomes, rather than quantitatively distinguishing visual preservation, improvement, stability, and worsening after radiation-based treatment. This distinction is clinically relevant because maintenance of existing visual function may represent a meaningful benefit even in the absence of visual recovery.

Accordingly, this systematic review and meta-analysis aimed to quantify visual outcomes after radiotherapy and radiosurgery for OPHG. The primary outcome was visual preservation, defined as stable or improved vision, while secondary outcomes included visual improvement, visual stability, and visual worsening.

## Methods

### Protocol and reporting

This systematic review and meta-analysis was conducted according to a predefined protocol registered in PROSPERO (CRD420261440136) and was reported in accordance with the PRISMA 2020 statement. The protocol specified the review question, eligibility criteria, information sources, search strategy, screening procedures, data extraction, risk-of-bias assessment, quantitative synthesis, subgroup analyses, and sensitivity analyses. The completed PRISMA 2020 checklist is provided as Supplementary File 1, the registered protocol as Supplementary File 2, and the PRISMA flow diagram as Figure 1.

**Figure 1.**
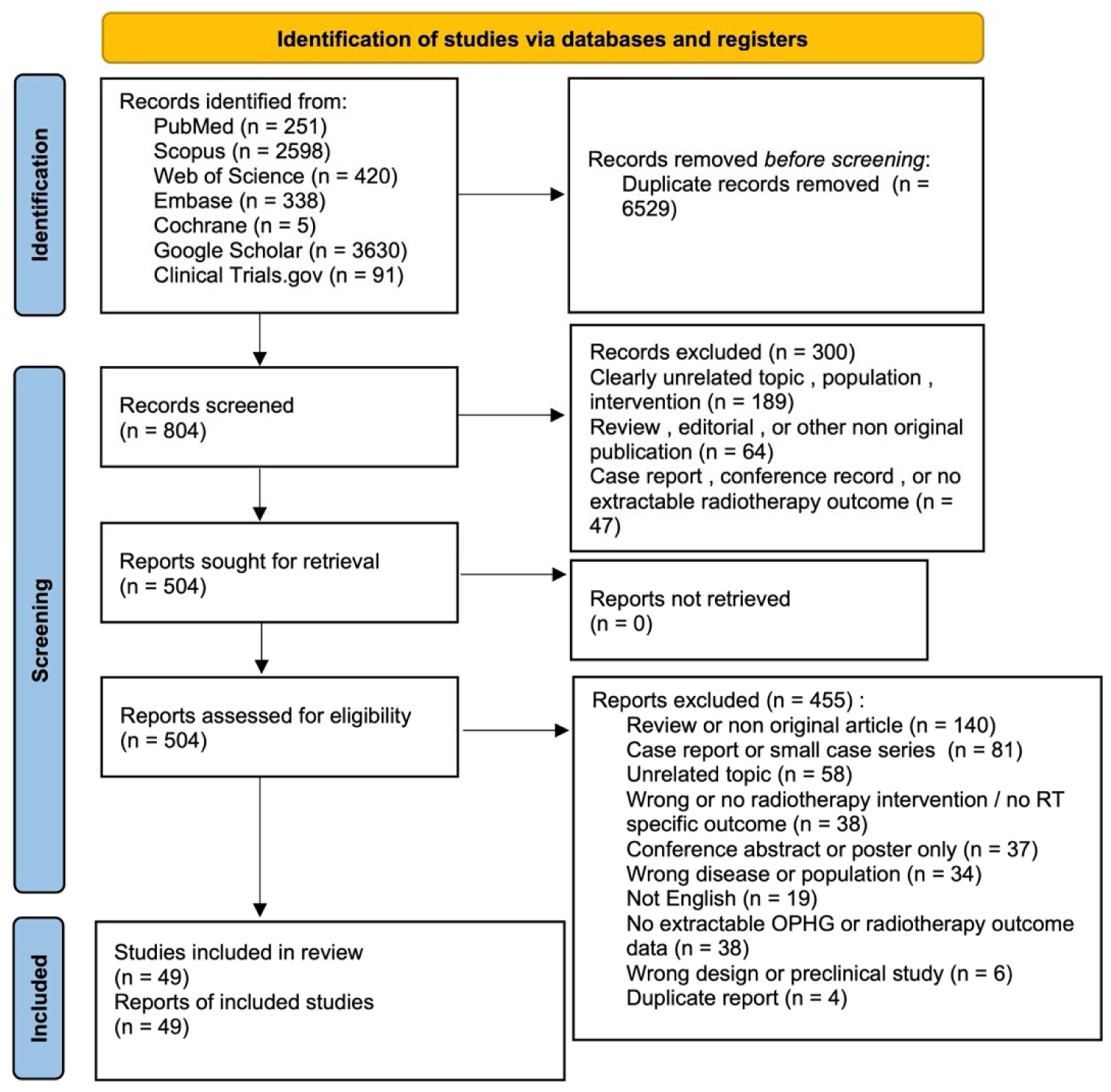
PRISMA 2020 flow diagram. PRISMA 2020 flow diagram illustrating study identification, screening, eligibility assessment, and study inclusion for the systematic review and meta-analysis.

### Search strategy and information sources

A systematic search was performed in PubMed/MEDLINE, Scopus, Web of Science, Embase, and the Cochrane Library from database inception to 1 June 2026, without restriction by publication year. Supplementary searches were conducted in Google Scholar and ClinicalTrials.gov to identify additional published or registered studies, particularly reports containing visual outcome data.

The search strategies combined controlled vocabulary and free-text terms related to optic pathway, optic nerve, chiasmatic, and hypothalamic gliomas with terms related to radiotherapy, radiosurgery, visual function, and treatment outcomes. Disease-related terms included “optic pathway glioma,” “optic nerve glioma,” “hypothalamic glioma,” “chiasmatic glioma,” “chiasmal glioma,” “optic pathway hypothalamic glioma,” “optic pathway/hypothalamic glioma,” and “OPHG.” Intervention-related terms included “radiotherapy,” “radiation therapy,” “irradiation,” “external beam radiotherapy,” “proton therapy,” “stereotactic radiosurgery,” “radiosurgery,” “Gamma Knife,” “IMRT,” and “fractionated radiotherapy.” Supplementary searches also incorporated visual-outcome terms, including “visual acuity,” “visual outcome,” “visual function,” “visual field,” “vision preservation,” “ophthalmologic,” and “blindness.”

Search syntax was adapted to each information source, including Medical Subject Headings in PubMed/MEDLINE and Emtree terms in Embase. Complete strategies for all databases, Google Scholar, and ClinicalTrials.gov are provided in Supplementary File 3. The PubMed/MEDLINE search syntax was:

((“Optic Nerve Glioma”[Mesh]) OR (“optic pathway glioma*”[Title/Abstract]) OR (“optic nerve glioma*”[Title/Abstract]) OR (“hypothalamic glioma*”[Title/Abstract]) OR (“chiasmatic glioma*”[Title/Abstract]) OR (OPHG[Title/Abstract])) AND ((“Radiotherapy”[Mesh]) OR (“radiation therap*”[Title/Abstract]) OR (radiotherap*[Title/Abstract]) OR (“proton therap*”[Title/Abstract]) OR (“stereotactic radiosurgery”[Title/Abstract]) OR (IMRT[Title/Abstract]))

### Eligibility criteria

Eligibility criteria were defined according to the PICOS framework.

#### Population

Eligible populations included patients of any age with optic pathway hypothalamic glioma or related gliomas involving the optic nerve, optic chiasm, optic tract, optic pathway, visual pathway, chiasmatic-hypothalamic region, or hypothalamic region. Studies describing optic pathway glioma, optic nerve glioma, optic chiasm glioma, optochiasmatic glioma, visual pathway glioma, optic pathway/hypothalamic glioma, hypothalamic glioma, or hypothalamic low-grade glioma were eligible. Pediatric and adult patients and both NF1-associated and sporadic tumors were considered eligible.

#### Intervention

Eligible interventions included conventional external-beam radiotherapy, historical photon radiotherapy, three-dimensional conformal radiotherapy, intensity-modulated radiotherapy, fractionated stereotactic radiotherapy, stereotactic radiosurgery, Gamma Knife radiosurgery, CyberKnife, proton therapy, and other radiation-based approaches. Studies were eligible for the systematic review when they reported original data concerning radiotherapy or radiosurgery in patients with OPHG. Eligibility for the quantitative visual synthesis additionally required extractable visual outcomes after RT/SRS or separately extractable outcomes for a clearly identifiable RT/SRS-treated subgroup.

#### Comparator

No comparator was required because the principal quantitative analyses were single-arm meta-analyses of proportions. Comparative studies were eligible when data for the RT/SRS-treated group could be extracted separately.

#### Outcomes

The primary outcome was visual preservation, defined as stable or improved vision after radiotherapy or radiosurgery. Secondary outcomes were visual improvement, visual stability, and visual worsening. Eligible visual assessments included visual acuity, visual fields, ophthalmological examination, blindness status, and author-defined categorical visual outcomes. Quantitative synthesis required an extractable number of patients, eyes, or visual-outcome observations categorized as improved, stable, or worsened, together with a corresponding denominator.

#### Study design

Eligible designs included prospective and retrospective cohort studies, longitudinal and multicenter observational cohorts, mixed retrospective/prospective cohorts, prospective or non-randomized interventional studies, single-arm observational studies, and retrospective case series containing original patient-level or aggregate outcome data. Conference abstracts or limited reports were eligible when they provided sufficient information to determine eligibility and extract relevant outcomes. When a publication combined an original cohort with a literature review, only the original cohort was considered for extraction and synthesis.

Studies were excluded when they did not satisfy the PICOS criteria; did not include OPHG or a related optic pathway/hypothalamic glioma population; did not involve radiotherapy or radiosurgery; did not provide original patient data or separable RT/SRS-specific outcomes; or involved non-glioma optic pathway tumors. Preclinical, laboratory, and animal studies were excluded. Reviews, meta-analyses, editorials, commentaries, expert opinions, protocols, letters without original patient data, and duplicate publications were also excluded. Case reports and case series containing fewer than 10 relevant patients without sufficiently extractable outcome data were excluded.

Eligibility for the systematic review and for the quantitative visual synthesis was assessed separately. Studies satisfying the broader clinical criteria but reporting visual outcomes only qualitatively, lacking a clear denominator, using non-separable patient-level and eye-level units, or failing to provide RT/SRS-specific visual results could remain eligible for narrative synthesis but were not included in the quantitative meta-analysis.

### Study selection and screening

All records retrieved from the searches were imported into EndNote 2025, and duplicate records were removed before screening. After deduplication, records were transferred to standardized Excel screening forms designed by the senior author (FF). The forms captured study identification, first author, publication year, title, relevance of the population, intervention and outcomes, study design, screening decision, exclusion reason, and reviewer comments.

Title and abstract screening was conducted by two reviewers (VA and MPA) according to the predefined eligibility criteria. The records were divided between the reviewers, while uncertain or potentially eligible records were retained for full-text assessment. Questions or disagreements were resolved through discussion and, when necessary, adjudication by AMM. The complete title and abstract exclusion sheet is provided as Supplementary File 4.

Potentially eligible reports subsequently underwent independent full-text assessment by two reviewers (SK and MH) using standardized Excel forms designed by FF. Each report was classified as included, excluded, or requiring discussion, and a predefined reason was recorded for each full-text exclusion. Disagreements were resolved by consensus and, when required, adjudication by AMM. The inclusion file and PICOS eligibility framework are provided as Supplementary File 5, and the full-text exclusion sheet is provided as Supplementary File 6.

The overall study-selection process was documented in a PRISMA 2020 flow diagram presented as Figure 1.

### Data extraction

Data extraction was performed using a standardized extraction sheet designed by FF. Two trained reviewers independently extracted the available data, and disagreements or uncertainties were resolved through discussion with AMM.

Extracted variables included study identification, bibliographic information, country, participating center or centers, study design, study period, inclusion and exclusion criteria, total study population, eligible OPHG population, RT/SRS subgroup size, age, sex, NF1 status, histology, tumor location and laterality, baseline visual manifestations, endocrine and neurological features, previous and concomitant treatments, radiation modality, treated target, radiation dose, fractionation, timing and indication, follow-up duration, radiological outcomes, visual outcomes, endocrine outcomes, progression, survival, adverse events, and eligibility for quantitative synthesis.

For visual analyses, the extracted data included the visual outcome denominator and the number of patients, eyes, or evaluable visual assessments classified as improved, stable, worsened, or preserved. A **visual-outcome observation** represented the unit used by the source study to report an evaluable post-treatment visual result. Depending on the original report, this could represent an individual patient, an individual eye, or another explicitly defined evaluable visual unit. Patient-level outcomes were prioritized whenever available. Eye-level or other observation-level data were used only when patient-level data could not be extracted separately, and the reported unit was retained to avoid treating these observations as equivalent to unique patients.

Visual preservation was calculated as the sum of improved and stable outcomes when these categories were reported separately. When several visual measures were available, an ophthalmologist-assessed or author-defined global visual outcome was prioritized for the principal analysis. Visual acuity- and visual field-specific outcomes were extracted separately when reported. Quantitative eligibility and the reason a study could not contribute to pooled visual outcomes were also recorded. The complete extraction dataset is provided as Supplementary File 7.

### Risk-of-bias assessment

Risk of bias was evaluated using the Joanna Briggs Institute critical appraisal tool appropriate to each study design. Cohort studies were assessed using the JBI cohort checklist, non-randomized interventional studies using the JBI quasi-experimental checklist, and case series or single-arm observational studies using the JBI case series checklist.

Two reviewers (ZZ and ZD) independently performed the critical appraisal. Each applicable item was judged as “yes,” “no,” “unclear,” or “not applicable.” An item was marked not applicable only when the criterion did not logically apply to the study design, comparator structure, or outcome framework. Not-applicable items were excluded from the scoring denominator and were not treated as unclear or negative judgments.

For each study, the proportion of fulfilled applicable criteria was calculated by dividing the number of “yes” judgments by the number of applicable items. Studies were then categorized as having low, moderate, or high risk of bias according to the predefined scoring framework. Studies fulfilling ≥75% of applicable items were categorized as low risk of bias, those fulfilling 50%–74% as moderate risk, and those fulfilling <50% as high risk. Disagreements were resolved through discussion and, when necessary, adjudication by AMM. The item-level traffic-light plot is presented as Figure 5, and the complete assessment is provided as Supplementary File 8.

### Data synthesis and quantitative analysis

The principal quantitative synthesis consisted of single-arm meta-analyses of proportions. The primary pooled outcome was visual preservation, defined as stable or improved vision after radiotherapy or radiosurgery. Secondary pooled outcomes were visual improvement, visual stability, and visual worsening. For each outcome, the event count and corresponding visual-outcome denominator were extracted from each eligible study.

Analyses were performed in R version 4.6.0 using the meta, metafor, readxl, dplyr, stringr, writexl, and ggplot2 packages. Random-effects models were selected a priori because clinical and methodological heterogeneity was anticipated across patient populations, tumor characteristics, NF1 status, radiation modalities, treatment timing, previous or concomitant therapies, follow-up durations, visual assessment methods, and outcome definitions.

Proportions were modeled on the logit scale using generalized linear mixed models and subsequently back-transformed for presentation. Statistical heterogeneity was evaluated using Cochran’s Q, I^2^, and tau^2^. Prediction intervals were reported when estimable to describe the expected range of effects across comparable future settings. Results were expressed as pooled proportions with 95% confidence intervals.

Forest plots were generated for visual preservation, visual improvement, visual stability, and visual worsening. Supplementary forest plots, funnel-plot analyses, and leave-one-out analyses are provided in Supplementary File 9.

### Subgroup, sensitivity, and small-study-effect analyses

Predefined subgroup analyses examined visual outcomes according to radiation technique, radiotherapy timing, study purity, and risk-of-bias category. Radiation-technique categories comprised historical conventional photon radiotherapy, three-dimensional conformal photon radiotherapy, fractionated stereotactic radiotherapy, stereotactic radiosurgery/Gamma Knife, and mixed or insufficiently specified radiation approaches. SRS/Gamma Knife cohorts were additionally examined as a separate exploratory subgroup because of differences in dose delivery, target precision, fractionation, and patient selection.

Radiotherapy-timing categories included salvage radiotherapy, primary or upfront radiotherapy, adjuvant radiotherapy, radiotherapy after surgery, and mixed or unclear timing. Study-purity analyses compared cohorts composed exclusively of RT/SRS-treated patients with mixed-treatment cohorts and abstract-only or limited-report cohorts. Risk-of-bias analyses compared studies categorized as low, moderate, or high risk of bias.

Sensitivity analyses included restriction to full-text reports, exclusion of studies at high risk of bias, restriction to pure RT/SRS cohorts, and leave-one-out analysis. Leave-one-out analyses repeated the primary model after omitting one study at a time to assess the influence of individual studies on the pooled estimate.

For outcomes with a sufficient number of contributing studies, small-study effects were explored using funnel plots. Statistical tests for funnel-plot asymmetry and trim-and-fill analyses were treated as exploratory because of the observational evidence base, substantial clinical heterogeneity, and limited power of asymmetry tests in rare-disease meta-analyses. Detailed subgroup and sensitivity analyses are provided in Supplementary File 10.

## Results

### Study selection

The database, search-engine, and trial-registry searches identified 7,333 records: 251 from PubMed/MEDLINE, 2,598 from Scopus, 420 from Web of Science, 338 from Embase, 5 from the Cochrane Library, 3,630 from Google Scholar, and 91 from ClinicalTrials.gov. After removal of 6,529 duplicate records, 804 records underwent title and abstract screening. Of these, 300 were excluded because they addressed an unrelated topic, population, or intervention (n = 189); were reviews, editorials, or other non-original publications (n = 64); or were case reports, conference records, or reports without extractable radiotherapy-related outcomes (n = 47).

All 504 reports sought for retrieval were obtained and assessed in full text. Of these, 455 were excluded: review or other non-original publication (n = 140), case report or small case series (n = 81), unrelated topic (n = 58), wrong or absent radiotherapy intervention or absence of an RT-specific outcome (n = 38), conference abstract or poster without sufficient extractable data (n = 37), wrong disease or population (n = 34), non-English publication (n = 19), absence of extractable OPHG or radiotherapy outcome data (n = 38), ineligible design or preclinical study (n = 6), and duplicate report (n = 4).

Forty-nine studies met the eligibility criteria for the systematic review. Of these, 19 provided sufficiently complete and compatible categorical visual outcome data with corresponding denominators and were included in the quantitative synthesis, contributing 494 evaluable visual-outcome observations. The remaining 30 studies were retained for narrative synthesis because they provided relevant information on treatment, tumor control, survival, endocrine morbidity, surgery, imaging, toxicity, or descriptive visual outcomes but lacked a complete extractable visual denominator, separable RT/SRS-specific outcomes, or sufficiently compatible categorical visual data for pooling.

The study-selection process is presented in the PRISMA 2020 flow diagram (Figure 1). The title and abstract exclusion sheet is provided in Supplementary File 4, the inclusion file and PICOS eligibility framework in Supplementary File 5, and the full-text exclusion sheet in Supplementary File 6.

### Characteristics of the included studies

The 49 included studies were published between 1988 and 2025 and represented a predominantly observational evidence base. Forty studies were retrospective, six were prospective or protocol-based, and three used mixed or hybrid designs, including combined retrospective/prospective follow-up or retrospective analyses of non-randomized treatment studies. No randomized controlled trials or case-control studies were identified.

The studies were conducted across several geographical regions. The United States contributed the largest number of reports, followed by China, the United Kingdom, Canada, Italy, France, Turkey, and the Netherlands. Both single-center and multicenter studies were represented, although most were conducted at individual tertiary pediatric oncology, neurosurgical, or radiation oncology centers. Study characteristics are summarized in Table 1.

**Table 1.**
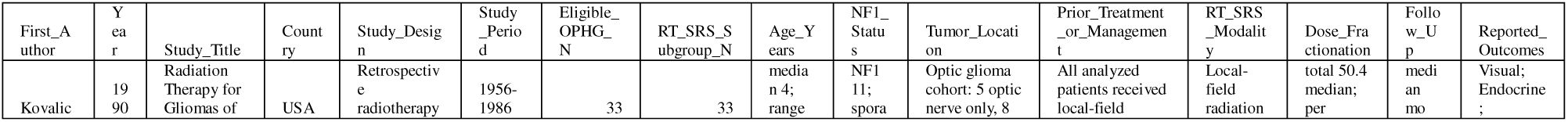
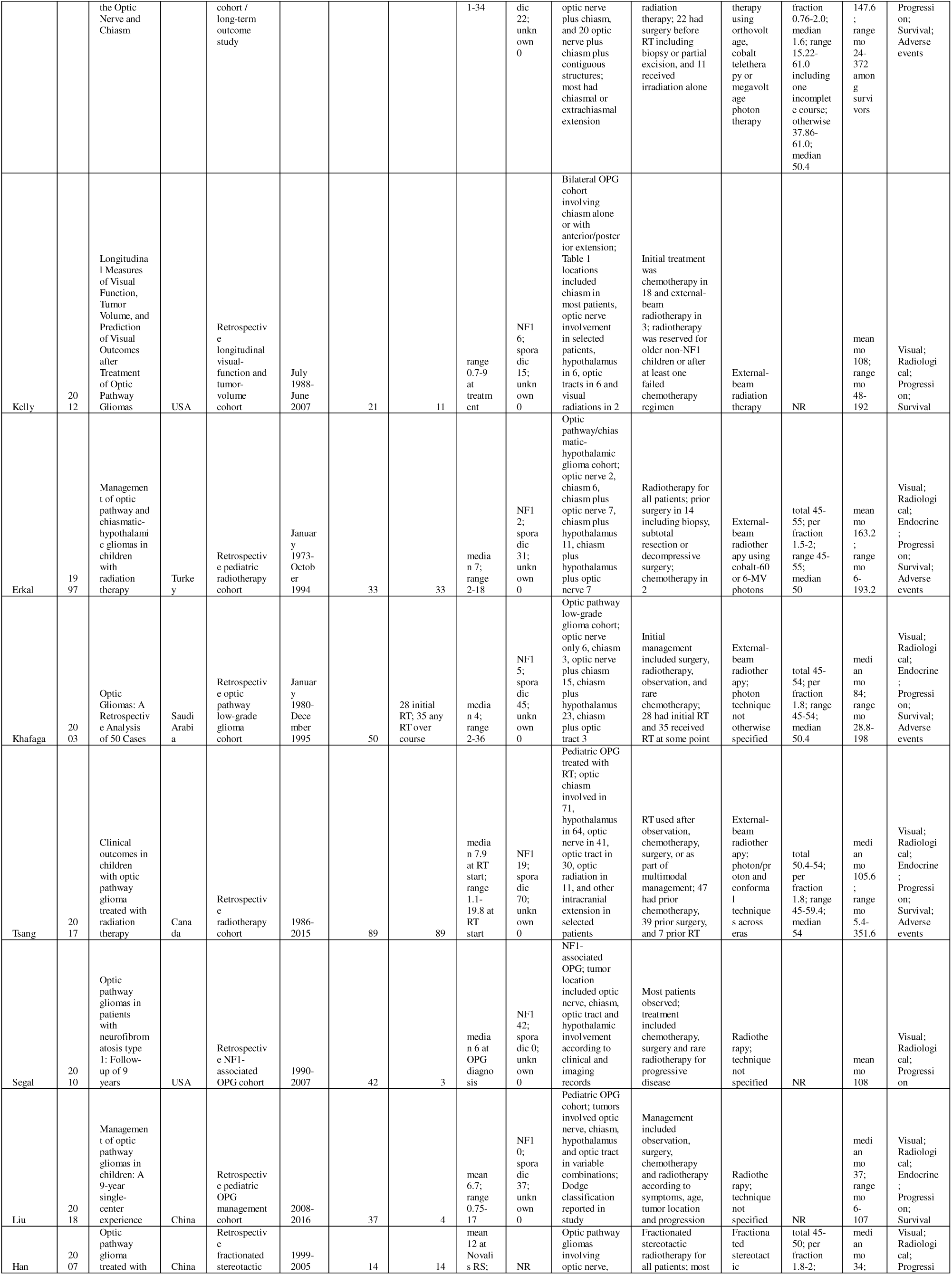
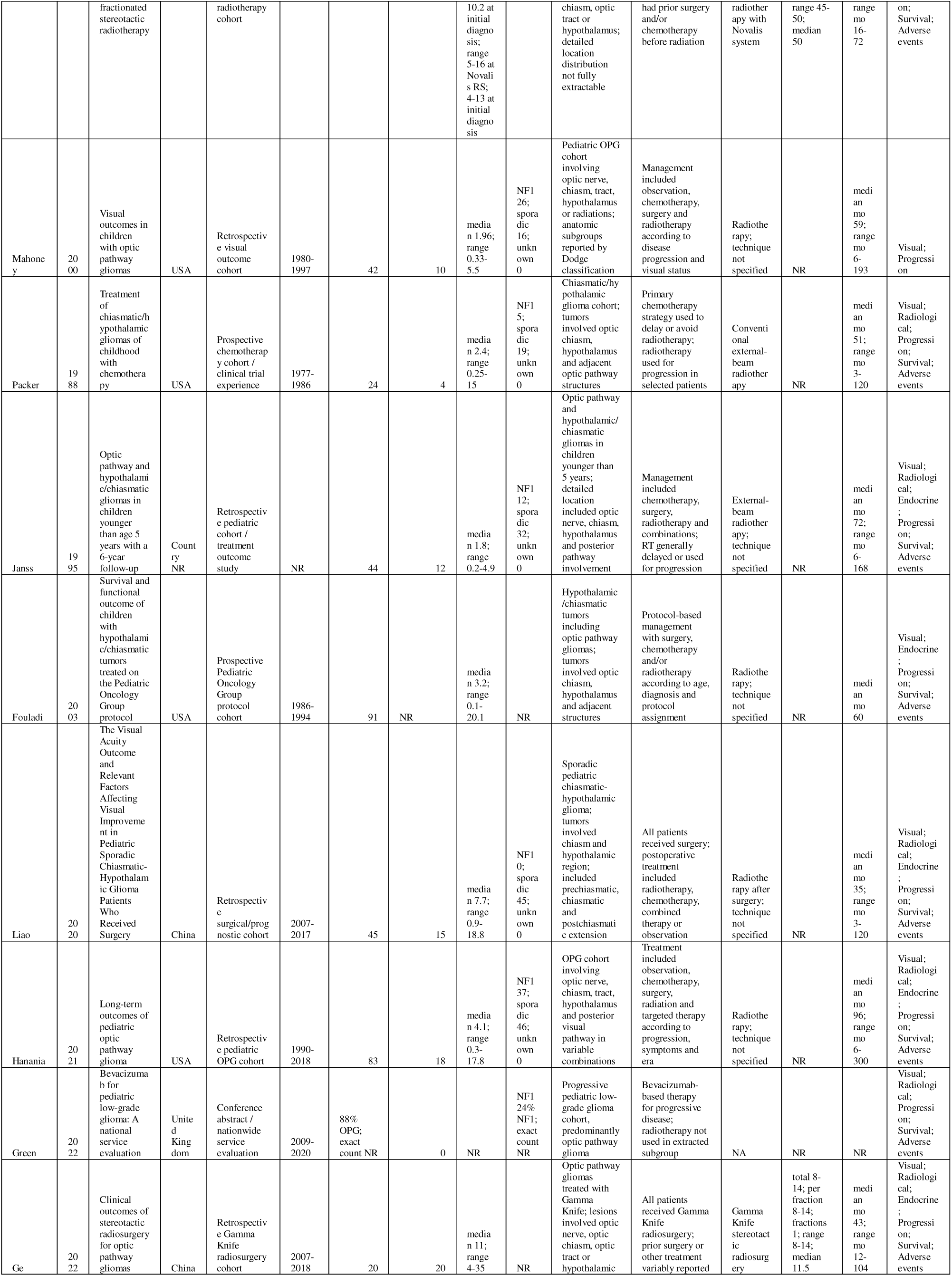
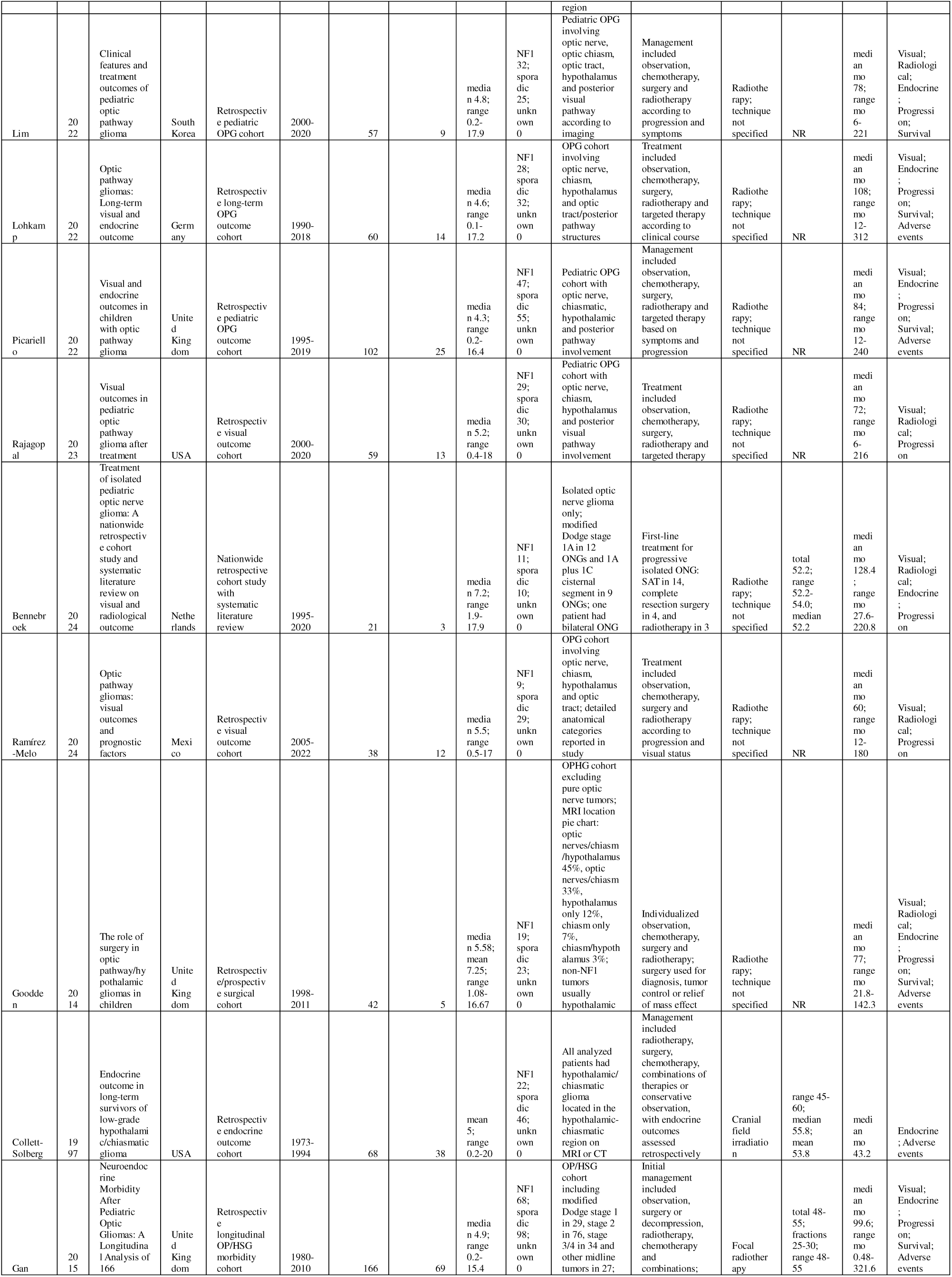
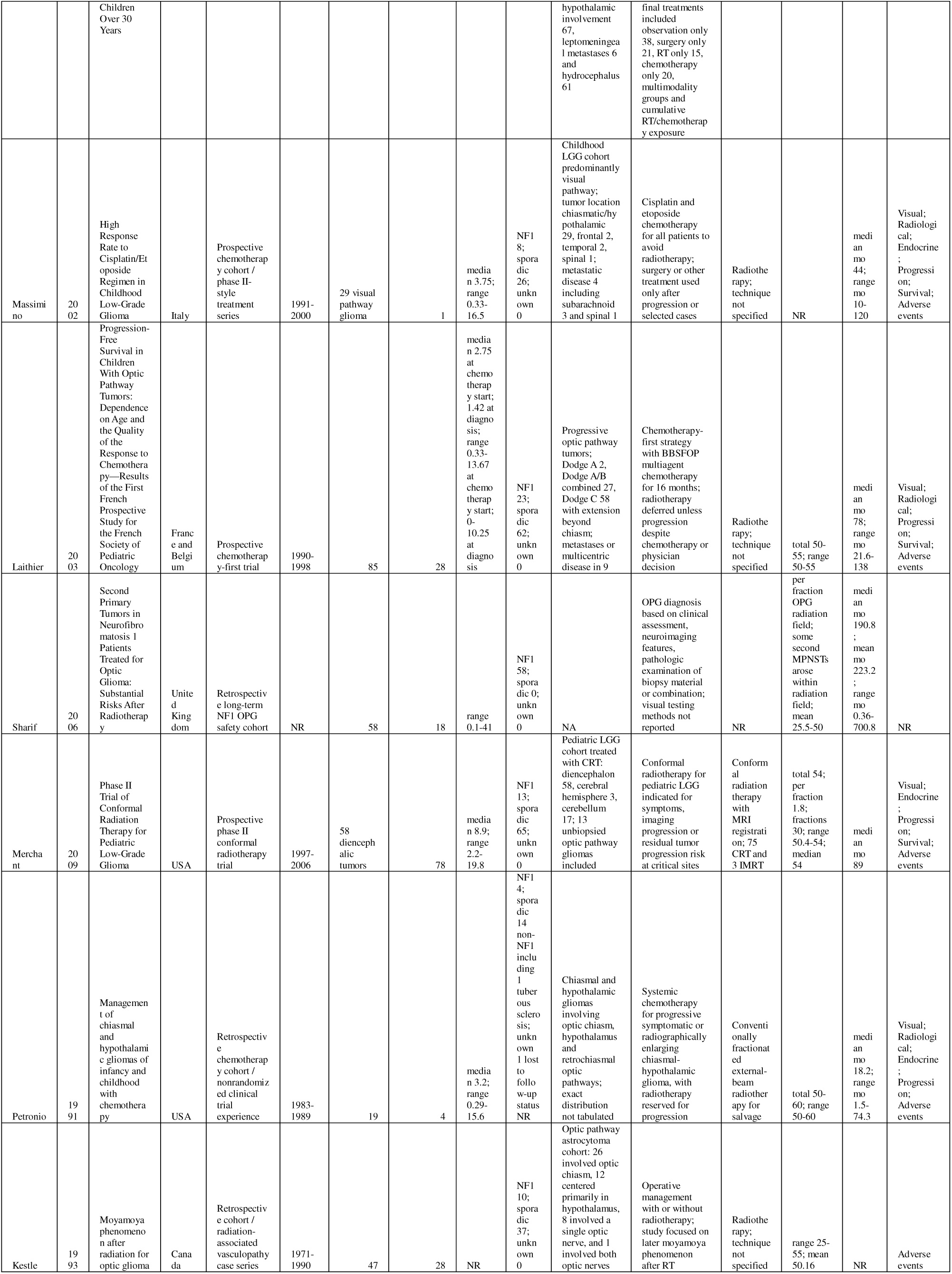
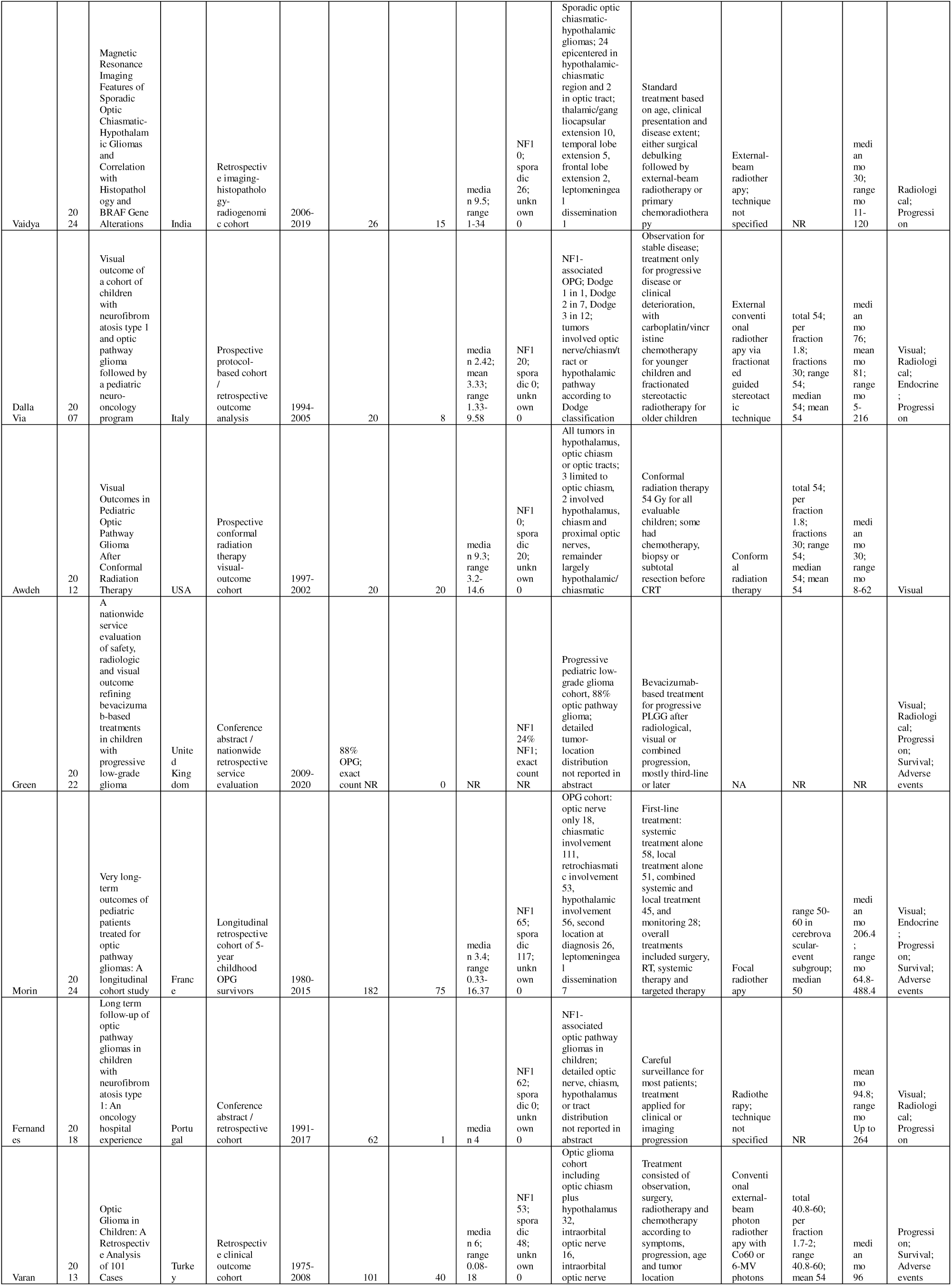
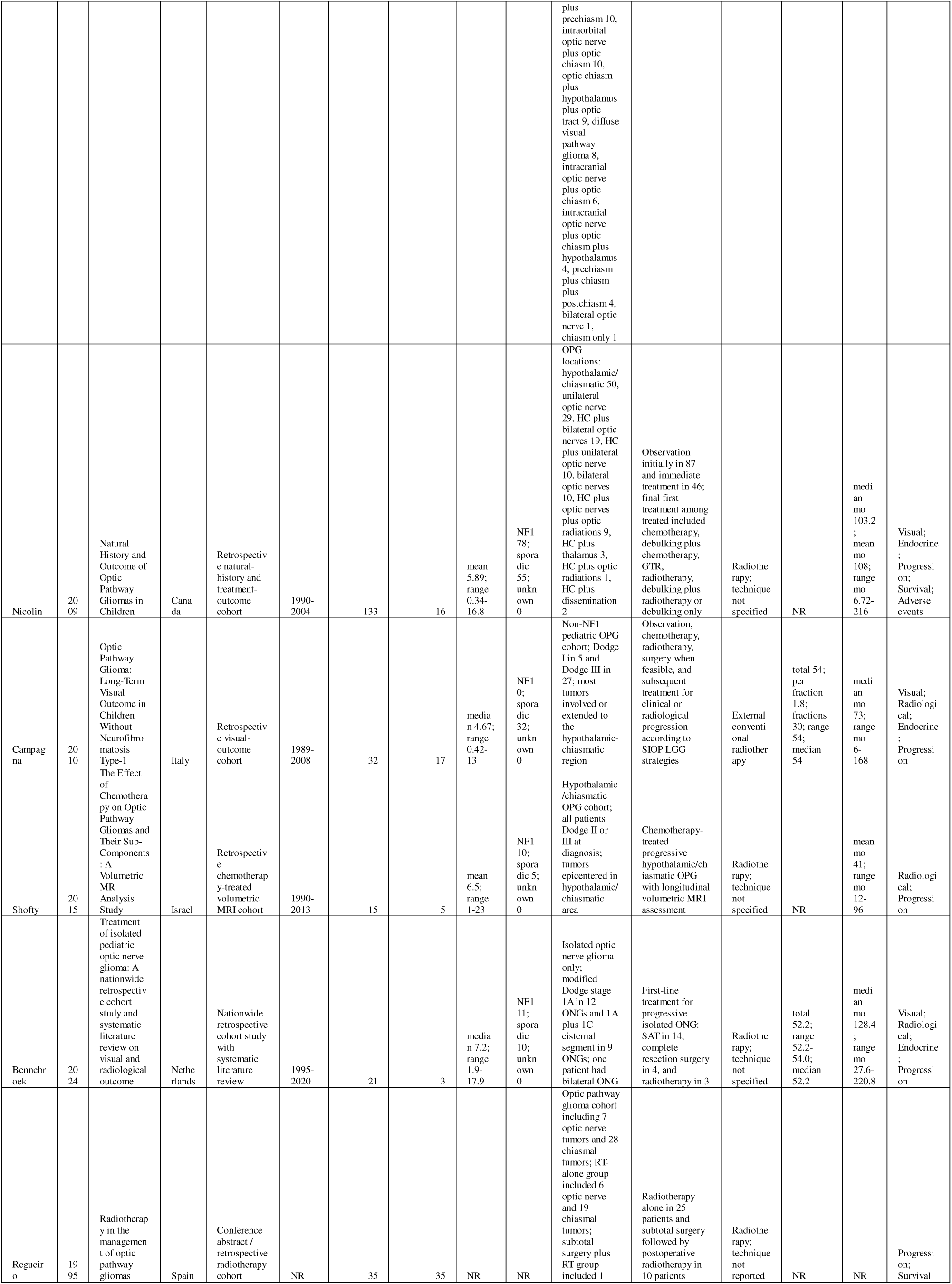
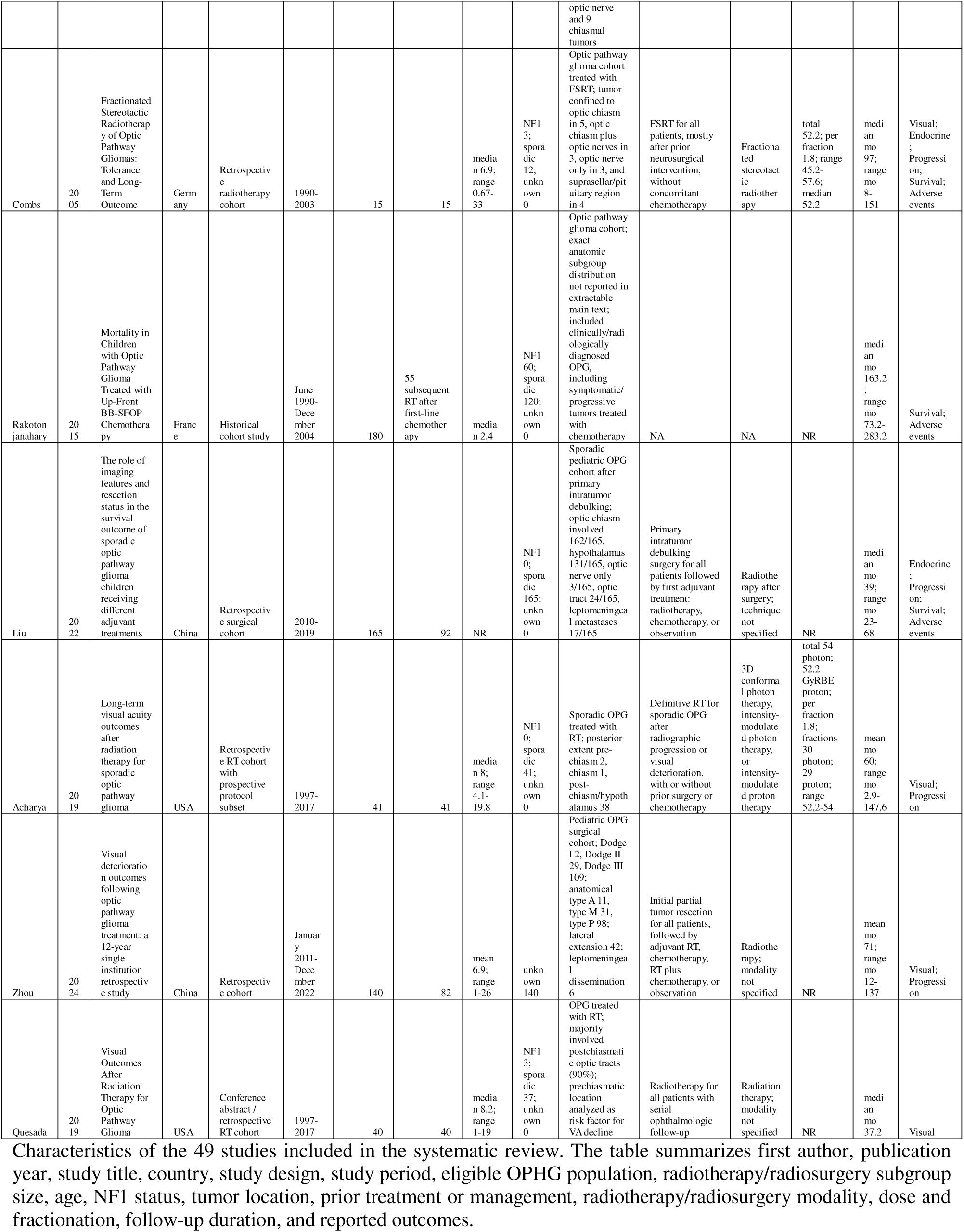
Study characteristics of included studies.

Across the included reports, data were abstracted for 3,125 patients with OPHG or related optic pathway/hypothalamic gliomas. Where a radiation-treated subgroup could be identified numerically, 1,307 patients had received radiotherapy or radiosurgery. These totals represent reported study populations rather than necessarily unique individuals, because some publications described broader institutional cohorts, treatment-specific subgroups, or potentially overlapping historical populations.

### Patient and tumor characteristics

The evidence base was predominantly pediatric. Thirty-one studies reported a numeric median age; the median of these study-level medians was 4.7 years, with individual study medians ranging from 1.6 to 16 years. Several reports included adolescents or adults, but exclusively adult cohorts were uncommon.

Numeric sex data were available in 41 studies and were nearly balanced, comprising 1,317 male and 1,330 female patients. NF1-associated and sporadic tumors were both well represented. Numeric NF1 counts were reported in 44 studies, comprising 1,018 NF1-associated cases, while 43 studies reported 1,802 sporadic cases. These values were not always derived from identical denominators, and several mixed cohorts did not provide treatment- or outcome-specific stratification by NF1 status.

Tumor anatomy was heterogeneous. Chiasmatic involvement was reported in 37 studies and hypothalamic involvement in 31; nearly all studies reporting these variables included at least some patients with chiasmatic or hypothalamic disease. Optic nerve, optic tract, bilateral, and more broadly defined visual pathway involvement were also represented. However, anatomical classification systems, laterality, histological confirmation, and the extent of hypothalamic involvement were not reported uniformly.

Baseline visual morbidity ranged from mild acuity or field deficits to severe bilateral impairment or blindness. Reported manifestations included reduced visual acuity, visual field loss, optic atrophy, strabismus, nystagmus, and proptosis. The method and completeness of baseline ophthalmological assessment varied substantially across studies.

Follow-up was reported using medians, means, or ranges. Thirty-four studies provided a numeric median follow-up; the median of these study-level medians was 79.8 months, with study medians ranging from 18.2 to 206.4 months. Several historical cohorts included follow-up extending beyond 20 years. Patient, tumor, and follow-up characteristics are summarized in Table 2.

**Table 2.**
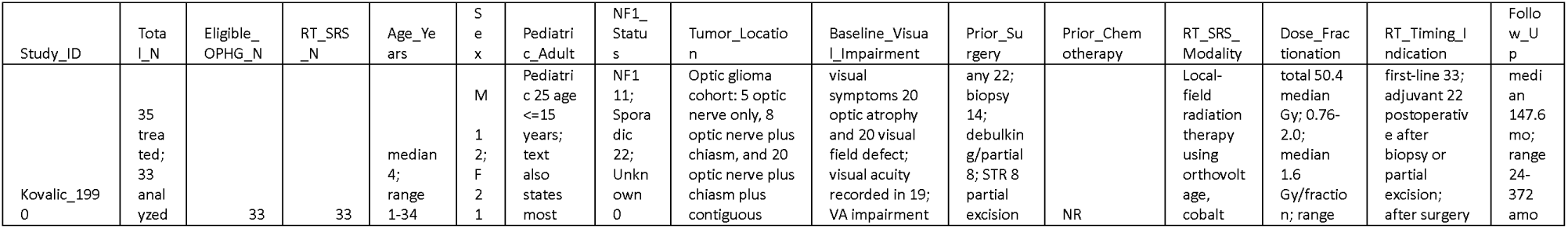
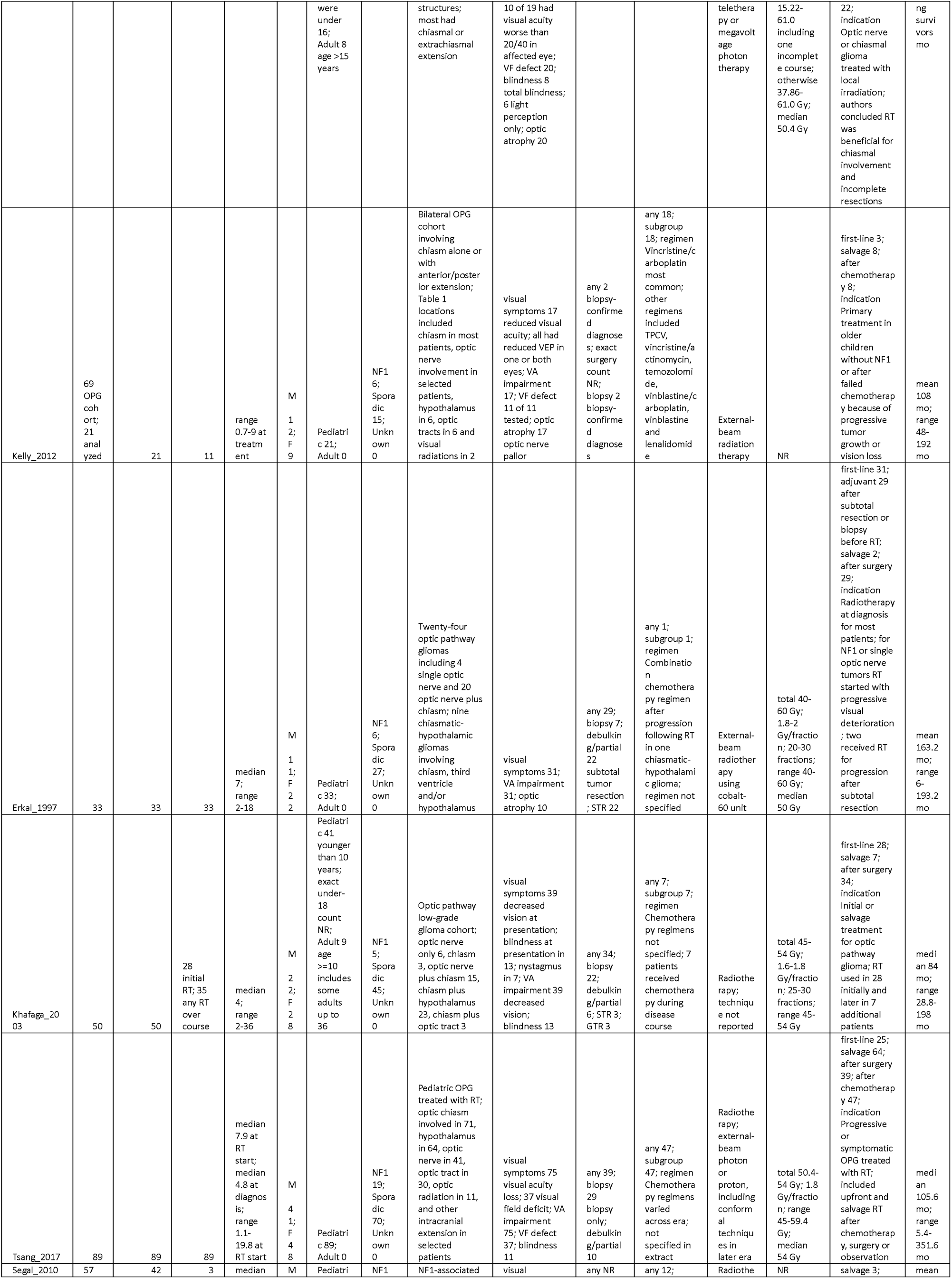
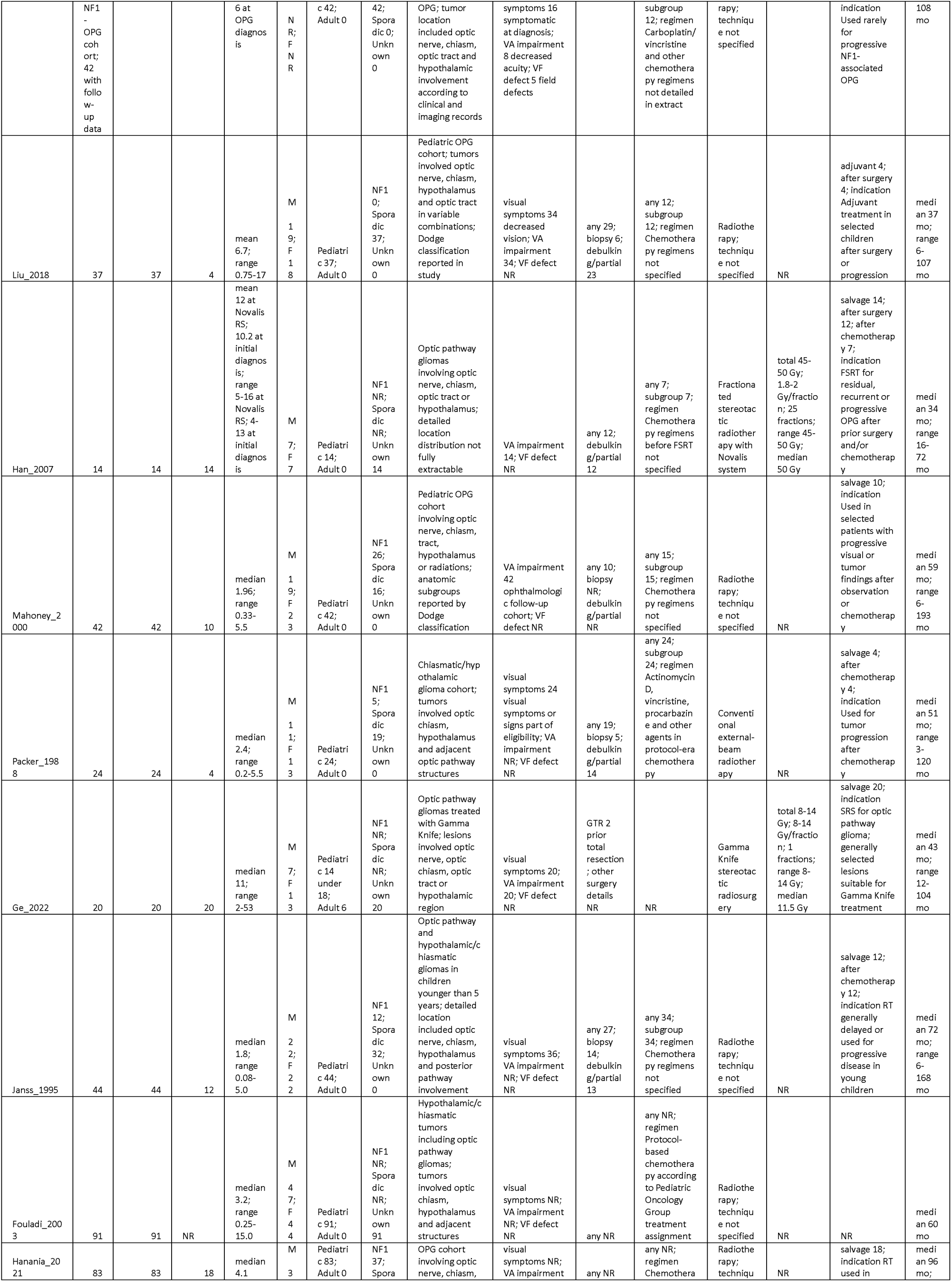
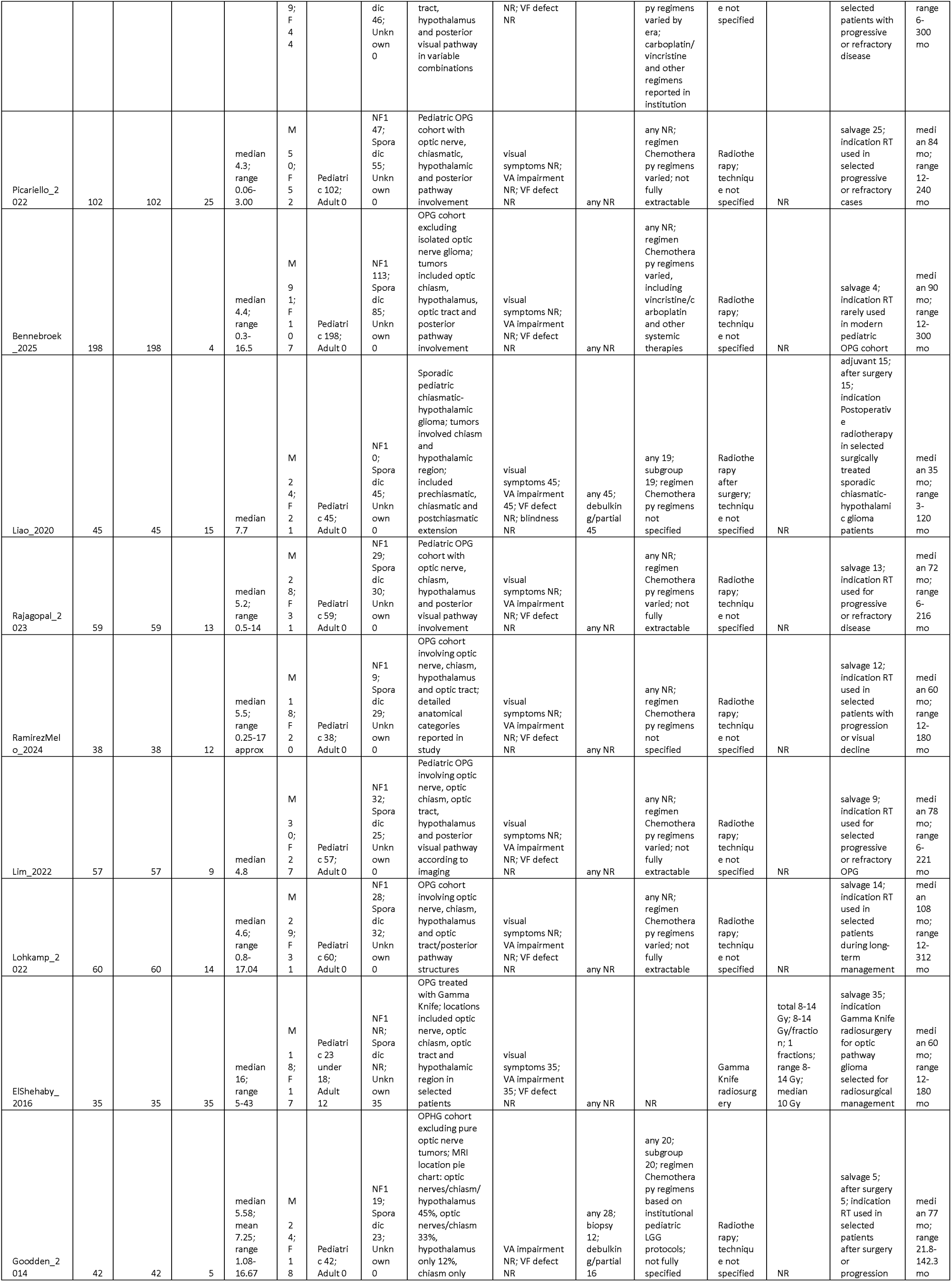
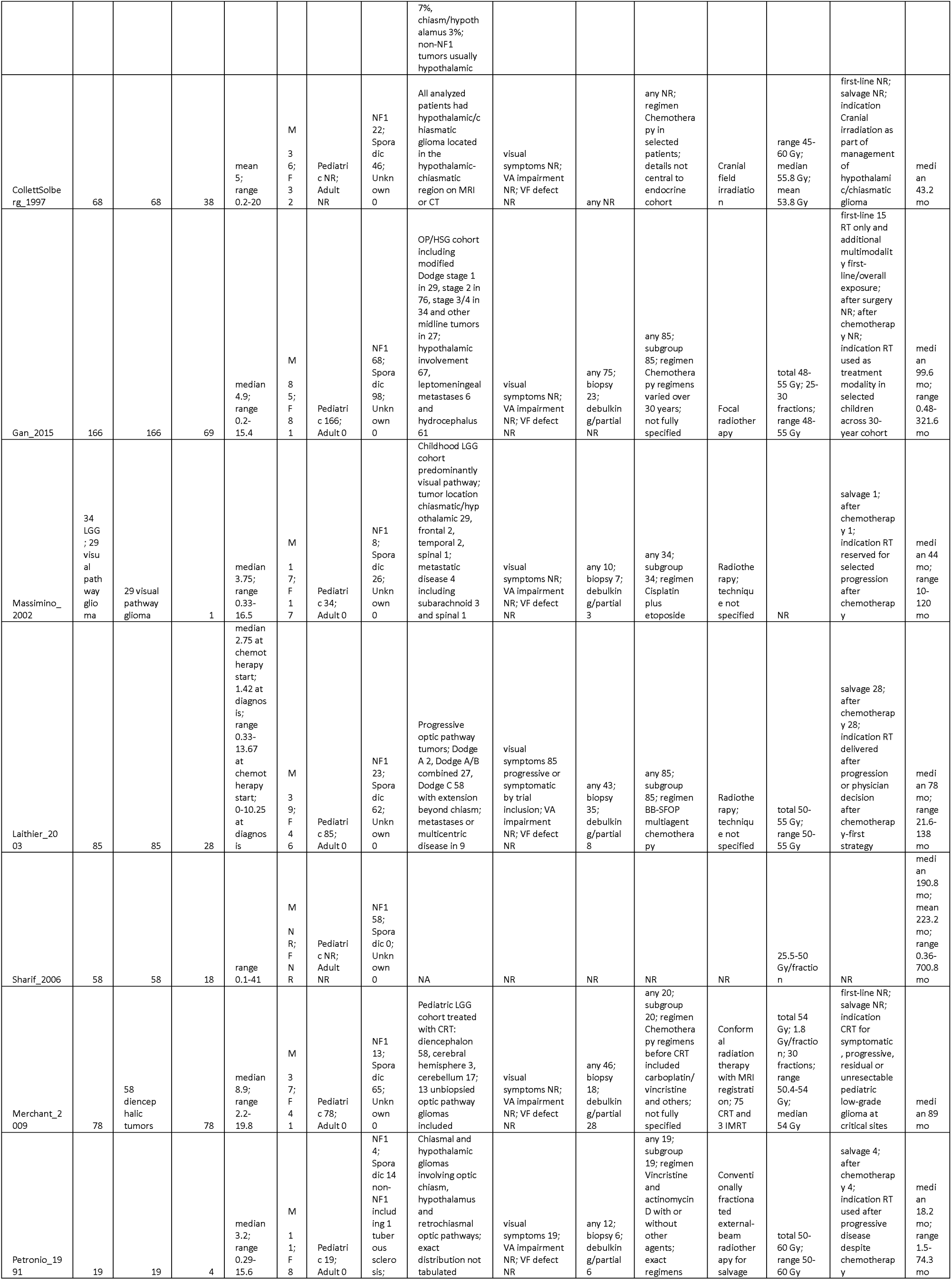
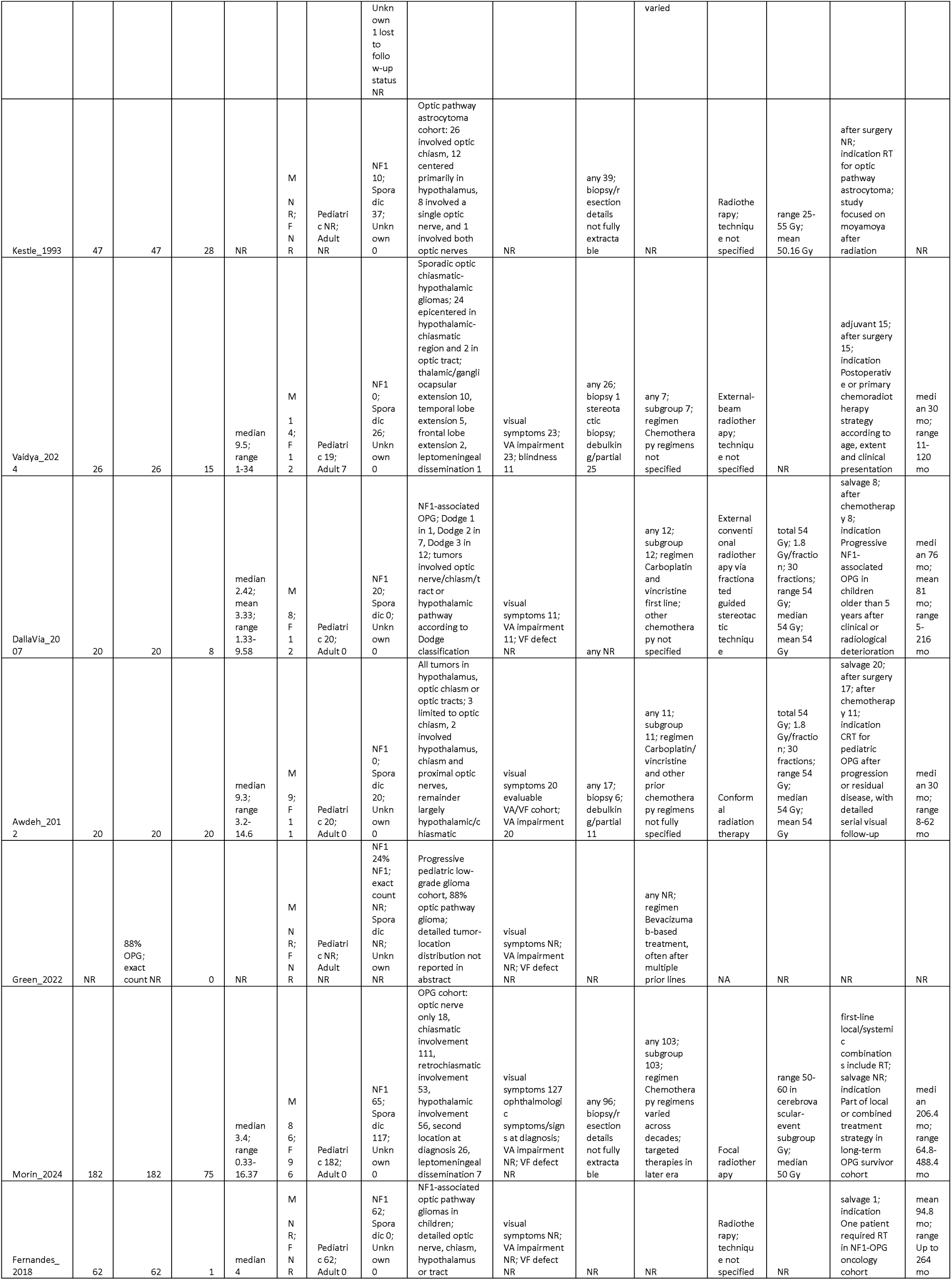
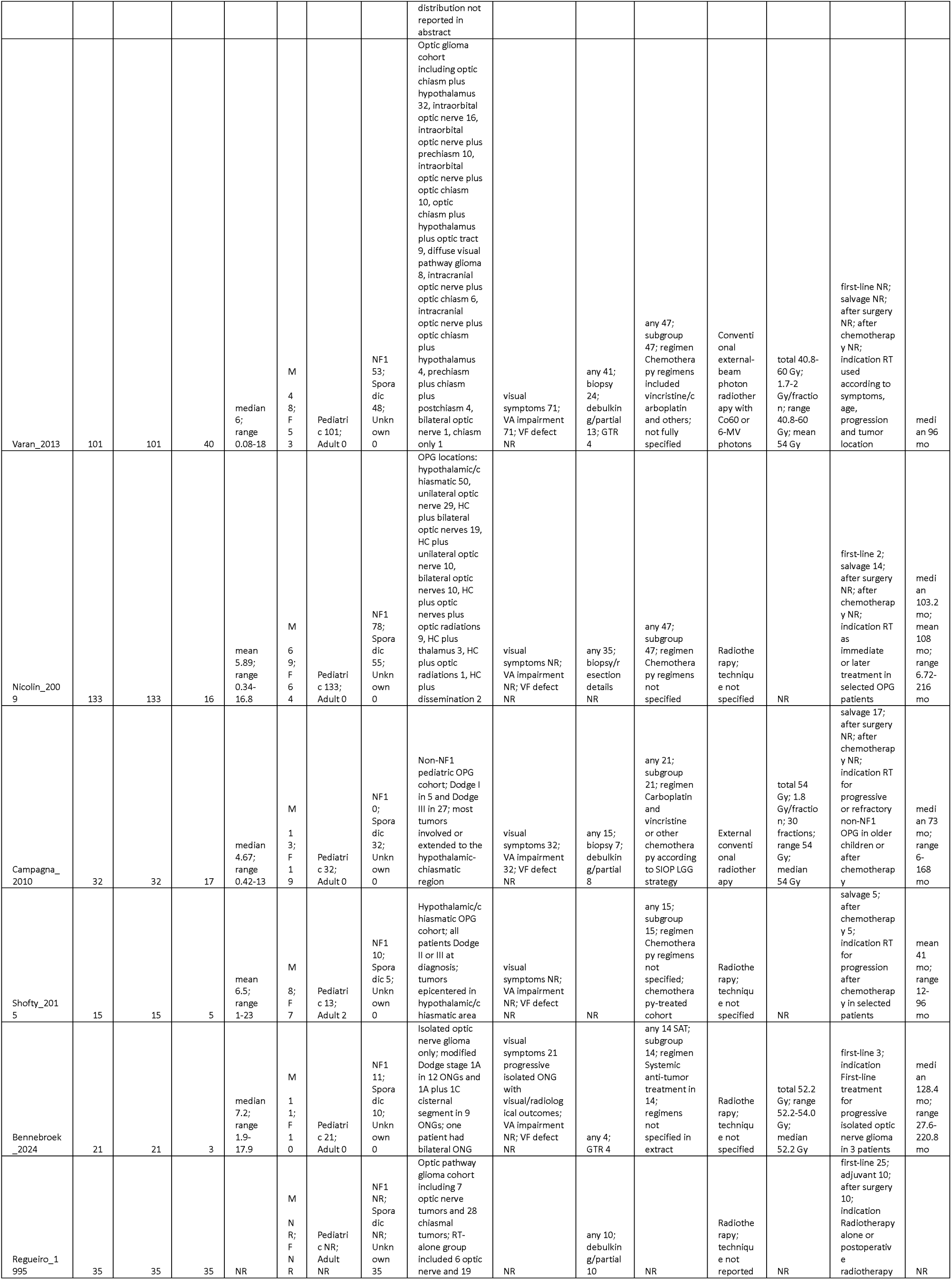
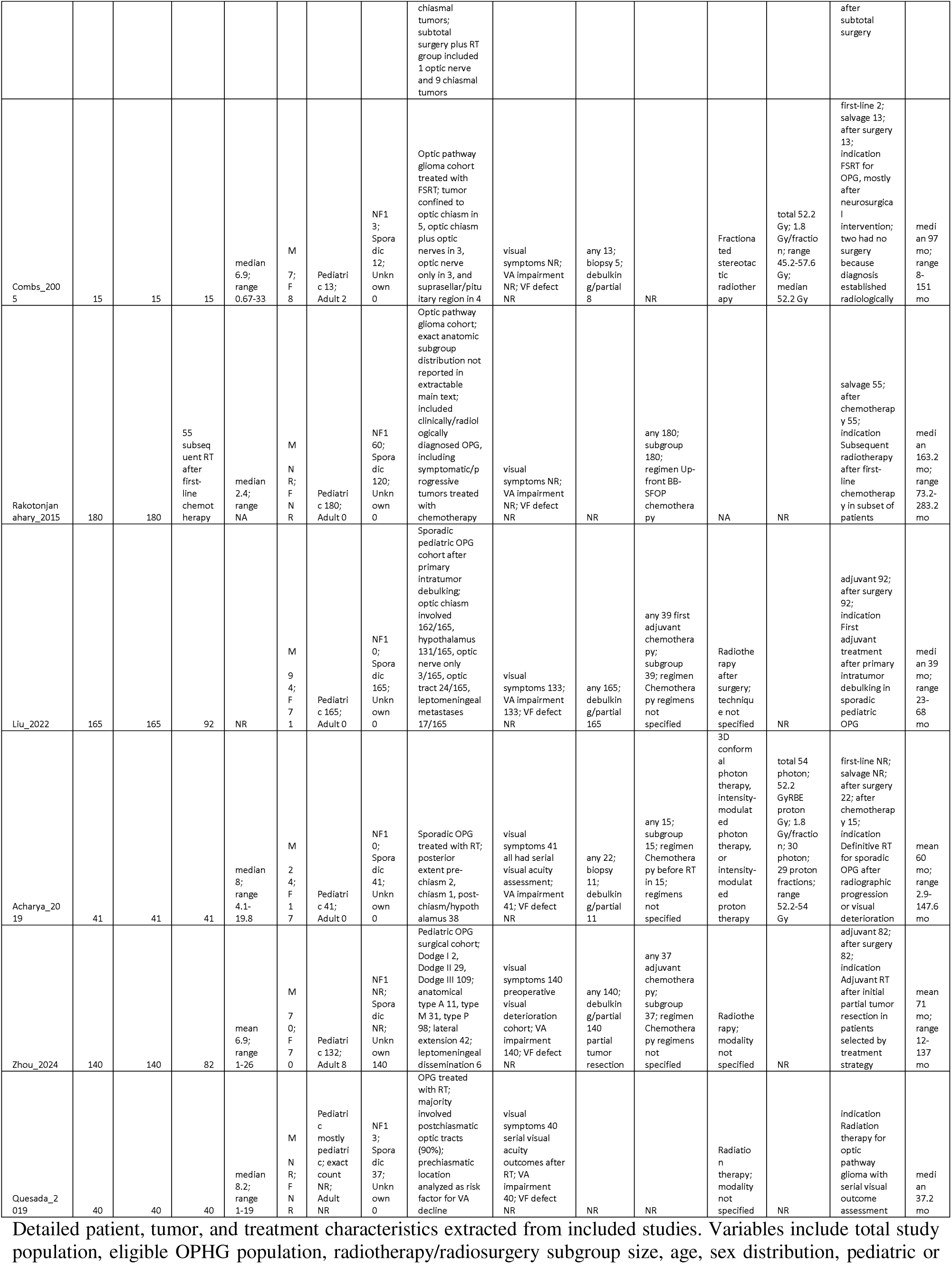
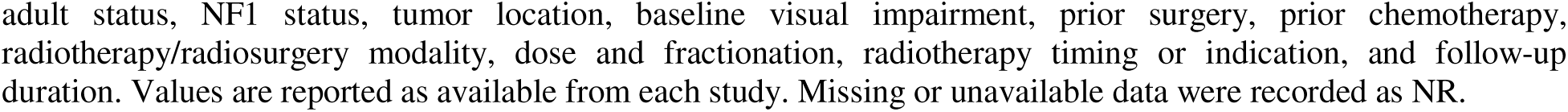
Patient, tumor, and treatment characteristics.

### Radiotherapy, radiosurgery, and concomitant treatment characteristics

Radiation treatment varied markedly across eras and institutions. Historical series predominantly used conventional photon radiotherapy, local-field cobalt irradiation, or megavoltage external-beam radiotherapy. Contemporary reports incorporated three-dimensional conformal radiotherapy, intensity-modulated radiotherapy, proton therapy, fractionated stereotactic radiotherapy, stereotactic radiosurgery, and Gamma Knife radiosurgery.

Where modality-specific numeric data were extractable, conventional photon radiotherapy was reported for 675 patients across 17 studies, three-dimensional conformal radiotherapy for 144 patients across three studies, intensity-modulated radiotherapy for 12 patients across two studies, proton therapy for 26 patients across three studies, fractionated stereotactic radiotherapy for 102 patients across four studies, and SRS/Gamma Knife for 78 patients across four studies. These categories were not mutually exclusive because several cohorts included more than one radiation technique or spanned transitions between treatment eras.

Conventionally fractionated treatment most commonly used total doses of approximately 45–54 Gy, frequently delivered in fractions of approximately 1.5–2.0 Gy. Some historical cohorts used broader dose ranges, whereas SRS/Gamma Knife series generally used single-session marginal doses of approximately 8–18 Gy. Target delineation, field size, optic apparatus constraints, and dosimetric reporting were inconsistent, particularly in older studies.

Radiotherapy or radiosurgery was delivered in several clinical contexts. Numeric treatment-timing data indicated first-line RT in 237 patients across 18 studies, adjuvant RT in 370 patients across 10 studies, salvage RT in 222 patients across 17 studies, RT after surgery in 615 patients across 18 studies, and RT after chemotherapy in 193 patients across 17 studies. These categories overlapped because individual patients could have received surgery, chemotherapy, and radiotherapy sequentially.

Some studies consisted exclusively of RT/SRS-treated patients, whereas others reported radiation-treated subgroups within broader cohorts managed with surgery, chemotherapy, observation, or multimodal treatment. Among the 19 studies included in the visual meta-analysis, seven were classified as pure RT/SRS cohorts, 11 were RT-containing mixed-treatment cohorts, and one was an abstract or limited-report cohort. This distinction informed the study-purity subgroup analysis.

### Visual outcome assessment and quantitative eligibility

Nineteen studies reported sufficiently complete categorical visual outcome data for quantitative synthesis [1, 4, 12–14, 17, 23–28, 34–40].

Visual outcomes were assessed using heterogeneous methods, including formal visual acuity testing, visual field examination, comprehensive ophthalmological assessment, blindness status, author-defined global visual categories, or combinations of these measures. Assessment timing also varied, ranging from early post-treatment evaluation to visual status at the last available follow-up.

The quantitative dataset comprised 494 evaluable visual-outcome observations. A visual-outcome observation represented the unit used by the original study to report an evaluable post-treatment visual result and could correspond to an individual patient, an individual eye, or another explicitly defined visual assessment unit. Patient-level outcomes were prioritized whenever available. Because some reports used eye-level or other observation-level denominators, the 494 observations should not be interpreted as 494 unique patients.

Visual preservation was defined as stable or improved vision. When improvement and stability were reported separately, preservation was calculated as their sum. Improvement represented a favorable change in visual acuity, visual fields, blindness status, global ophthalmological assessment, or an author-defined categorical outcome. Stability represented no clinically meaningful deterioration in the reported visual measure, while worsening represented a decline in one or more of these domains. The study-level distribution of improved, stable, and worsened visual outcomes is shown in Figure 2.

**Figure 2.**
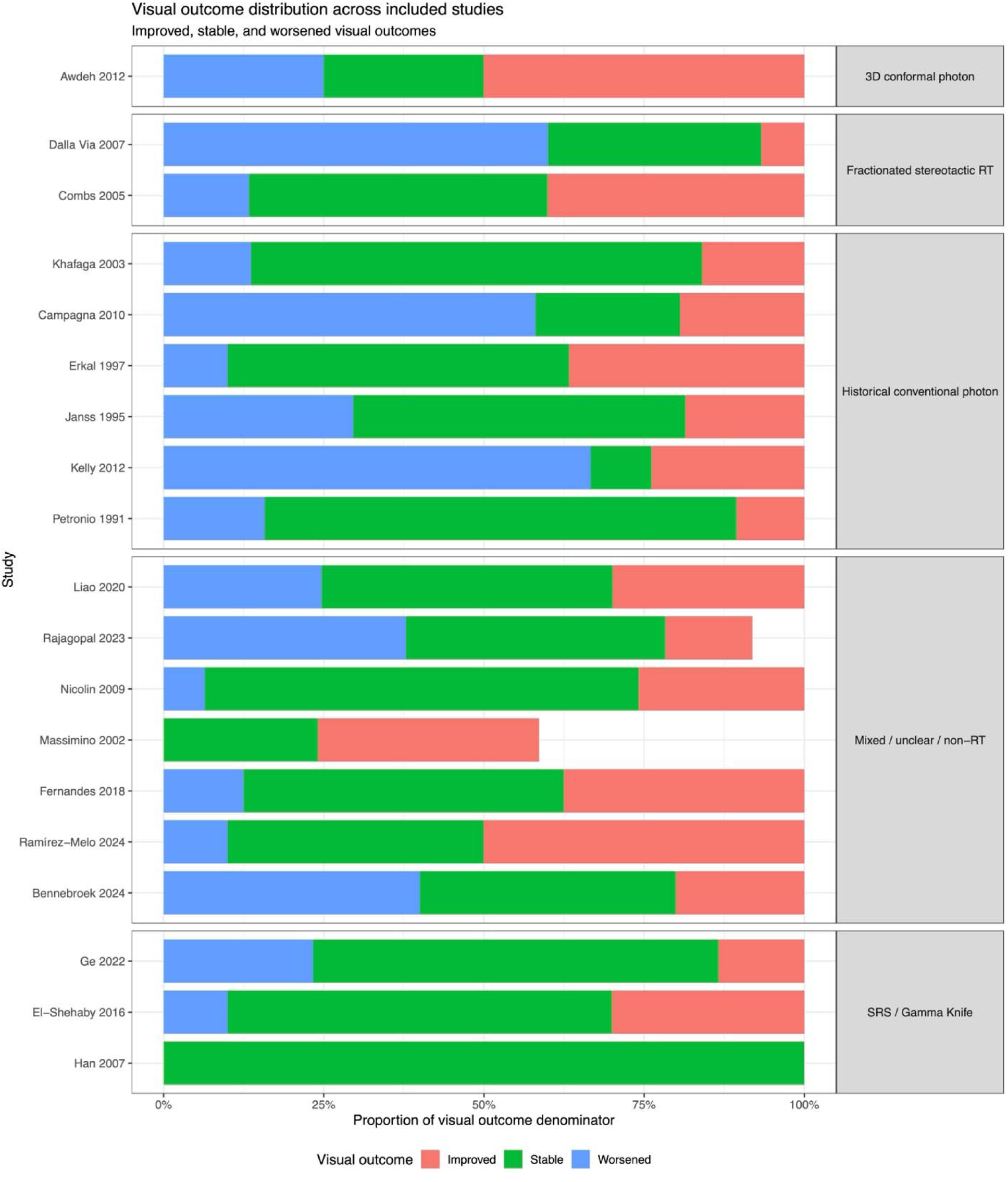
Distribution of visual outcomes across included studies. Study-level distribution of improved, stable, and worsened visual outcomes among studies included in the quantitative synthesis. Visual preservation was defined as the combination of stable and improved vision.

### Primary outcome: visual preservation

Across the 19 quantitatively eligible studies, 359 of 494 visual-outcome observations were classified as preserved. The pooled proportion of visual preservation was 75.6% (95% CI, 65.1%–83.7%). Between-study heterogeneity was substantial (I^2^ = 71.6%; τ^2^ = 0.7654; Cochran Q p < 0.0001), and the prediction interval was wide (31.5%–95.4%), indicating considerable variation in the expected preservation rate across clinical settings, patient populations, treatment approaches, and visual outcome definitions. The primary forest plot is presented in Figure 3.

**Figure 3.**
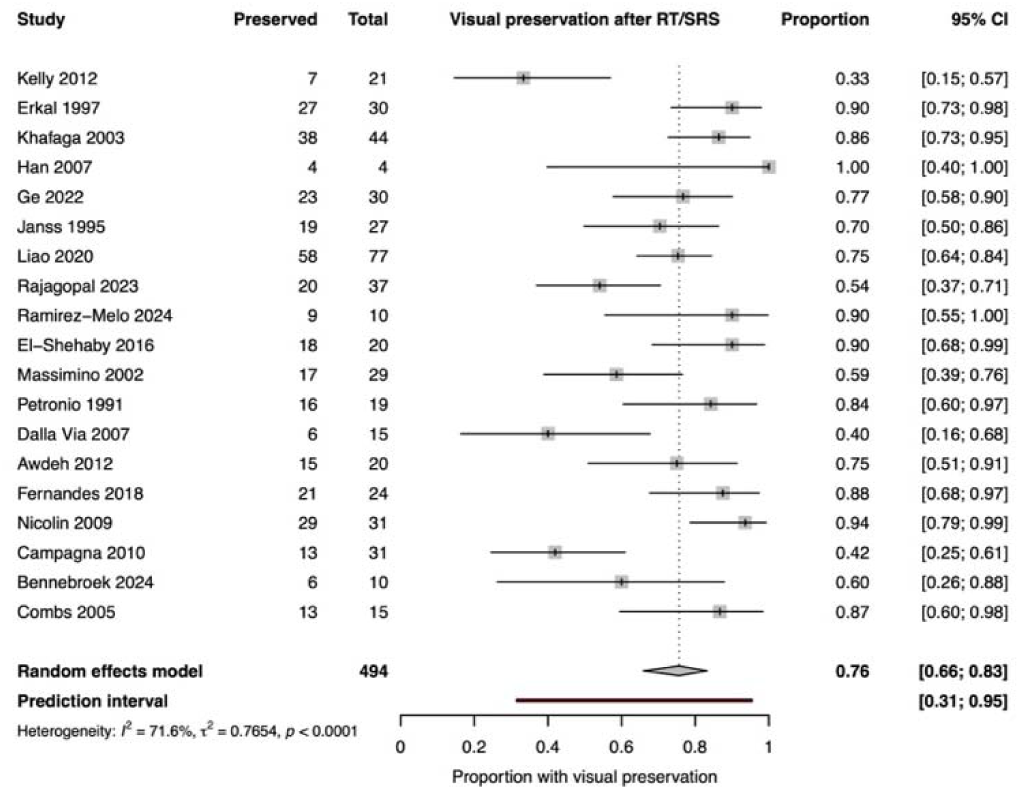
Forest plot of visual preservation after radiotherapy or radiosurgery. Random-effects meta-analysis of visual preservation following radiotherapy or radiosurgery. Visual preservation was defined as stable or improved vision.

Subgroup analysis did not identify a statistically significant difference according to radiation technique (p = 0.7269). Pooled visual preservation was 71% in historical conventional photon radiotherapy cohorts, 83% in SRS/Gamma Knife cohorts, 76% in mixed or insufficiently specified radiation cohorts, and 66% in fractionated stereotactic radiotherapy cohorts. The three-dimensional conformal photon category was represented by a single study with a preservation estimate of 75%. Because only three studies contributed to the SRS/Gamma Knife subgroup, the apparent favorable estimate should be regarded as exploratory and may reflect selection of smaller, anatomically suitable lesions rather than a modality-specific treatment advantage.

Study purity was associated with a statistically significant subgroup difference (p = 0.0104). Pure RT/SRS cohorts had a pooled visual preservation estimate of 85% (95% CI, 76%–90%), compared with 66% (95% CI, 50%–79%) in RT-containing mixed-treatment cohorts. Although this was the clearest radiation-specific subgroup signal, it may also reflect differences in cohort composition, treatment attribution, patient selection, and reporting completeness. The study-purity analysis is presented in Figure 4.

**Figure 4.**
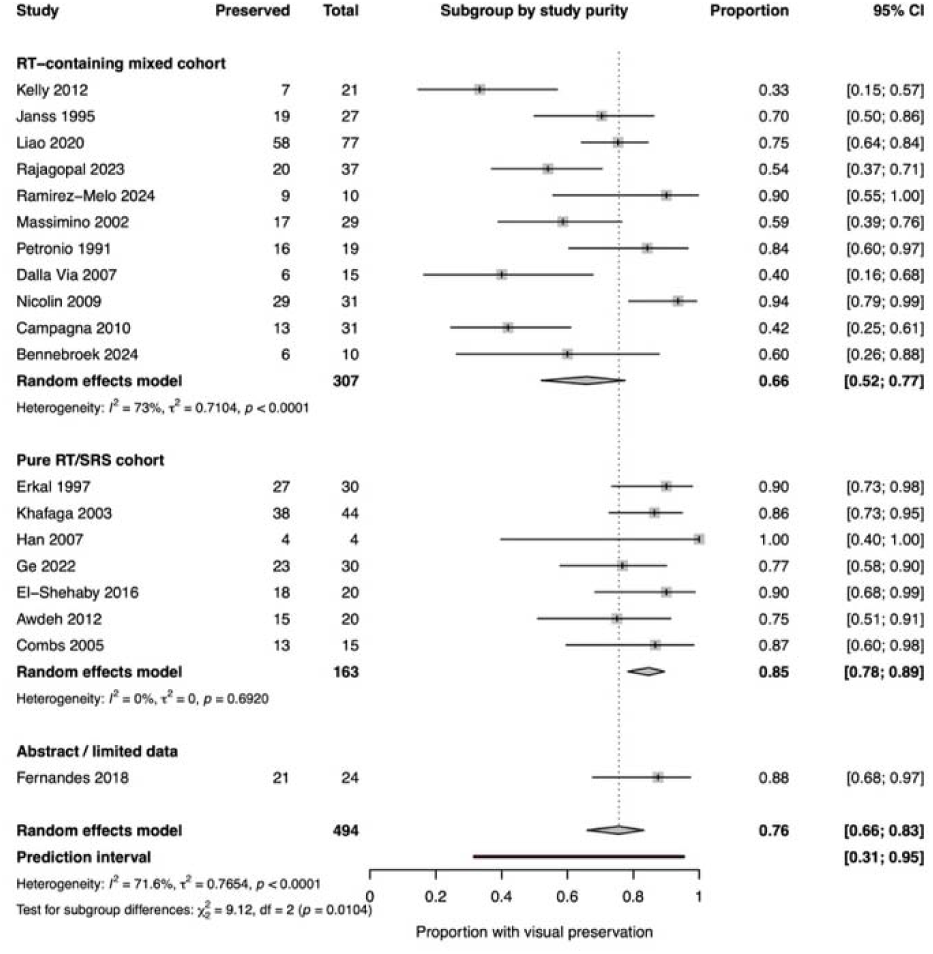
Visual preservation according to study purity. Subgroup analysis comparing visual preservation between pure radiotherapy/radiosurgery cohorts and mixed-treatment cohorts.

**Figure 5.**
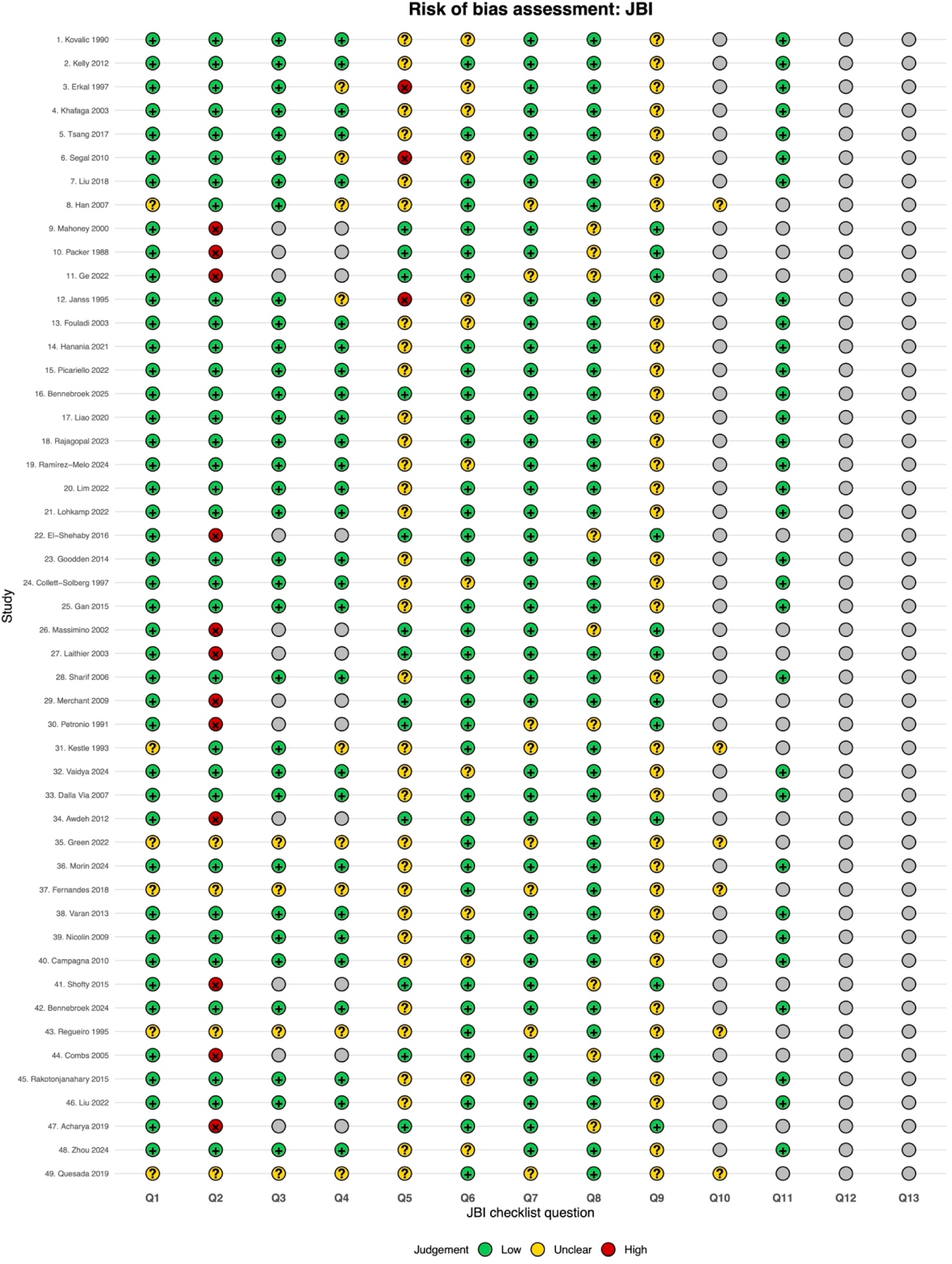
Risk-of-bias assessment. Traffic-light plot summarizing study-level risk of bias according to the Joanna Briggs Institute critical appraisal tools.

Radiotherapy timing showed a borderline subgroup difference (p = 0.0580). The pooled preservation estimate was 77% in salvage RT cohorts, 63% in primary or upfront RT cohorts, and 89% in cohorts with mixed or unclear treatment timing. These comparisons were supported by small and heterogeneous subgroups and should not be interpreted causally. No statistically significant difference was observed according to risk-of-bias category (p = 0.1284).

Sensitivity analyses yielded estimates similar to the primary model. Exclusion of high-risk-of-bias studies resulted in pooled visual preservation of 73% (95% CI, 60%–83%). Restriction to full-text studies yielded an estimate of 75% (95% CI, 64%–83%). Restriction to pure RT/SRS cohorts produced a pooled estimate of 85% (95% CI, 76%–90%) with no observed heterogeneity (I^2^ = 0%). Leave-one-out estimates remained between approximately 74% and 77%, suggesting that the pooled result was not dependent on any single study.

### Secondary visual outcomes

#### Visual improvement

Across the 19 studies, 125 of 494 visual-outcome observations were classified as improved. The pooled proportion of visual improvement was 24.7% (95% CI, 19.5%–30.8%), with low-to-moderate heterogeneity (I^2^ = 36.3%; τ^2^ = 0.1499; p = 0.0583) and a prediction interval of 12.1%– 43.9%.

Radiation technique was associated with a statistically significant subgroup difference (p = 0.0446), although several categories contained few studies. The SRS/Gamma Knife subgroup had a pooled improvement estimate of 19%, whereas the three-dimensional conformal photon subgroup was represented by a single study with an estimate of 50%. No statistically significant differences were observed according to risk-of-bias category, radiotherapy timing, or study purity. Sensitivity analyses excluding high-risk-of-bias studies, restricting the analysis to full-text reports, and sequentially omitting individual studies produced broadly similar estimates.

#### Visual stability

Visual stability was reported for 234 of 494 observations. The pooled proportion was 46.7% (95% CI, 37.0%–56.6%), with substantial heterogeneity (I^2^ = 64.1%; τ^2^ = 0.4668; p < 0.0001) and a prediction interval of 16.5%–79.5%.

Subgroup differences were observed according to risk-of-bias category (p = 0.0126), radiation technique (p = 0.0265), and radiotherapy timing (p = 0.0041). The SRS/Gamma Knife subgroup had a pooled stability estimate of 65%. Pure RT/SRS cohorts had a numerically higher stability estimate than RT-containing mixed cohorts (57% vs 40%), although this subgroup difference was not statistically significant (p = 0.1572). Sensitivity and leave-one-out analyses produced estimates similar to the main model.

#### Visual worsening

Visual worsening occurred in 120 of 494 evaluable observations. The pooled proportion was 20.6% (95% CI, 12.7%–31.6%), with substantial heterogeneity (I^2^ = 69.1%; τ^2^ = 1.0220; p < 0.0001) and a wide prediction interval of 2.8%–70.0%.

Radiotherapy timing was associated with a borderline statistically significant subgroup difference (p = 0.0498). The pooled worsening estimate was 35% in primary or upfront RT cohorts and 14% in salvage RT cohorts. This difference should not be interpreted as evidence that upfront radiotherapy increases the risk of visual deterioration because it may reflect confounding by indication, baseline visual status, disease severity, treatment era, or differences in subgroup composition. No statistically significant subgroup differences were observed according to radiation technique, study purity, or risk-of-bias category.

Among pure RT/SRS cohorts, the pooled proportion of visual worsening was 15% (95% CI, 10%–24%), with no observed heterogeneity (I^2^ = 0%). Leave-one-out estimates remained between approximately 19% and 23%.

The primary and secondary pooled outcomes are summarized in Table 3. Key subgroup and sensitivity findings are presented in Table 4. Supplementary forest plots and leave-one-out analyses are provided in Supplementary File 9, and detailed subgroup and sensitivity analyses are provided in Supplementary File 10.

**Table 3.**
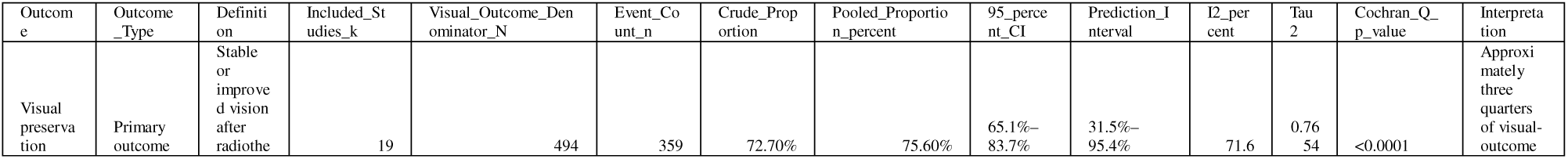
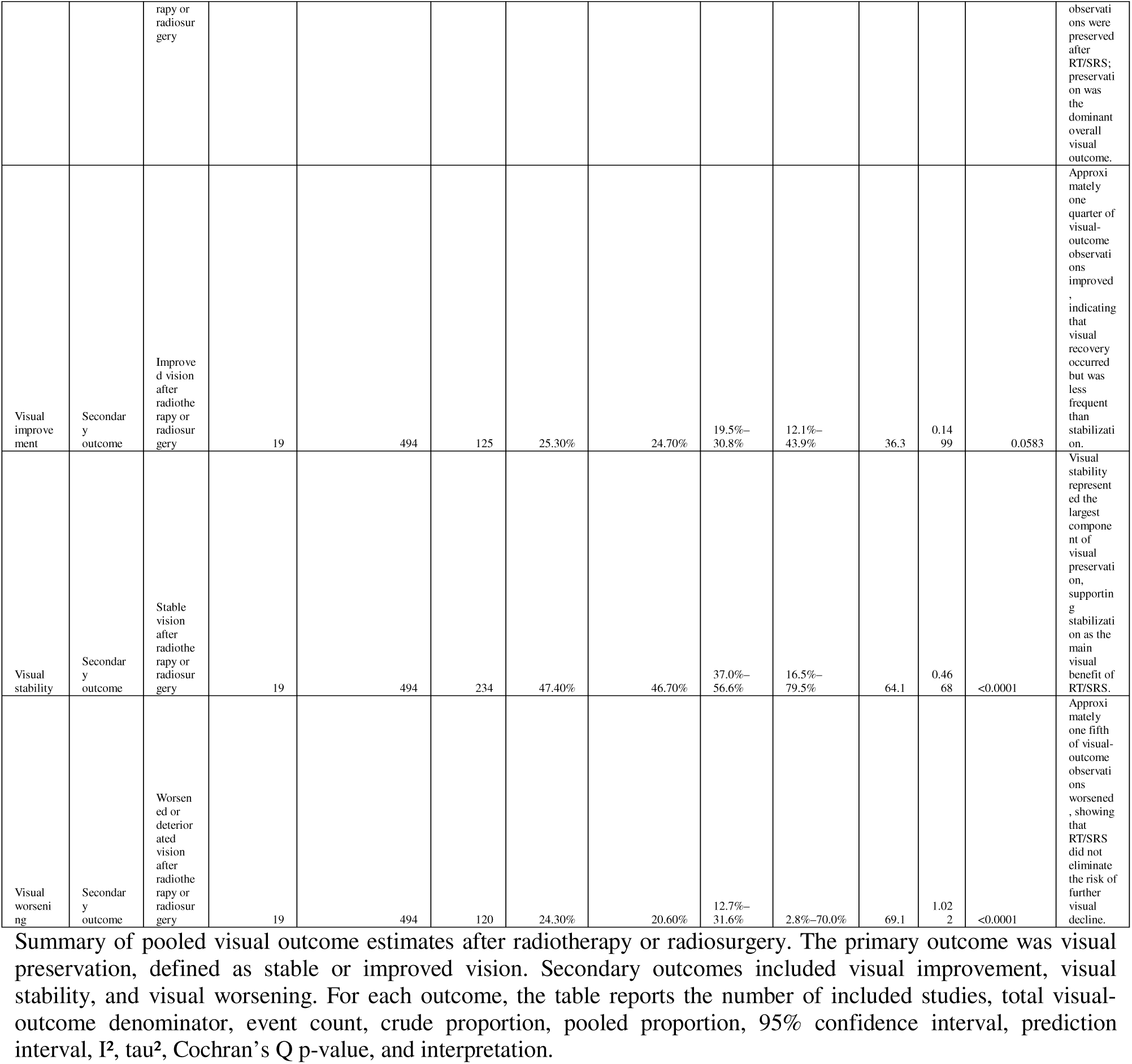
Summary of visual outcome meta-analyses.

**Table 4.**
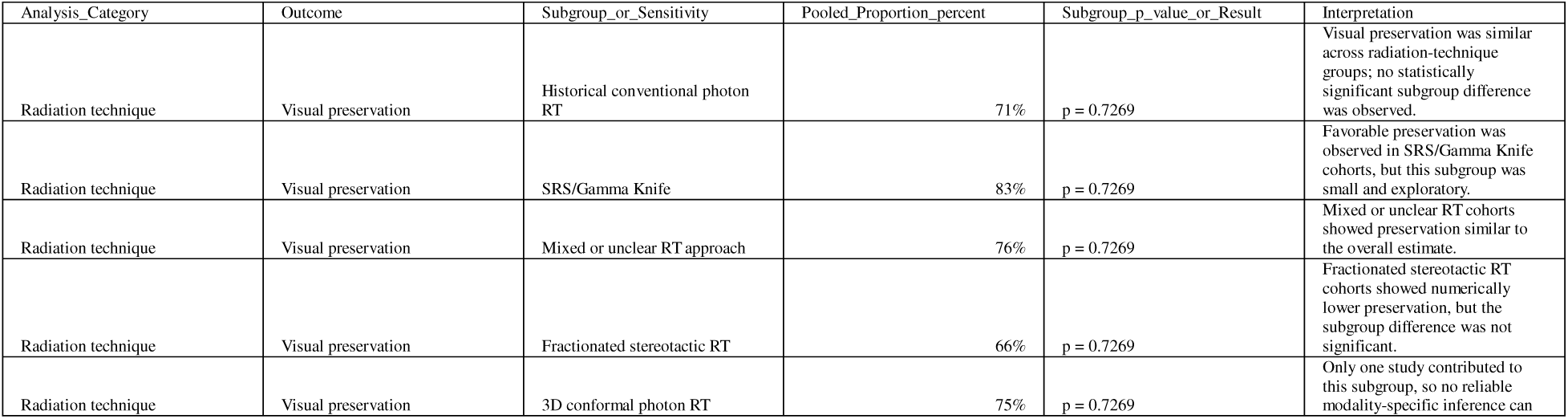
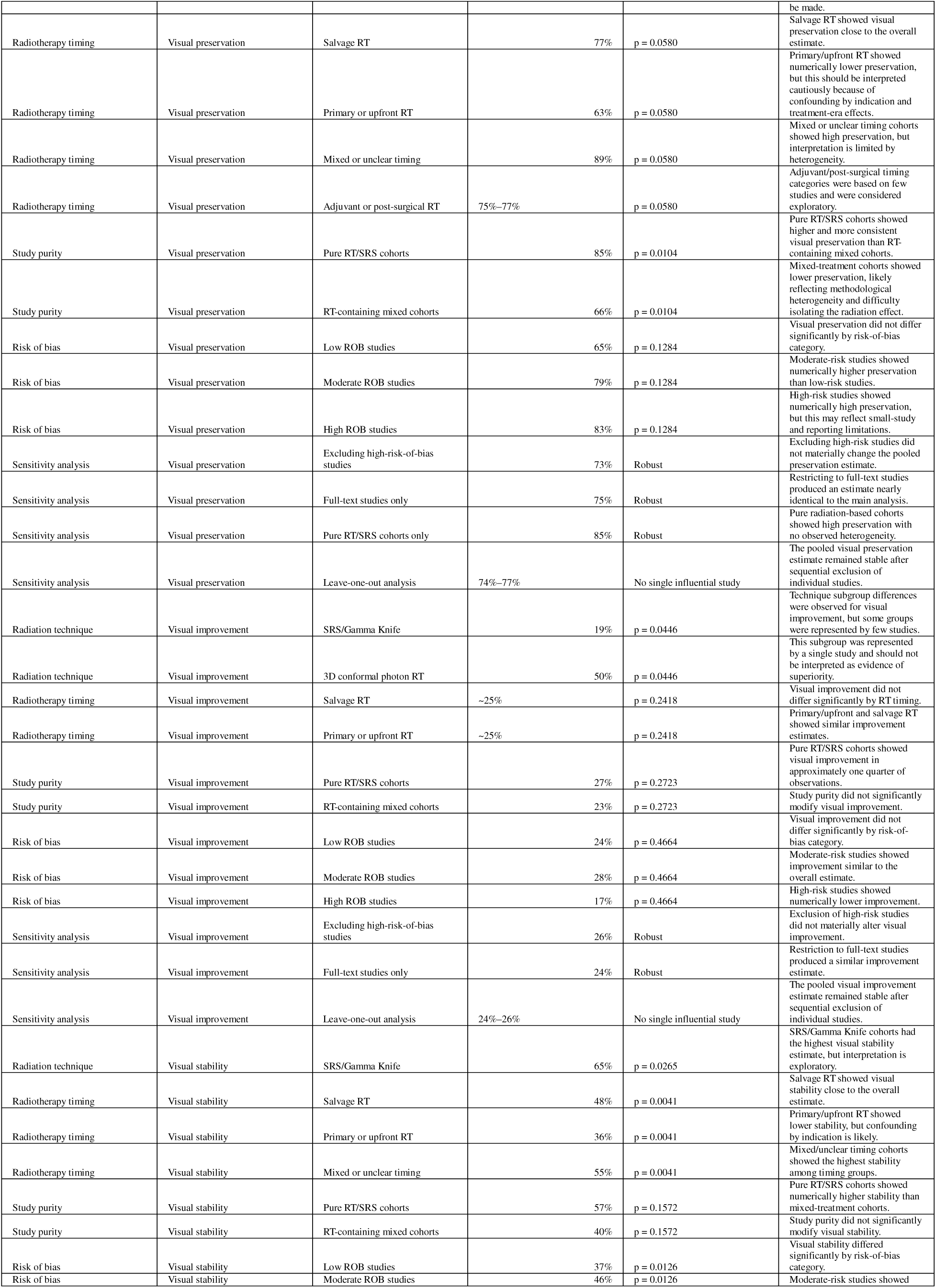
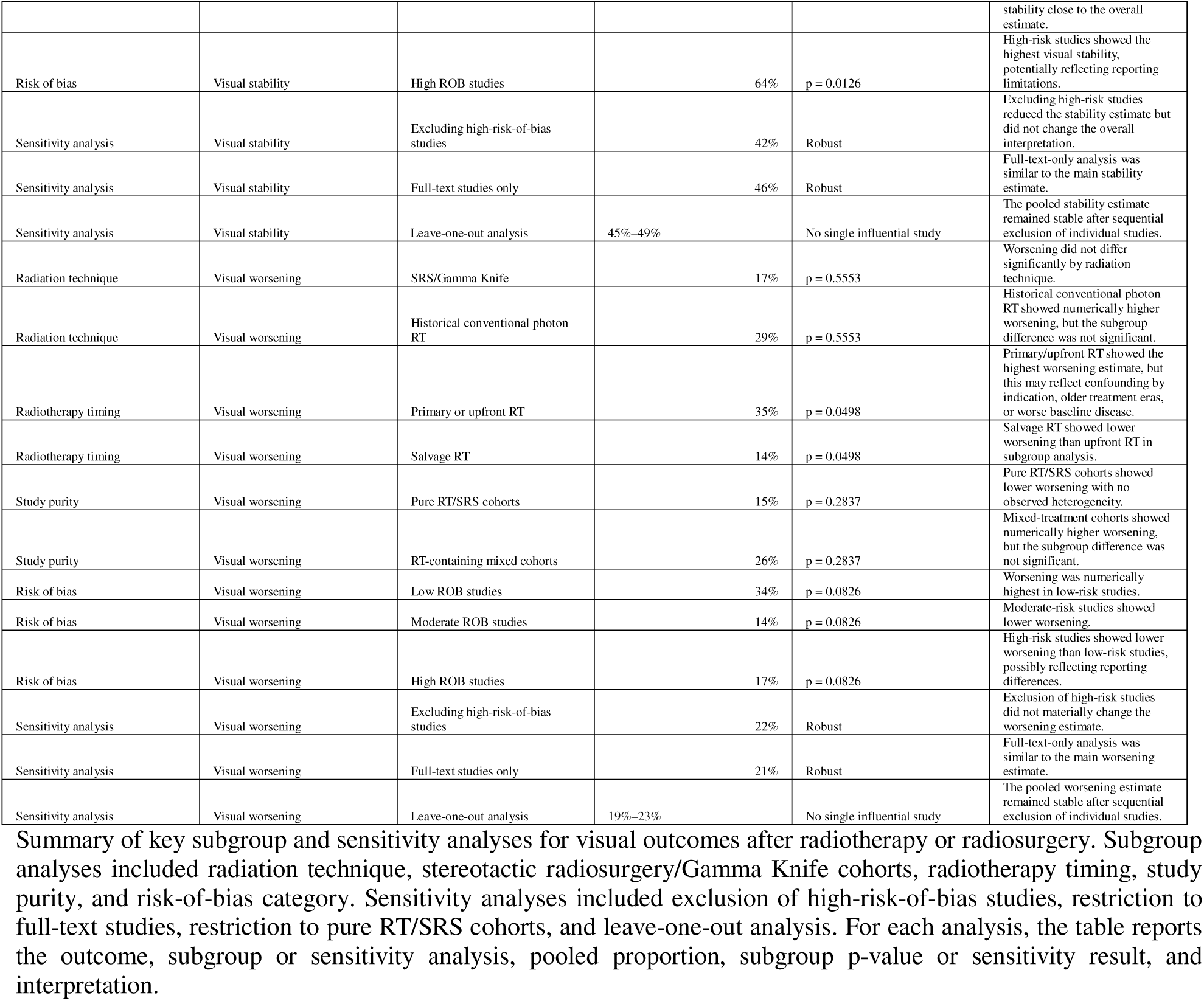
Key subgroup and sensitivity analyses.

### Small-study effects and publication-bias assessment

The funnel plot for visual preservation showed some visual asymmetry. Egger-type regression did not provide clear statistical evidence of funnel-plot asymmetry (intercept, 2.29; p = 0.060), although the test had limited power and cannot exclude small-study effects.Given the limited number of studies, substantial clinical heterogeneity, variable outcome definitions, and predominantly observational designs, funnel-plot asymmetry could reflect genuine between-study differences rather than publication bias alone. The funnel plot and related exploratory analyses are provided in Supplementary File 9.

### Risk-of-bias assessment

Among the 49 included studies, 21 were categorized as having low risk of bias, 20 as moderate risk, and eight as high risk. Common methodological concerns included retrospective patient selection, limited adjustment for confounding, incomplete characterization of baseline visual status, variable follow-up, incomplete outcome ascertainment, small study populations, and limited methodological detail in abstract-only reports.

Within the 19-study quantitative visual synthesis, seven studies were categorized as low risk of bias: Kelly et al. (2012), Liao et al. (2020), Rajagopal et al. (2023), Dalla Via et al. (2007), Awdeh et al. (2012), Nicolin et al. (2009), and Bennebroek et al. (2024). Eight were categorized as moderate risk: Erkal et al. (1997), Khafaga et al. (2003), Janss et al. (1995), Ramírez-Melo et al. (2024), El-Shehaby et al. (2016), Massimino et al. (2002), Campagna et al. (2010), and Combs et al. (2005). Four were categorized as high risk: Han et al. (2007), Ge et al. (2022), Petronio et al. (1991), and Fernandes et al. (2018).

High-risk studies were most commonly limited by small sample size, abstract-only reporting, insufficient methodological detail, incomplete follow-up information, or incomplete visual outcome reporting. The traffic-light plot is presented in Figure 5, and the complete item-level assessment is provided in Supplementary File 8.

## Discussion

### Principal findings

In this systematic review and meta-analysis, radiotherapy and radiosurgery were associated with preservation of visual function in approximately three quarters of evaluable visual outcomes among patients with optic pathway hypothalamic glioma. Across 19 studies and 494 visual-outcome observations, the pooled proportion of visual preservation was 75.6%, while visual improvement, stability, and worsening were 24.7%, 46.7%, and 20.6%, respectively. These findings suggest that the principal visual benefit of radiation-based treatment may lie in maintaining existing visual function rather than reliably restoring established visual loss. Visual stability occurred almost twice as frequently as visual improvement, supporting stabilization as the predominant component of the overall preservation estimate.

This distinction is clinically important. Visual decline in OPHG may reflect tumor infiltration, compression of the optic pathways, delayed treatment, axonal injury, optic atrophy, vascular compromise, or irreversible retinal ganglion cell loss. Once substantial structural damage has occurred, the potential for meaningful recovery may be limited. Accordingly, prevention of further deterioration may still represent an important therapeutic benefit, even when measurable visual improvement is uncommon. Nevertheless, approximately one fifth of evaluable visual outcomes worsened despite treatment, indicating that radiotherapy and radiosurgery cannot be expected to prevent visual decline in all patients.

The quantitative synthesis was based on visual-outcome observations rather than exclusively on unique patients. Depending on the source study, an observation could represent an individual patient, an eye, or another explicitly defined evaluable visual unit. This approach allowed inclusion of a heterogeneous historical literature but also limits direct patient-level interpretation and underscores the need for standardized reporting of visual outcomes in future studies.

### Biological and pathophysiological interpretation

The predominance of visual stability over improvement is biologically plausible given the mechanisms of visual pathway injury in OPHG. These tumors involve eloquent structures, including the optic nerve, chiasm, optic tract, hypothalamus, and adjacent deep midline regions. Visual dysfunction may arise through direct tumor infiltration, compression of the optic nerves or chiasm, disruption of axonal transport, vascular compromise, gliosis, demyelination, and secondary optic atrophy[1, 4–7, 11, 25–28, 33, 34]. These mechanisms are not uniformly reversible, particularly when longstanding optic atrophy or retinal ganglion cell loss has already developed.

Radiotherapy and radiosurgery are thought to preserve visual function primarily through local tumor control, thereby limiting further compression, infiltration, or anatomical disruption of the visual pathway. Conventional fractionated radiotherapy permits treatment of infiltrative disease while limiting the dose delivered per fraction to adjacent critical structures. Modern conformal, intensity-modulated, proton, and stereotactic techniques may improve dose localization and reduce unnecessary irradiation of surrounding brain, endocrine structures, and vasculature. Radiosurgery and Gamma Knife approaches provide highly focused dose delivery and may be appropriate for selected small-volume, well-defined lesions, although treatment near the optic apparatus requires strict dose constraints and careful anatomical selection.[12–17, 22, 35, 41–43]

The difference between tumor control and neural recovery may explain why stability was more common than improvement. Radiation may arrest progression, but visual recovery depends on the presence of viable axons, preserved retinal ganglion cells, intact conduction through the optic pathways, and sufficient time for functional improvement. In patients with severe baseline visual loss, optic pallor, prolonged symptom duration, or extensive chiasmatic-hypothalamic involvement, the opportunity for recovery may already be limited. These considerations suggest that treatment timing could be clinically important, although the present aggregate data cannot define a precise threshold beyond which visual recovery becomes unlikely.

### Comparison with previous literature

Previous OPHG literature has largely emphasized tumor control, progression-free survival, endocrine morbidity, and overall treatment strategy rather than visual outcome as a primary endpoint. Earlier radiotherapy series suggested that radiation could provide durable disease control in selected patients, but visual outcomes were frequently reported as secondary, descriptive, or incompletely defined endpoints [23, 24, 36, 38, 41, 42, 44, 45] . Later studies incorporated chemotherapy, conformal radiotherapy, proton therapy, fractionated stereotactic radiotherapy, and radiosurgery, yet visual reporting remained inconsistent across treatment eras and modalities [12–15, 17, 25–27, 35, 43] .

The present study differs from prior reviews by focusing specifically on visual outcomes after radiotherapy and radiosurgery. Previous reviews of optic pathway glioma management have generally evaluated treatment modalities broadly, often combining radiological response, clinical response, survival, and visual outcomes. Although informative for overall management, such approaches may obscure the question most relevant to patients and families: whether treatment is likely to preserve, improve, or fail to protect vision. By separating preservation into improvement and stability and by quantifying worsening independently, this review provides a more clinically interpretable framework.

The pooled preservation estimate of approximately 76% is compatible with the possibility that radiation-based treatment stabilizes visual function in many patients. However, most of this apparent benefit was attributable to stability rather than improvement. A single composite category of “favorable visual outcome” may therefore be misleading if it combines recovery with absence of deterioration. For clinical counseling, improvement and stability should be reported separately, because preservation of function may constitute a meaningful therapeutic success but should not be equated with reversal of established injury.

### Radiation technique and subgroup findings

Subgroup analyses suggested that treatment context may influence visual outcomes, although these findings should be interpreted cautiously. SRS/Gamma Knife cohorts showed pooled estimates of approximately 83% for preservation, 65% for stability, 19% for improvement, and 17% for worsening. These results are encouraging but exploratory because only a small number of studies contributed to this subgroup. Moreover, radiosurgery is typically reserved for selected small, well-circumscribed lesions with favorable anatomy and acceptable optic apparatus dose constraints. The observed outcomes may therefore reflect patient and lesion selection rather than an intrinsic advantage of radiosurgery[14, 17, 35].

The pure RT/SRS cohort analysis produced one of the clearest subgroup signals observed in this review. In these treatment-specific cohorts, visual preservation was approximately 85%, visual improvement 27%, visual stability 57%, and visual worsening 15%. Mixed-treatment cohorts showed lower preservation and higher worsening. This should not be interpreted as evidence that multimodal treatment is inferior. Rather, mixed cohorts are methodologically less specific because visual outcomes may be influenced by surgery, chemotherapy, observation, delayed radiation, or complex treatment sequences. Pure RT/SRS cohorts may provide a more direct estimate of radiation-associated outcomes, whereas mixed cohorts may better reflect clinical practice but offer less certainty regarding attribution.

Treatment timing also appeared potentially relevant. Subgroup analyses suggested differences in visual stability and worsening according to whether radiation was delivered upfront, as salvage treatment, after surgery, or in mixed or unclear settings. However, these categories were heterogeneous and often contained few studies. The higher worsening estimate observed in some primary or upfront RT cohorts should not be interpreted as evidence of harm from early radiotherapy. It may instead reflect confounding by indication, older treatment eras, more aggressive disease, worse baseline vision, or selection of patients with rapidly progressive tumors[15, 16, 19, 21, 22, 29–31, 43, 46–49]. The timing signal should therefore be viewed as hypothesis-generating and highlights the need to determine when treatment is most likely to preserve vision before irreversible damage develops.

### Clinical implications

The principal clinical implication is that radiotherapy and radiosurgery may be more appropriately discussed as vision-preserving rather than reliably vision-restoring treatments. In patients with progressive OPHG, particularly those with declining visual acuity or visual fields despite observation, chemotherapy, or targeted therapy, the therapeutic objective may be to prevent further deterioration rather than to recover already lost function. This may be especially relevant when residual vision remains functionally important, when progression threatens bilateral visual loss, or when other treatment options have failed.

Visual outcomes should therefore be integrated more explicitly into multidisciplinary treatment decisions, particularly because radiological stability does not necessarily correspond to preservation of visual function[1, 4, 13, 15, 16, 25–28, 33, 34]. Conventional oncological endpoints such as radiological response and progression-free survival do not fully capture patient-centered benefit in OPHG. A tumor may remain radiologically stable while vision deteriorates, whereas treatment may still be clinically meaningful if it preserves visual function without producing substantial tumor shrinkage. Ophthalmological assessment should therefore be incorporated alongside imaging progression, endocrine risk, age, NF1 status, tumor anatomy, prior therapy, and anticipated treatment toxicity.

Selection of radiation technique also requires individualized judgment. Fractionated radiotherapy may be more suitable for larger or infiltrative disease involving the optic pathway, while stereotactic radiosurgery or Gamma Knife should remain restricted to carefully selected lesions with favorable anatomy, limited treatment volume, and acceptable optic apparatus dose constraints. Proton therapy and modern conformal techniques may reduce integral dose and late toxicity, particularly in children, but the available visual outcome literature is too heterogeneous to establish the superiority of any single modality.

### Visual preservation as a core endpoint

A major methodological implication of this review is that vision should not remain a secondary descriptive variable in OPHG research. Visual function is often a central determinant of long-term independence and quality of life, yet visual outcomes have been reported inconsistently across decades of literature. Some studies evaluated visual acuity, others visual fields, comprehensive ophthalmological findings, blindness status, or broad author-defined categories[1, 4, 11, 13, 15, 16, 25–28, 32, 34, 40]. Denominators, follow-up intervals, testing methods, and thresholds for meaningful change also varied substantially. Few studies incorporated objective retinal biomarkers such as optical coherence tomography despite their increasing clinical relevance.

The present findings suggest that visual preservation is measurable and clinically interpretable when reported systematically. Separating improvement, stability, and worsening provides more useful information than combining outcomes into broad favorable or unfavorable categories. Future OPHG studies should report visual outcomes at predefined intervals using standardized definitions, ideally including baseline and longitudinal visual acuity, visual fields, optic disc status, retinal nerve fiber layer and ganglion cell-inner plexiform layer measurements, and patient-reported visual function. Development of a standardized OPHG visual outcome core set would substantially improve future evidence synthesis, comparative effectiveness research, and guideline development.

### Future directions

Future research should move beyond determining whether radiotherapy or radiosurgery controls OPHG and instead address which patients are most likely to preserve vision, which treatment modality minimizes lifetime morbidity, and when radiation should be delivered before visual pathway injury becomes irreversible.

Prospective multicenter registries are needed because the rarity of OPHG limits the power and generalizability of single-institution retrospective series. An international registry focused on visual outcomes could capture baseline visual acuity, visual fields, OCT metrics, tumor location, hypothalamic involvement, NF1 status, molecular alterations, treatment sequence, radiation dose distribution, and long-term endocrine, vascular, neurocognitive, and quality-of-life outcomes. Such a resource would enable risk-adjusted analyses that are not possible using current aggregate data.[7, 30, 32]

Future studies should also incorporate modern imaging and visual pathway biomarkers. Diffusion MRI, tractography, volumetric tumor segmentation, optic nerve and chiasm dose-volume metrics, and OCT-derived retinal nerve fiber layer or ganglion cell-inner plexiform layer thickness may help identify patients at greatest risk of deterioration and distinguish potentially reversible dysfunction from established irreversible injury. Integration of radiological, ophthalmological, molecular, and dosimetric variables could support individualized prediction of visual preservation.

Radiation studies should report dose to the optic apparatus, hypothalamus, pituitary, circle of Willis, temporal lobes, and hippocampi using standardized methods. Late effects remain a major concern, particularly in children and patients with NF1, including endocrinopathy, vasculopathy, neurocognitive impairment, and secondary malignancy[8, 10, 20, 22, 31, 42, 43]. Comparisons of proton therapy, IMRT, conformal photon radiotherapy, fractionated stereotactic radiotherapy, and radiosurgery should therefore evaluate visual preservation together with tumor control and lifetime toxicity burden. The preferred radiation strategy is not simply the one associated with visual preservation, but the one that achieves durable control while minimizing long-term morbidity.

The timing of radiation also requires prospective evaluation. The field needs to determine whether a clinically meaningful “point of no return” exists beyond which treatment is unlikely to restore function. Baseline acuity, rate and duration of visual decline, optic atrophy, OCT thinning, tumor volume, chiasmatic involvement, and prior treatment may all influence whether radiation is followed by improvement, stability, or continued worsening. Future studies should stratify patients according to pretreatment visual trajectory and report outcomes at uniform post-treatment intervals.

The evolving molecular treatment landscape should also be incorporated into future research. MAPK pathway-directed therapies are increasingly used in pediatric low-grade glioma, and the role of radiation may shift toward salvage, consolidation, or selected durable-control scenarios. Outcomes should be examined according to molecular subtype, including BRAF-related alterations, whenever possible. Individual patient data meta-analysis would also be valuable because aggregate data cannot adequately adjust for baseline vision, age, NF1 status, tumor anatomy, molecular profile, dose, fractionation, prior chemotherapy, surgery, or follow-up duration. Ultimately, future OPHG trials and registries should treat preservation of useful vision as a primary endpoint rather than a secondary consequence of tumor control.

### Limitations

This study has several limitations. First, the evidence base was predominantly observational and retrospective, with inherent risks of selection bias, confounding by indication, incomplete follow-up, and incomplete outcome reporting. Patients selected for radiotherapy or radiosurgery may differ systematically from those managed with observation, chemotherapy, surgery, or targeted therapy, and these differences could not be fully addressed using aggregate study-level data.

Second, visual outcomes were not defined or measured uniformly across studies. Although the present review harmonized findings into preserved, improved, stable, and worsened categories, the original studies differed substantially in the visual tests performed, the timing of assessment, the thresholds used to define meaningful change, and whether outcomes were based on acuity, fields, global ophthalmological assessment, blindness status, or author-defined categories. This variability likely contributed materially to the observed clinical and statistical heterogeneity.

Third, only 19 of the 49 included studies could contribute to the quantitative visual synthesis. The remaining studies provided clinically relevant information but lacked sufficiently complete, compatible, or treatment-specific visual data for pooling. Consequently, the pooled estimates represent the subset of the literature with extractable categorical visual outcomes rather than the entire OPHG evidence base.

Fourth, outcome denominators sometimes represented patients, eyes, or other visual-outcome observations. Although patient-level outcomes were prioritized, combining different reporting units introduces potential unit-of-analysis heterogeneity and may violate independence when both eyes from the same patient are represented. Future studies should report patient-level and eye-level outcomes separately and account for within-patient correlation when eye-level analyses are performed.[11, 20, 22]

Fifth, radiation techniques, total doses, fractionation schedules, target definitions, treatment timing, and multimodal treatment sequences varied substantially across studies and treatment eras. Historical conventional radiotherapy cohorts are not directly comparable with modern conformal, proton, stereotactic, or radiosurgical approaches. Modality-specific and timing-based subgroup findings should therefore be regarded as exploratory.

Sixth, several subgroup analyses contained few studies, particularly those evaluating SRS/Gamma Knife, proton therapy, specific timing categories, and treatment-pure cohorts. Statistical significance in small subgroups may be unstable and susceptible to ecological bias, multiple testing, and confounding by study-level characteristics. These analyses should be considered hypothesis-generating rather than evidence of superiority or harm.

Seventh, one abstract-only or limited-report study contributed to the quantitative synthesis. Such reports provide less methodological detail than full-text publications and may carry greater uncertainty. However, restriction to full-text studies produced a similar preservation estimate, suggesting that this report did not materially alter the primary result.

Eighth, small-study effects and publication bias could not be excluded. Funnel-plot interpretation and Egger testing were exploratory because of the limited number of studies, substantial heterogeneity, and predominantly observational evidence base. Apparent asymmetry may therefore reflect differences in study size, treatment era, outcome definition, patient selection, or reporting quality rather than publication bias alone.

Finally, long-term toxicity was not the principal focus of this analysis. Endocrinopathy, vasculopathy, neurocognitive impairment, secondary malignancy, quality of life, and other late effects are essential to determining the overall therapeutic value of radiotherapy and radiosurgery in OPHG and must be considered alongside the possibility of visual preservation.

## Conclusion

Radiotherapy and radiosurgery were associated with preservation of visual function in approximately three quarters of evaluable visual outcomes in OPHG. The observed benefit appeared to be driven predominantly by visual stability rather than consistent recovery of established visual loss. A clinically meaningful proportion of outcomes nevertheless worsened, indicating that radiation-based treatment is not uniformly protective and should be individualized according to visual trajectory, tumor anatomy, prior treatment, age, NF1 status, and anticipated long-term toxicity.

These findings suggest that radiation-based treatment may be viewed primarily as a strategy for preserving remaining visual function rather than reliably restoring lost vision. Future OPHG research should prioritize standardized visual endpoints, prospective multicenter data collection, modern dosimetry, OCT and imaging biomarkers, molecular stratification, toxicity-adjusted outcomes, and individualized treatment timing. In a disease characterized by prolonged survival but potentially lifelong visual disability, preservation of useful vision should be considered a central therapeutic endpoint.

## Supporting information

Supplementary File 9, Panel A

Supplementary File 9, Panel B

Supplementary File 9, Panel C

Supplementary File 9, Panel D

Supplementary File 9, Panel E

Supplementary File 10, Panel A

Supplementary File 10, Panel B

Supplementary File 10, Panel C

Supplementary File 10, Panel D

Supplementary File 10, Panel E

Supplementary File 10, Panel F

## Declarations

## Acknowledgements

The authors have no additional acknowledgements to report.

## Ethics approval

Not applicable. This systematic review and meta-analysis was based exclusively on aggregate data obtained from previously published studies. No new human participants were recruited, no intervention was performed, no identifiable personal information was accessed, and no new individual-level participant data or biological material was collected. Therefore, approval from an institutional review board or research ethics committee was not required.

The review was prospectively registered in PROSPERO under registration number **CRD420261440136** and was conducted and reported in accordance with the PRISMA 2020 statement.

## Consent to participate

Not applicable. No human participants were recruited, contacted, or enrolled, and no new individual-level participant data were collected for this systematic review and meta-analysis.

## Consent for publication

Not applicable. This manuscript does not contain identifiable personal information, individual participant data, or identifiable clinical images.

## Data availability

All data supporting the findings of this systematic review and meta-analysis are included in the article and its supplementary materials. The complete database and supplementary search strategies are provided in **Supplementary File 3**. Title and abstract screening decisions are provided in **Supplementary File 4**, the final inclusion file and PICOS eligibility framework in **Supplementary File 5**, and the full-text exclusion decisions and corresponding reasons in **Supplementary File 6**.

The complete study-level data-extraction dataset is provided in **Supplementary File 7**, and the complete Joanna Briggs Institute risk-of-bias assessments are provided in **Supplementary File 8**. Supplementary visual-outcome forest plots, small-study-effect assessments, and leave-one-out analyses are provided in **Supplementary File 9**, while the complete subgroup and sensitivity analyses are provided in **Supplementary File 10**.

## Author contributions

**Farzan Fahim (FF)** conceived the study, contributed to the development of the research question and methodology, designed the standardized screening and data-extraction forms, coordinated the screening and extraction procedures, participated in data extraction and verification, contributed to the interpretation of the findings, supervised the preparation of the manuscript, and critically reviewed and revised the manuscript for important intellectual content.

**Amirmahdi Mojtahedzadeh (AMM)** conceived the study, developed the research question and methodology, contributed to preparation of the protocol, adjudicated disagreements during study selection, data extraction, and risk-of-bias assessment, verified the extracted data, performed the statistical analyses, prepared the figures and tables, interpreted the findings, and drafted the original manuscript.

**Vesal Abbasian (VA)** and **Mobina Puraminaie (MPA)** performed title and abstract screening according to the predefined eligibility criteria, contributed to the verification of screening decisions, and critically reviewed and revised the manuscript.

**Shima Khalili (SK)** and **Seyyed Mohammad Hosseini Marvast (SMHM)** performed full-text eligibility assessment, contributed to the verification of study inclusion and exclusion decisions, and critically reviewed and revised the manuscript.

**Zahra Zali (ZZ)** and **Zahra Darvishi (ZD)** independently performed the Joanna Briggs Institute risk-of-bias assessments, contributed to the resolution and verification of methodological assessments, and critically reviewed and revised the manuscript.

**Negin Aflaki (NA)**, **Parisa Fooladi (PF)**, **Parshana Farsandaj (PFa)**, **Mohammadreza AmaniTehrani (MAT)**, and **Farzin Mohammad Moradi (FMM)** contributed to verification and interpretation of the study-level findings and participated in the critical review, editing, and revision of the manuscript for important intellectual content.

**Nastaran Sadat Mahdavi (NSM)** contributed to study supervision, methodological and clinical interpretation of the findings, refinement of the manuscript, and critical review and revision of the manuscript for important intellectual content.

**Saeed Safari (SS)** contributed methodological and clinical supervision, interpretation of the findings, refinement of the study presentation, and critical review and revision of the manuscript for important intellectual content.

**Alireza Zali (AZ)** provided senior supervision, contributed neurosurgical and clinical expertise, assisted with interpretation of the findings and refinement of the clinical implications, and critically reviewed and revised the manuscript for important intellectual content.

FF and AMM contributed equally to this work and share first authorship. FF and NSM are co-corresponding authors. All authors contributed substantially to the intellectual development of the work, reviewed and approved the final version of the manuscript, agreed to its submission, and accept accountability for all aspects of the work.

## Supplementary File legends

Supplementary File 1. PRISMA 2020 checklist.

Completed PRISMA 2020 reporting checklist.

Supplementary File 2. Registered protocol.

Registered PROSPERO protocol describing the review rationale, eligibility criteria, methods, and planned analyses.

Supplementary File 3. Complete search strategies.

Complete search strategies for all electronic databases and supplementary information sources.

Supplementary File 4. Title and abstract exclusion sheet.

List of studies excluded during title and abstract screening with the corresponding reason for exclusion.

Supplementary File 5. Inclusion file and PICOS eligibility framework.

Final list of included studies together with the PICOS eligibility framework used for study selection.

Supplementary File 6. Full-text exclusion sheet.

Studies excluded following full-text assessment together with the corresponding reasons for exclusion.

Supplementary File 7. Complete data extraction sheet.

Complete data extraction sheet containing study, patient, tumor, treatment, outcome, and meta-analysis variables extracted from the included studies.

Supplementary File 8. Complete JBI risk-of-bias assessment.

Study-level Joanna Briggs Institute risk-of-bias assessments for all included studies.

Supplementary File 9. Supplementary visual outcome analyses.

Supplementary forest plots for visual improvement, visual stability, and visual worsening, together with publication-bias and leave-one-out analyses.

Supplementary File 10. Supplementary subgroup and sensitivity analyses.

Additional subgroup and sensitivity analyses of visual preservation according to treatment characteristics, study design, and risk of bias.

## Panel legends (Supplementary File 9)

Panel A. Visual improvement.

Forest plot of visual improvement following radiotherapy or radiosurgery. Panel B. Visual stability.

Forest plot of visual stability following radiotherapy or radiosurgery. Panel C. Visual worsening.

Forest plot of visual worsening following radiotherapy or radiosurgery. Panel D. Funnel plot.

Funnel plot evaluating potential small-study effects for visual preservation. Panel E. Leave-one-out analysis.

Leave-one-out sensitivity analysis of the primary visual preservation meta-analysis.

## Panel legends (Supplementary File 10)

Panel A. Visual preservation by radiation technique. Subgroup analysis according to radiation technique.

Panel B. Visual preservation by treatment timing.

Subgroup analysis according to radiotherapy/radiosurgery timing.

Panel C. Visual preservation by risk of bias.

Subgroup analysis according to Joanna Briggs Institute risk-of-bias category.

Panel D. Excluding high-risk-of-bias studies.

Sensitivity analysis excluding studies at high risk of bias.

Panel E. Full-text studies only.

Sensitivity analysis restricted to full-text publications.

Panel F. Pure RT/SRS cohorts.

Sensitivity analysis restricted to studies reporting outcomes from radiotherapy/radiosurgery-only cohorts.

