## Supplementary File 1 for "Visual Outcomes After Radiotherapy and Radiosurgery for Optic Pathway Hypothalamic Glioma: A Systematic Review and Meta-analysis"

| **Section and Topic** | **Item #** | **Checklist item** | **Location where item is reported** |
| --- | --- | --- | --- |
| **TITLE** | | |  |
| Title | 1 | Identify the report as a systematic review. |  |
| **ABSTRACT** | | |  |
| Abstract | 2 | See the PRISMA 2020 for Abstracts checklist. |  |
| **INTRODUCTION** | | |  |
| Rationale | 3 | Describe the rationale for the review in the context of existing knowledge. |  |
| Objectives | 4 | Provide an explicit statement of the objective(s) or question(s) the review addresses. |  |
| **METHODS** | | |  |
| Eligibility criteria | 5 | Specify the inclusion and exclusion criteria for the review and how studies were grouped for the syntheses. |  |
| Information sources | 6 | Specify all databases, registers, websites, organisations, reference lists and other sources searched or consulted to identify studies. Specify the date when each source was last searched or consulted. |  |
| Search strategy | 7 | Present the full search strategies for all databases, registers and websites, including any filters and limits used. |  |
| Selection process | 8 | Specify the methods used to decide whether a study met the inclusion criteria of the review, including how many reviewers screened each record and each report retrieved, whether they worked independently, and if applicable, details of automation tools used in the process. |  |
| Data collection process | 9 | Specify the methods used to collect data from reports, including how many reviewers collected data from each report, whether they worked independently, any processes for obtaining or confirming data from study investigators, and if applicable, details of automation tools used in the process. |  |
| Data items | 10a | List and define all outcomes for which data were sought. Specify whether all results that were compatible with each outcome domain in each study were sought (e.g. for all measures, time points, analyses), and if not, the methods used to decide which results to collect. |  |
|  | 10b | List and define all other variables for which data were sought (e.g. participant and intervention characteristics, funding sources). Describe any assumptions made about any missing or unclear information. |  |
| Study risk of bias assessment | 11 | Specify the methods used to assess risk of bias in the included studies, including details of the tool(s) used, how many reviewers assessed each study and whether they worked independently, and if applicable, details of automation tools used in the process. |  |
| Effect measures | 12 | Specify for each outcome the effect measure(s) (e.g. risk ratio, mean difference) used in the synthesis or presentation of results. |  |
| Synthesis methods | 13a | Describe the processes used to decide which studies were eligible for each synthesis (e.g. tabulating the study intervention characteristics and comparing against the planned groups for each synthesis (item #5)). |  |
|  | 13b | Describe any methods required to prepare the data for presentation or synthesis, such as handling of missing summary statistics, or data conversions. |  |
|  | 13c | Describe any methods used to tabulate or visually display results of individual studies and syntheses. |  |
|  | 13d | Describe any methods used to synthesize results and provide a rationale for the choice(s). If meta-analysis was performed, describe the model(s), method(s) to identify the presence and extent of statistical heterogeneity, and software package(s) used. |  |
|  | 13e | Describe any methods used to explore possible causes of heterogeneity among study results (e.g. subgroup analysis, meta-regression). |  |
|  | 13f | Describe any sensitivity analyses conducted to assess robustness of the synthesized results. |  |
| Reporting bias assessment | 14 | Describe any methods used to assess risk of bias due to missing results in a synthesis (arising from reporting biases). |  |
| Certainty assessment | 15 | Describe any methods used to assess certainty (or confidence) in the body of evidence for an outcome. |  |
| **RESULTS** | | |  |
| Study selection | 16a | Describe the results of the search and selection process, from the number of records identified in the search to the number of studies included in the review, ideally using a flow diagram. |  |
|  | 16b | Cite studies that might appear to meet the inclusion criteria, but which were excluded, and explain why they were excluded. |  |
| Study characteristics | 17 | Cite each included study and present its characteristics. |  |
| Risk of bias in studies | 18 | Present assessments of risk of bias for each included study. |  |
| Results of individual studies | 19 | For all outcomes, present, for each study: (a) summary statistics for each group (where appropriate) and (b) an effect estimate and its precision (e.g. confidence/credible interval), ideally using structured tables or plots. |  |
| Results of syntheses | 20a | For each synthesis, briefly summarise the characteristics and risk of bias among contributing studies. |  |
|  | 20b | Present results of all statistical syntheses conducted. If meta-analysis was done, present for each the summary estimate and its precision (e.g. confidence/credible interval) and measures of statistical heterogeneity. If comparing groups, describe the direction of the effect. |  |
|  | 20c | Present results of all investigations of possible causes of heterogeneity among study results. |  |
|  | 20d | Present results of all sensitivity analyses conducted to assess the robustness of the synthesized results. |  |
| Reporting biases | 21 | Present assessments of risk of bias due to missing results (arising from reporting biases) for each synthesis assessed. |  |
| Certainty of evidence | 22 | Present assessments of certainty (or confidence) in the body of evidence for each outcome assessed. |  |
| **DISCUSSION** | | |  |
| Discussion | 23a | Provide a general interpretation of the results in the context of other evidence. |  |
|  | 23b | Discuss any limitations of the evidence included in the review. |  |
|  | 23c | Discuss any limitations of the review processes used. |  |
|  | 23d | Discuss implications of the results for practice, policy, and future research. |  |
| **OTHER INFORMATION** | | |  |
| Registration and protocol | 24a | Provide registration information for the review, including register name and registration number, or state that the review was not registered. |  |
|  | 24b | Indicate where the review protocol can be accessed, or state that a protocol was not prepared. |  |
|  | 24c | Describe and explain any amendments to information provided at registration or in the protocol. |  |
| Support | 25 | Describe sources of financial or non-financial support for the review, and the role of the funders or sponsors in the review. |  |
| Competing interests | 26 | Declare any competing interests of review authors. |  |
| Availability of data, code and other materials | 27 | Report which of the following are publicly available and where they can be found: template data collection forms; data extracted from included studies; data used for all analyses; analytic code; any other materials used in the review. |  |

*From:*  Page MJ, McKenzie JE, Bossuyt PM, Boutron I, Hoffmann TC, Mulrow CD, et al. The PRISMA 2020 statement: an updated guideline for reporting systematic reviews. BMJ 2021;372:n71. doi: 10.1136/bmj.n71. This work is licensed under CC BY 4.0. To view a copy of this license, visit <https://creativecommons.org/licenses/by/4.0/>

| Section/topic | Item # | Location where item is reported in manuscript |
| --- | --- | --- |
| Title | 1 | Title page: “Visual Outcomes After Radiotherapy and Radiosurgery for Optic Pathway Hypothalamic Glioma: A Systematic Review and Meta-analysis.” |
| Abstract | 2 | Structured Abstract: Background, Objective, Methods, Results, and Conclusion. |
| Introduction: Rationale | 3 | Introduction, paragraphs 1–3: clinical characteristics and morbidity of OPHG; current management and role of RT/SRS; heterogeneity of the available visual-outcome evidence and need for a focused quantitative synthesis. |
| Introduction: Objectives | 4 | Introduction, final paragraph: objective to quantify visual outcomes after RT/SRS; visual preservation defined as the primary outcome, with visual improvement, stability, and worsening as secondary outcomes. |
| Methods: Eligibility criteria | 5 | Methods – Eligibility criteria: PICOS framework; eligible populations, radiation interventions, comparator requirements, visual outcomes, study designs, and exclusion criteria; separate eligibility requirements for the systematic review and quantitative visual synthesis. |
| Methods: Information sources | 6 | Methods – Search strategy and information sources: PubMed/MEDLINE, Scopus, Web of Science, Embase, and the Cochrane Library searched from inception to 1 June 2026; supplementary searches performed in Google Scholar and ClinicalTrials.gov. |
| Methods: Search strategy | 7 | Methods – Search strategy and information sources and Supplementary File 3: principal concepts and search terms described; complete PubMed/MEDLINE syntax presented in the manuscript; complete strategies for all databases and supplementary sources provided in Supplementary File 3. |
| Methods: Selection process | 8 | Methods – Study selection and screening: records imported into EndNote 2025 and deduplicated; title and abstract screening performed by VA and MPA; full-text assessment independently performed by SK and MH; disagreements or uncertainties resolved through discussion and, when required, adjudication by AMM. |
| Methods: Data collection process | 9 | Methods – Data extraction: standardized extraction sheet designed by FF; two trained reviewers independently extracted data; disagreements and uncertainties resolved through discussion with AMM. |
| Methods: Data items — outcomes | 10a | Methods – Eligibility criteria and Data extraction: primary outcome was visual preservation, defined as stable or improved vision; secondary outcomes were visual improvement, visual stability, and visual worsening; eligible visual assessment methods and quantitative outcome requirements were specified. |
| Methods: Data items — other variables | 10b | Methods – Data extraction: study identification, bibliographic information, country, centers, design, study period, population characteristics, age, sex, NF1 status, histology, tumor anatomy, baseline manifestations, previous and concomitant treatments, radiation modality, target, dose, fractionation, timing, indication, follow-up, radiological outcomes, endocrine outcomes, progression, survival, adverse events, and meta-analysis eligibility. |
| Methods: Study risk-of-bias assessment | 11 | Methods – Risk-of-bias assessment: appropriate Joanna Briggs Institute tool selected according to study design; assessments independently performed by ZZ and ZD; items judged as yes, no, unclear, or not applicable; scoring thresholds for low, moderate, and high risk of bias reported; disagreements adjudicated by AMM; complete assessments provided in Supplementary File 8. |
| Methods: Effect measures | 12 | Methods – Data synthesis and quantitative analysis: single-arm study-level proportions; pooled proportions reported with 95% confidence intervals and prediction intervals when estimable. |
| Methods: Synthesis methods — eligibility for synthesis | 13a | Methods – Eligibility criteria, Data extraction, and Data synthesis and quantitative analysis: quantitative inclusion required extractable improved, stable, or worsened visual outcomes with a corresponding denominator and sufficiently separable RT/SRS-associated data. |
| Methods: Synthesis methods — data preparation | 13b | Methods – Data extraction: visual preservation calculated as the sum of improved and stable outcomes when reported separately; patient-level outcomes prioritized; eye-level or other observation-level data retained when patient-level data were unavailable; ophthalmologist-assessed or author-defined global visual outcomes prioritized when multiple measures were reported. |
| Methods: Synthesis methods — tabulation and graphical display | 13c | Methods – Data synthesis and quantitative analysis and Subgroup, sensitivity, and small-study-effect analyses: study-level visual-outcome distribution plot, forest plots, funnel plot, leave-one-out analyses, subgroup analyses, sensitivity analyses, and summary Tables 3–4. |
| Methods: Synthesis methods — statistical model | 13d | Methods – Data synthesis and quantitative analysis: analyses performed in R version 4.6.0 using meta, metafor, readxl, dplyr, stringr, writexl, and ggplot2; random-effects generalized linear mixed models; logit transformation and back-transformation; heterogeneity evaluated using Cochran’s Q, I², and tau²; prediction intervals reported when estimable. |
| Methods: Synthesis methods — heterogeneity exploration | 13e | Methods – Subgroup, sensitivity, and small-study-effect analyses: subgroup analyses according to radiation technique, SRS/Gamma Knife, RT timing, study purity, and risk-of-bias category. |
| Methods: Synthesis methods — sensitivity analyses | 13f | Methods – Subgroup, sensitivity, and small-study-effect analyses: restriction to full-text reports, exclusion of high-risk-of-bias studies, restriction to pure RT/SRS cohorts, and leave-one-out analysis. |
| Methods: Reporting-bias assessment | 14 | Methods – Subgroup, sensitivity, and small-study-effect analyses: funnel plots used to explore small-study effects when sufficient studies were available; statistical testing for funnel-plot asymmetry and trim-and-fill analyses treated as exploratory because of the limited number of studies, observational designs, and clinical heterogeneity. |
| Methods: Certainty assessment | 15 | Not applicable. |
| Results: Study selection | 16a | Results – Study selection and Figure 1: 7,333 records identified; 6,529 duplicates removed; 804 records screened; 300 excluded during title and abstract screening; 504 full texts assessed; 455 excluded; 49 studies included in the systematic review; 19 studies with 494 evaluable visual-outcome observations included in the quantitative synthesis. |
| Results: Excluded studies | 16b | Results – Study selection, Supplementary File 4, and Supplementary File 6: title and abstract exclusion categories reported; complete title and abstract exclusion sheet provided in Supplementary File 4; full-text exclusion reasons and the complete full-text exclusion sheet provided in Supplementary File 6. |
| Results: Study characteristics | 17 | Results – Characteristics of the included studies; Patient and tumor characteristics; Radiotherapy, radiosurgery, and concomitant treatment characteristics; Tables 1 and 2: design, setting, population, age, sex, NF1 status, tumor anatomy, follow-up, treatment modality, dose, fractionation, and treatment timing summarized. |
| Results: Risk of bias in studies | 18 | Results – Risk-of-bias assessment, Figure 5, and Supplementary File 8: numbers of studies at low, moderate, and high risk of bias; common methodological limitations; study-level classifications for the quantitative synthesis; complete item-level assessments in Supplementary File 8. |
| Results: Results of individual studies | 19 | Results – Visual outcome assessment and quantitative eligibility; Figure 2; Figure 3; Supplementary File 9; and Supplementary File 10: study-level visual-outcome data, individual study estimates, forest plots, and subgroup displays. |
| Results: Results of syntheses — contributing studies | 20a | Results – Visual outcome assessment and quantitative eligibility; Primary outcome: visual preservation; Secondary visual outcomes; Tables 3 and 4: 19 contributing studies and 494 visual-outcome observations described, with contributing studies identified in the manuscript and supplementary analyses. |
| Results: Results of syntheses — pooled estimates | 20b | Results – Primary outcome: visual preservation; Secondary visual outcomes; Table 3; and Figure 3: event counts, denominators, pooled proportions, 95% confidence intervals, prediction intervals, I², tau², and Cochran’s Q p-values reported. |
| Results: Heterogeneity investigations | 20c | Results – Primary outcome: visual preservation; Secondary visual outcomes; Table 4; and Supplementary File 10: subgroup findings according to radiation technique, RT timing, study purity, and risk-of-bias category. |
| Results: Sensitivity analyses | 20d | Results – Primary outcome: visual preservation; Secondary visual outcomes; Table 4; Supplementary File 9; and Supplementary File 10: exclusion of high-risk-of-bias studies, full-text-only analyses, restriction to pure RT/SRS cohorts, and leave-one-out analyses. |
| Results: Reporting biases | 21 | Results – Small-study effects and publication-bias assessment and Supplementary File 9: funnel-plot appearance described; Egger-type regression result reported as intercept 2.29, p = 0.060; limitations of interpretation acknowledged. |
| Results: Certainty of evidence | 22 | Not applicable. |
| Discussion: General interpretation | 23a | Discussion – Principal findings; Biological and pathophysiological interpretation; Comparison with previous literature; and Radiation technique and subgroup findings: interpretation of visual preservation, improvement, stability, worsening, heterogeneity, and subgroup signals. |
| Discussion: Limitations of evidence | 23b | Discussion – Limitations: observational and retrospective evidence base; selection bias and confounding by indication; heterogeneous visual assessments and denominators; incomplete reporting; treatment-era differences; small subgroup sizes; abstract-only evidence; and possible small-study effects. |
| Discussion: Limitations of review processes | 23c | Discussion – Limitations: only 19 of 49 studies were quantitatively eligible; use of aggregate data; mixture of patient-, eye-, and observation-level denominators; inability to adjust fully for baseline vision, age, NF1 status, tumor anatomy, treatment sequence, dose, fractionation, and follow-up; exploratory publication-bias assessment. |
| Discussion: Implications | 23d | Discussion – Clinical implications; Visual preservation as a core endpoint; Future directions; and Conclusion: implications for clinical counseling, treatment selection, standardized visual outcomes, prospective registries, biomarkers, dosimetry, molecular stratification, and future research. |
| Other information: Registration | 24a | Methods – Protocol and reporting: PROSPERO registration number CRD420261440136. |
| Other information: Protocol access | 24b | Methods – Protocol and reporting and Supplementary File 2: registered protocol provided as Supplementary File 2. |
| Other information: Amendments | 24c | No amendments to the registered protocol. |
| Other information: Support | 25 | Declarations – Funding: “No funding was received for this study.” |
| Other information: Competing interests | 26 | Declarations – Conflicts of interest: “The authors declare that they have no conflicts of interest relevant to this work.” |
| Other information: Availability of data, code, and materials | 27 | Supplementary Files 3–10 and Declarations – Data availability. |
