## Supplementary File 2 for "Visual Outcomes After Radiotherapy and Radiosurgery for Optic Pathway Hypothalamic Glioma: A Systematic Review and Meta-analysis"

### Registered protocol

#### Title

#### Registration

This systematic review and meta-analysis was registered in PROSPERO under the registration number **CRD420261440136**.

#### Background and rationale

Optic pathway hypothalamic glioma (OPHG) is a rare glioma involving the optic nerve, optic chiasm, optic tract, hypothalamus, or adjacent visual pathway structures. OPHG occurs predominantly in children and may be sporadic or associated with neurofibromatosis type 1 (NF1). Although some tumors remain indolent, others progress and cause substantial long-term morbidity, particularly visual impairment, endocrine dysfunction, hypothalamic injury, neurodevelopmental consequences, and reduced quality of life.

Visual dysfunction is often the most clinically important outcome in OPHG. Patients may develop reduced visual acuity, visual field loss, optic atrophy, strabismus, nystagmus, proptosis, or blindness. Because survival is often prolonged, preservation of useful vision is a central therapeutic goal.

Management of OPHG is individualized according to age, NF1 status, tumor location, symptoms, radiological progression, visual trajectory, prior treatment, and expected treatment-related toxicity. Observation may be appropriate for stable disease, while chemotherapy and targeted therapies are commonly used for progressive pediatric low-grade glioma. Surgery is generally reserved for biopsy, decompression, cyst management, or selected exophytic components because of the risk of visual and hypothalamic injury. Radiotherapy and radiosurgery remain important options for progressive, recurrent, or treatment-refractory disease when durable tumor control and visual stabilization are needed.

Despite decades of experience with radiotherapy and radiosurgery in OPHG, the expected visual benefit remains incompletely defined. Existing studies vary in radiation technique, treatment timing, dose and fractionation, patient selection, baseline visual status, follow-up duration, and visual outcome definitions. Many reports also combine radiotherapy with surgery, chemotherapy, or other treatments, making treatment-specific interpretation difficult. Previous reviews have generally emphasized tumor control, survival, endocrine outcomes, or broad treatment strategies rather than quantitatively separating visual preservation, improvement, stability, and worsening after radiation-based treatment.

Therefore, this systematic review and meta-analysis was designed to quantify visual outcomes after radiotherapy and radiosurgery for OPHG, with particular emphasis on visual preservation.

### Objectives

The primary objective was to estimate the pooled proportion of patients or visual-outcome observations with **visual preservation** after radiotherapy or radiosurgery for OPHG.

The secondary objectives were to estimate the pooled proportions of:

1. **Visual improvement**
2. **Visual stability**
3. **Visual worsening or deterioration**

Additional objectives were to explore whether visual outcomes differed according to:

1. Radiation technique
2. Stereotactic radiosurgery/Gamma Knife use
3. Timing of radiotherapy
4. Study purity, defined as pure RT/SRS cohorts versus RT-containing mixed-treatment cohorts
5. Risk-of-bias category

### Review question

Among patients with optic pathway hypothalamic glioma or related optic pathway/hypothalamic gliomas treated with radiotherapy or radiosurgery, what proportion experience visual preservation, visual improvement, visual stability, or visual worsening?

### PICO framework

#### Population

Eligible populations included patients of any age with OPHG or related gliomas involving the optic nerve, optic chiasm, optic tract, optic pathway, visual pathway, chiasmatic-hypothalamic region, or hypothalamic region.

Eligible disease terms included:

- Optic pathway hypothalamic glioma
- Optic pathway glioma
- Optic nerve glioma
- Optic chiasm glioma
- Chiasmatic glioma
- Chiasmatic-hypothalamic glioma

- Optochiasmatic glioma
- Visual pathway glioma
- Optic pathway/hypothalamic glioma
- Hypothalamic glioma
- Hypothalamic low-grade glioma

Pediatric and adult populations were eligible. Both NF1-associated and sporadic tumors were eligible.

#### Intervention

Eligible interventions included radiotherapy or radiosurgery, including:

- Conventional external-beam radiotherapy
- Historical photon radiotherapy
- Local-field radiotherapy
- 3D conformal radiotherapy
- Intensity-modulated radiotherapy
- Proton therapy
- Fractionated stereotactic radiotherapy
- Stereotactic radiosurgery
- Gamma Knife radiosurgery
- CyberKnife radiosurgery
- Other radiation-based approaches

Studies were eligible for the systematic review if they reported relevant data on radiotherapy or radiosurgery in OPHG. Studies were eligible for quantitative visual outcome synthesis if they reported extractable visual outcomes after radiotherapy or radiosurgery, or if visual outcomes for a clearly identifiable RT/SRS subgroup could be extracted.

#### Comparator

No comparator was required because the primary analysis was a single-arm meta-analysis of proportions after radiotherapy or radiosurgery. Comparative studies were eligible if the RT/SRS group or subgroup was extractable separately.

#### Outcomes

The primary outcome was **visual preservation**, defined as stable or improved vision after radiotherapy or radiosurgery.

Secondary outcomes were:

- **Visual improvement**
- **Visual stability**
- **Visual worsening or deterioration**

Visual outcomes could be based on visual acuity, visual field assessment, ophthalmological examination, blindness status, optic pathway functional assessment, or author-defined categorical visual outcome.

For quantitative synthesis, studies were required to report the number of patients, eyes, or visual-outcome observations with improved, stable, or worsened vision, together with the corresponding denominator.

### Eligibility criteria

#### Inclusion criteria

Studies were eligible if they met all of the following criteria:

1. Included patients with OPHG or related optic pathway/hypothalamic glioma.
2. Reported radiotherapy or radiosurgery as part of treatment.
3. Reported clinical, radiological, visual, endocrine, survival, progression, or adverse-event outcomes relevant to OPHG.
4. Were original human studies.
5. Had extractable OPHG-related data.
6. For quantitative visual outcome synthesis, reported extractable visual outcome counts after RT/SRS or for a clearly identifiable RT/SRS subgroup.

Eligible study designs included:

- Randomized studies
- Non-randomized interventional studies
- Prospective cohort studies
- Retrospective cohort studies
- Single-arm observational studies
- Case series with extractable outcome data
- Conference abstracts if extractable relevant data were available

#### Exclusion criteria

Studies were excluded if they met any of the following criteria:

1. Did not include OPHG or related optic pathway/hypothalamic glioma.
2. Included non-glioma optic pathway tumors only.
3. Did not involve radiotherapy or radiosurgery.
4. Did not report extractable OPHG or RT/SRS-related outcomes.
5. Did not provide extractable visual outcome data for quantitative synthesis, where quantitative inclusion was being considered.
6. Were reviews, editorials, commentaries, letters without original patient data, or guidelines.
7. Were preclinical, animal, or laboratory-only studies.

8. Were duplicate reports of the same cohort without additional extractable data.
9. Were case reports or very small case series without extractable outcome data.
10. Were conference abstracts without sufficient extractable information.

Non-English studies were not excluded a priori. Potentially eligible non-English studies were planned for translation using artificial intelligence-assisted translation with manual verification.

### Information sources

The following databases were searched from inception to **1 June 2026**:

1. PubMed/MEDLINE
2. Scopus
3. Web of Science
4. Embase
5. Cochrane

No restrictions were applied by publication year or language during the database search.

### Search strategy

The search strategy combined controlled vocabulary and free-text terms related to OPHG and radiation-based treatment.

Disease-related terms included:

- optic pathway glioma
- optic nerve glioma
- hypothalamic glioma
- chiasmatic glioma
- chiasmal glioma
- optic pathway hypothalamic glioma
- optic pathway/hypothalamic glioma
- OPHG

Intervention-related terms included:

- radiotherapy
- radiation therapy
- irradiation
- external beam radiotherapy
- proton therapy
- stereotactic radiosurgery
- radiosurgery
- Gamma Knife

- IMRT
- fractionated radiotherapy

The PubMed/MEDLINE search syntax was:

(("Optic Nerve Glioma"[Mesh]) OR ("optic pathway glioma*"[Title/Abstract]) OR ("optic nerve glioma*"[Title/Abstract]) OR ("hypothalamic glioma*"[Title/Abstract]) OR ("chiasmatic glioma*"[Title/Abstract]) OR (OPHG[Title/Abstract])) AND (("Radiotherapy"[Mesh]) OR ("radiation therap*"[Title/Abstract]) OR (radiotherap*[Title/Abstract]) OR ("proton therap*"[Title/Abstract]) OR ("stereotactic radiosurgery"[Title/Abstract]) OR (IMRT[Title/Abstract]))

The search syntax was adapted for each database, including MeSH terms in PubMed/MEDLINE and Emtree terms in Embase. The complete search strategies for all databases were provided in Supplementary File 3.

### Study selection process

All retrieved records were imported into EndNote 2025, and duplicates were removed before screening. After deduplication, records were exported into standardized Excel screening forms designed by the senior author.

Title and abstract screening was performed by two reviewers. Records were assessed according to predefined eligibility criteria. Clearly irrelevant records were excluded, while unclear or potentially eligible records were retained for full-text review. Disagreements or uncertainties were resolved by a third reviewer.

Full-text screening was performed using standardized Excel forms. Each report was categorized as included, excluded, or requiring discussion. Reasons for exclusion were recorded using standardized exclusion categories. Disagreements were resolved by discussion and, when needed, adjudication by a third reviewer.

The study selection process was summarized using a PRISMA 2020 flow diagram.

### Data extraction

Data extraction was performed using a standardized extraction sheet. Extracted variables included:

- Study identification
- First author
- Publication year
- Study title
- Country

- Study center
- Study design
- Study period
- Inclusion criteria
- Exclusion criteria
- Total study population
- Eligible OPHG population
- RT/SRS subgroup size
- Age
- Sex
- Pediatric/adult status
- NF1 status
- Histology
- Tumor location
- Baseline visual symptoms
- Baseline endocrine features
- Baseline neurological features
- Prior treatment
- Prior surgery
- Prior chemotherapy
- Radiotherapy or radiosurgery modality
- Radiation dose
- Fractionation
- Timing of RT/SRS
- Treatment indication
- Follow-up duration
- Radiological outcomes
- Visual outcomes
- Endocrine outcomes
- Progression outcomes
- Survival outcomes
- Adverse events
- Risk-of-bias data
- Meta-analysis eligibility

For visual outcome synthesis, the following variables were extracted:

- Visual outcome denominator
- Number with improved vision
- Number with stable vision
- Number with worsened vision
- Number with preserved vision

Visual preservation was calculated as improved plus stable vision when these categories were reported separately.

When multiple visual outcome measures were available, ophthalmological or author-defined global visual outcome was prioritized for the main analysis. Visual acuity-specific and visual field-specific outcomes were extracted when available.

Missing or unclear information was recorded as not reported.

### Risk-of-bias assessment

Risk of bias was assessed using the appropriate Joanna Briggs Institute critical appraisal tool according to study design.

The following JBI tools were planned:

- JBI checklist for randomized controlled trials
- JBI checklist for cohort studies
- JBI checklist for quasi-experimental studies
- JBI checklist for case series

Each applicable item was judged as:

- Yes
- No
- Unclear
- Not applicable

For each study, a score was calculated as the number of "yes" responses divided by the number of applicable items. Studies were categorized as low, moderate, or high risk of bias according to the proportion of applicable criteria fulfilled.

Item-level risk-of-bias judgments were summarized in a traffic-light figure, and the complete assessment was provided as Supplementary File 8.

### Data synthesis and statistical analysis

The primary quantitative synthesis was a single-arm meta-analysis of proportions. The primary pooled outcome was visual preservation after radiotherapy or radiosurgery. Secondary pooled outcomes were visual improvement, visual stability, and visual worsening.

For each outcome, the event count and corresponding visual-outcome denominator were extracted from each study.

All analyses were planned in R using the following packages:

- meta
- metafor

- readxl
- dplyr
- stringr
- writexl
- ggplot2

Pooled proportions were estimated using random-effects models because clinical and methodological heterogeneity was expected. Sources of heterogeneity included differences in:

- Age distribution
- NF1 status
- Tumor location
- Baseline visual impairment
- Prior surgery
- Prior chemotherapy
- Radiation technique
- Dose and fractionation
- Treatment timing
- Treatment indication
- Follow-up duration
- Visual outcome definition

The main meta-analysis used logit transformation of proportions with a generalized linear mixed model. The logit-transformed proportion was defined as:

$$\text{logit}(p) = \log[p / (1 - p)]$$

where p represents the study-level visual outcome proportion.

Results were reported as pooled proportions with 95% confidence intervals. Prediction intervals were reported when appropriate.

Statistical heterogeneity was assessed using:

- $I^2$
- $tau^2$
- Cochran's Q
- prediction intervals

Forest plots were generated for:

- Visual preservation
- Visual improvement
- Visual stability
- Visual worsening

Funnel plots were planned to assess small-study effects when at least 10 studies were available for an outcome. Funnel plot interpretation was considered exploratory because of the rare-disease setting, observational study designs, small study sizes, and clinical heterogeneity.

### Planned subgroup analyses

Predefined subgroup analyses were planned to explore potential sources of heterogeneity.

#### Radiation technique

Radiation technique subgroups included:

- Historical conventional photon radiotherapy
- 3D conformal photon radiotherapy
- Fractionated stereotactic radiotherapy
- Stereotactic radiosurgery/Gamma Knife
- Mixed or unclear radiotherapy approach

A specific SRS/Gamma Knife subgroup was planned because radiosurgery differs from conventional fractionated radiotherapy in dose delivery, fractionation, target volume, and patient selection.

#### Radiotherapy timing

Radiotherapy timing subgroups included:

- Primary or upfront radiotherapy
- Salvage radiotherapy
- Adjuvant radiotherapy
- Radiotherapy after surgery
- Mixed or unclear radiotherapy timing

#### Study purity

Study-purity subgroups included:

- Pure RT/SRS cohorts
- RT-containing mixed-treatment cohorts
- Abstract or limited-data studies

Pure RT/SRS cohorts were defined as studies in which visual outcomes were clearly attributable to radiotherapy or radiosurgery cohorts without major mixing with other treatment groups.

#### Risk-of-bias category

Risk-of-bias subgroup analyses compared:

- Low risk of bias
- Moderate risk of bias
- High risk of bias

### Planned sensitivity analyses

Sensitivity analyses were planned to assess robustness of the primary findings. These included:

1. Exclusion of high-risk-of-bias studies
2. Restriction to full-text studies
3. Restriction to pure RT/SRS cohorts
4. Leave-one-out analysis

Leave-one-out analysis repeated the primary meta-analysis after excluding one study at a time to assess whether the pooled estimate was driven by any individual study.

### Reporting bias assessment

Small-study effects and potential reporting bias were assessed using funnel plots when at least 10 studies were available for an outcome. These analyses were considered exploratory because asymmetry in rare-disease meta-analyses may reflect clinical heterogeneity, selective reporting, small-study effects, differences in treatment era, or methodological variation rather than publication bias alone.

### Certainty assessment

A formal certainty-of-evidence assessment using GRADE was not performed. This was because the available evidence was expected to consist mainly of observational, retrospective, heterogeneous studies with variable visual outcome definitions and limited comparability across treatment eras and radiation techniques.

### Presentation of results

The study selection process was planned for presentation using a PRISMA 2020 flow diagram.

Included studies were summarized in tables describing:

- Study characteristics
- Patient, tumor, and treatment characteristics
- Visual outcome meta-analysis results
- Subgroup and sensitivity analyses

Figures were planned to include:

- PRISMA flow diagram
- Study-level distribution of visual outcomes
- Forest plot of visual preservation
- Subgroup analysis of visual preservation by study purity
- Risk-of-bias traffic-light plot

Supplementary materials were planned to include:

- PRISMA checklist
- Registered protocol
- Complete search strategies
- Screening and eligibility workflow forms
- Inclusion sheet
- Full exclusion table
- Complete data extraction sheet
- Complete risk-of-bias assessment
- Supplementary visual outcome forest plots and small-study-effect analyses

### Ethics

Ethics approval was not required because this study was a systematic review and meta-analysis of previously published aggregate data and did not involve new recruitment of human participants or use of individual patient-level identifiable data.

### Funding

No funding was received for this study.

### Conflicts of interest

The authors declare that they have no conflicts of interest relevant to this work.
