## Supplementary File 4 for "Visual Outcomes After Radiotherapy and Radiosurgery for Optic Pathway Hypothalamic Glioma: A Systematic Review and Meta-analysis"

PubMed

(
("Optic Nerve Glioma"[Mesh])
OR
("optic pathway glioma*"[Title/Abstract])
OR
("optic nerve glioma*"[Title/Abstract])
OR
("hypothalamic glioma*"[Title/Abstract])
OR
("chiasmatic glioma*"[Title/Abstract])
OR
(OPHG[Title/Abstract])
)
AND
(
("Radiotherapy"[Mesh])
OR
("radiation therap*"[Title/Abstract])
OR
(radiotherap*[Title/Abstract])
OR
("proton therap*"[Title/Abstract])
OR
("stereotactic radiosurgery"[Title/Abstract])
OR
(IMRT[Title/Abstract])
)

SCOPUS

(
("optic pathway glioma*"
OR "optic nerve glioma*"
OR "hypothalamic glioma*"
OR "chiasmatic glioma*"
OR OPHG)
AND
("radiotherapy"
OR "radiation therapy"
OR radiotherap*
OR "proton therapy"
OR "stereotactic radiosurgery"

OR IMRT)
)

WOS

(
("optic pathway glioma*"
OR "optic nerve glioma*"
OR "hypothalamic glioma*"
OR "chiasmatic glioma*"
OR OPHG)
AND
("radiotherapy"
OR "radiation therapy"
OR radiotherap*
OR "proton therapy"
OR "stereotactic radiosurgery"
OR IMRT)
)

Embase

(
'exp optic nerve glioma'/exp
OR
'optic pathway glioma*':ti,ab
OR
'optic nerve glioma*':ti,ab
OR
'hypothalamic glioma*':ti,ab
OR
'chiasmatic glioma*':ti,ab
OR
'OPHG':ti,ab
)
AND
(
'exp radiotherapy'/exp
OR
radiotherap*:ti,ab
OR
'radiation therapy':ti,ab
OR
'proton therapy':ti,ab

OR
'stereotactic radiosurgery':ti,ab
OR
IMRT:ti,ab
)

Cochrane

(

"optic pathway glioma"

OR "optic pathway gliomas"

OR "optic nerve glioma"

OR "optic nerve gliomas"

OR "hypothalamic glioma"

OR "hypothalamic gliomas"

OR "chiasmatic glioma"

OR "chiasmatic gliomas"

OR "chiasmal glioma"

OR "chiasmal gliomas"

OR "optic pathway hypothalamic glioma"

OR "optic pathway/hypothalamic glioma"

OR OPHG

)

AND

(

radiotherap*

OR "radiation therapy"

OR irradiation

OR "external beam radiotherapy"

OR "external beam radiation therapy"

OR "proton therapy"

OR "proton beam therapy"

OR radiosurgery

OR "stereotactic radiosurgery"

OR IMRT

OR "intensity modulated radiotherapy"

OR "fractionated radiotherapy"

)

Google Scholar

("optic pathway glioma" OR "optic pathway hypothalamic glioma" OR "optic chiasm glioma" OR "hypothalamic glioma") AND ("visual acuity" OR "visual outcome" OR "visual function" OR "vision preservation" OR blindness OR ophthalmology) AND (outcome OR prognosis OR follow-up OR treatment OR therapy)

Clinical Trials.gov

"optic pathway glioma" OR "optic pathway hypothalamic glioma" OR "optic glioma" OR "chiasmatic glioma" OR "hypothalamic glioma" | Other terms: vision OR visual OR "visual acuity" OR "visual field" OR "visual function" OR ophthalmologic OR blindness
