## Supplementary figures and images for "Visual Outcomes After Radiotherapy and Radiosurgery for Optic Pathway Hypothalamic Glioma: A Systematic Review and Meta-analysis"

### Supplementary File 9, Panel A

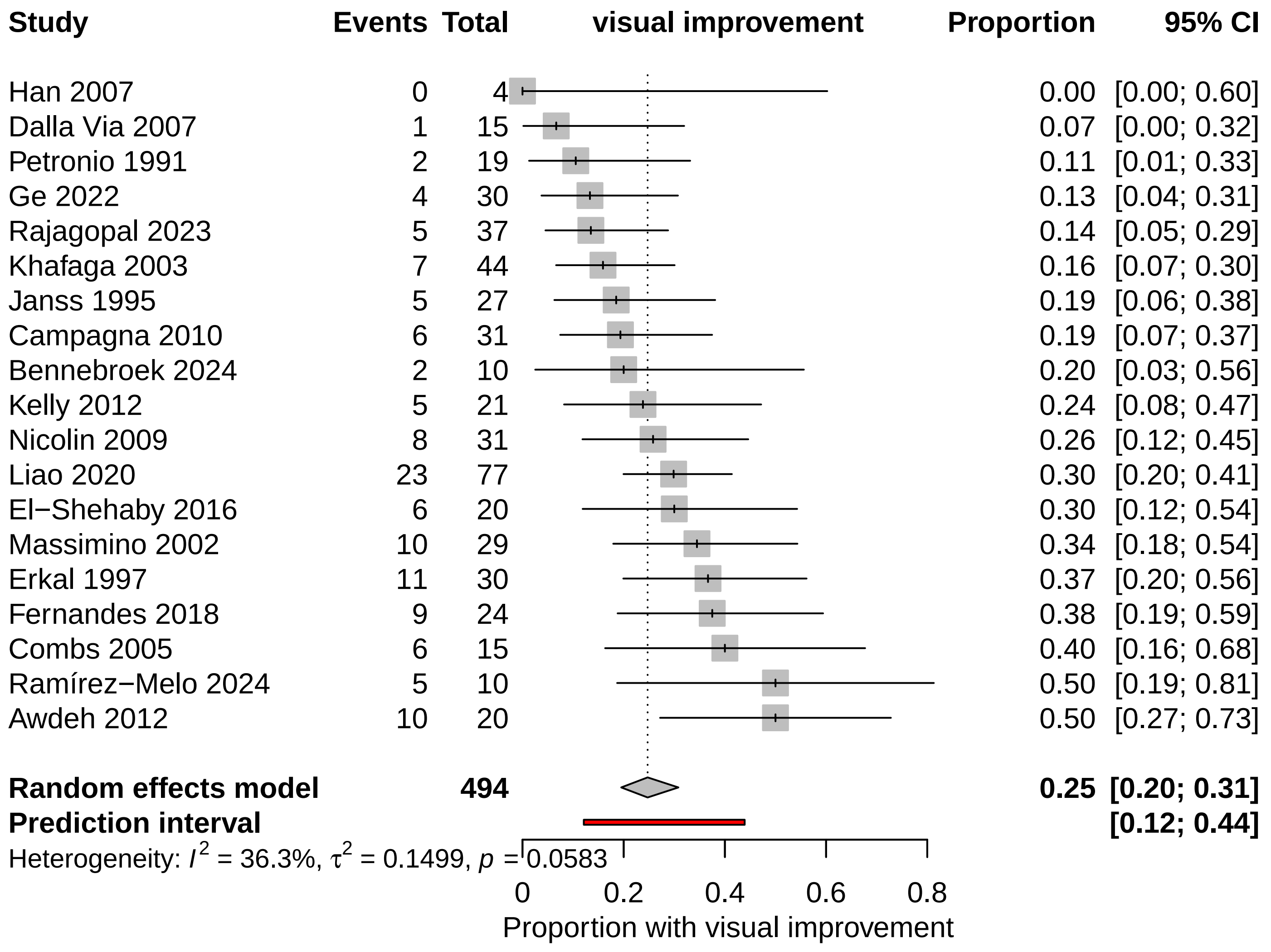

### Supplementary File 9, Panel B

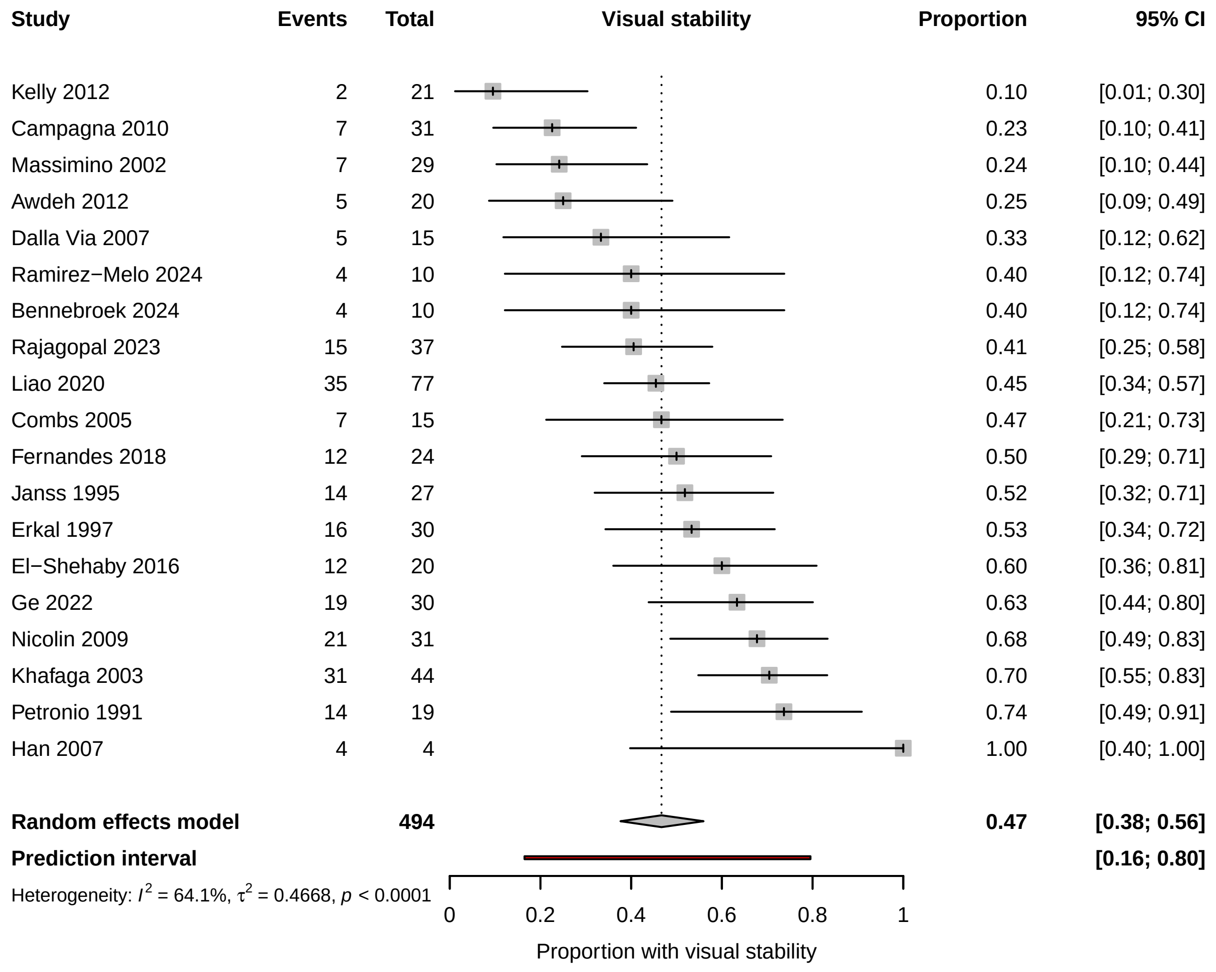

### Supplementary File 9, Panel C

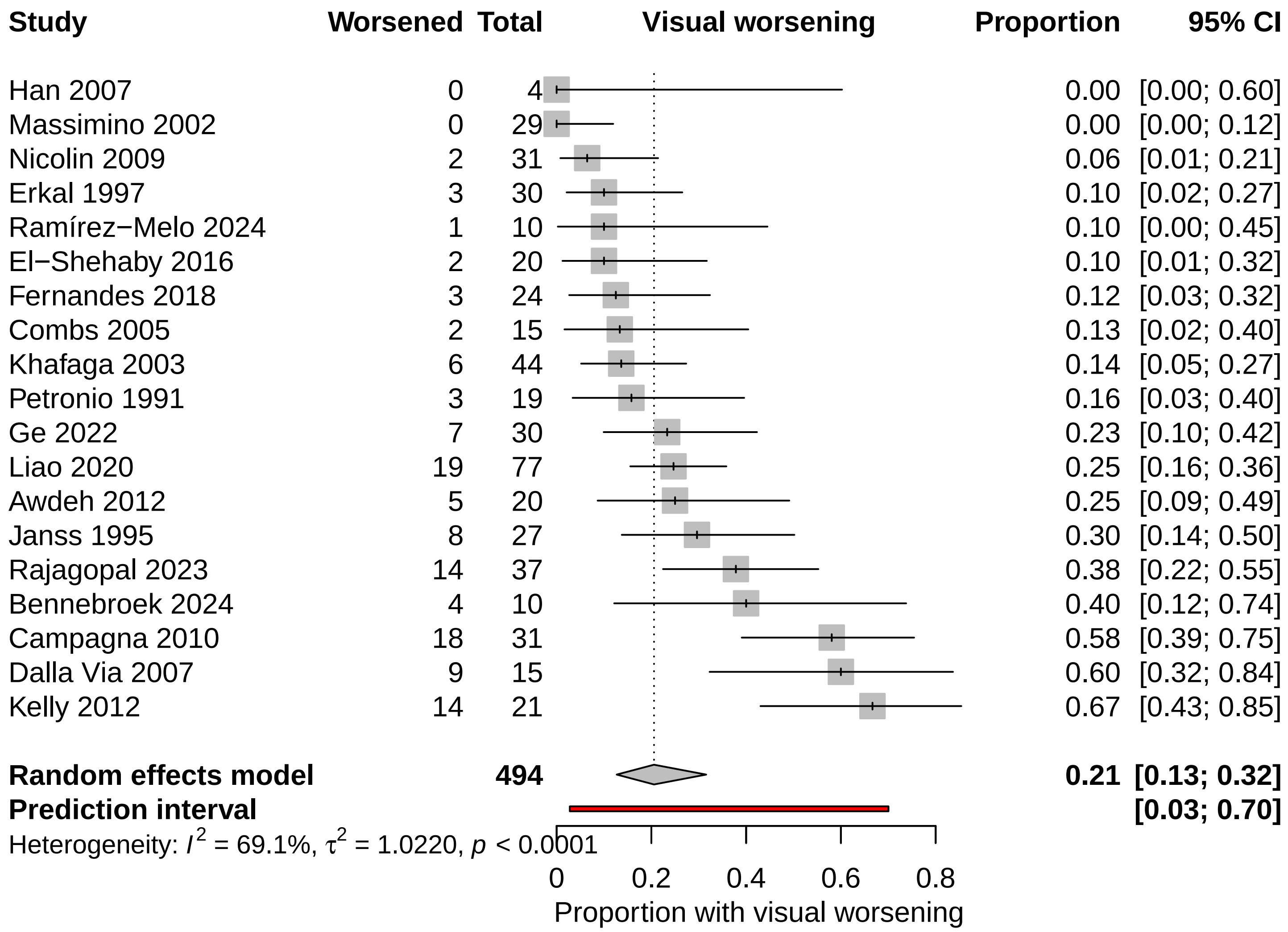

### Supplementary File 9, Panel D

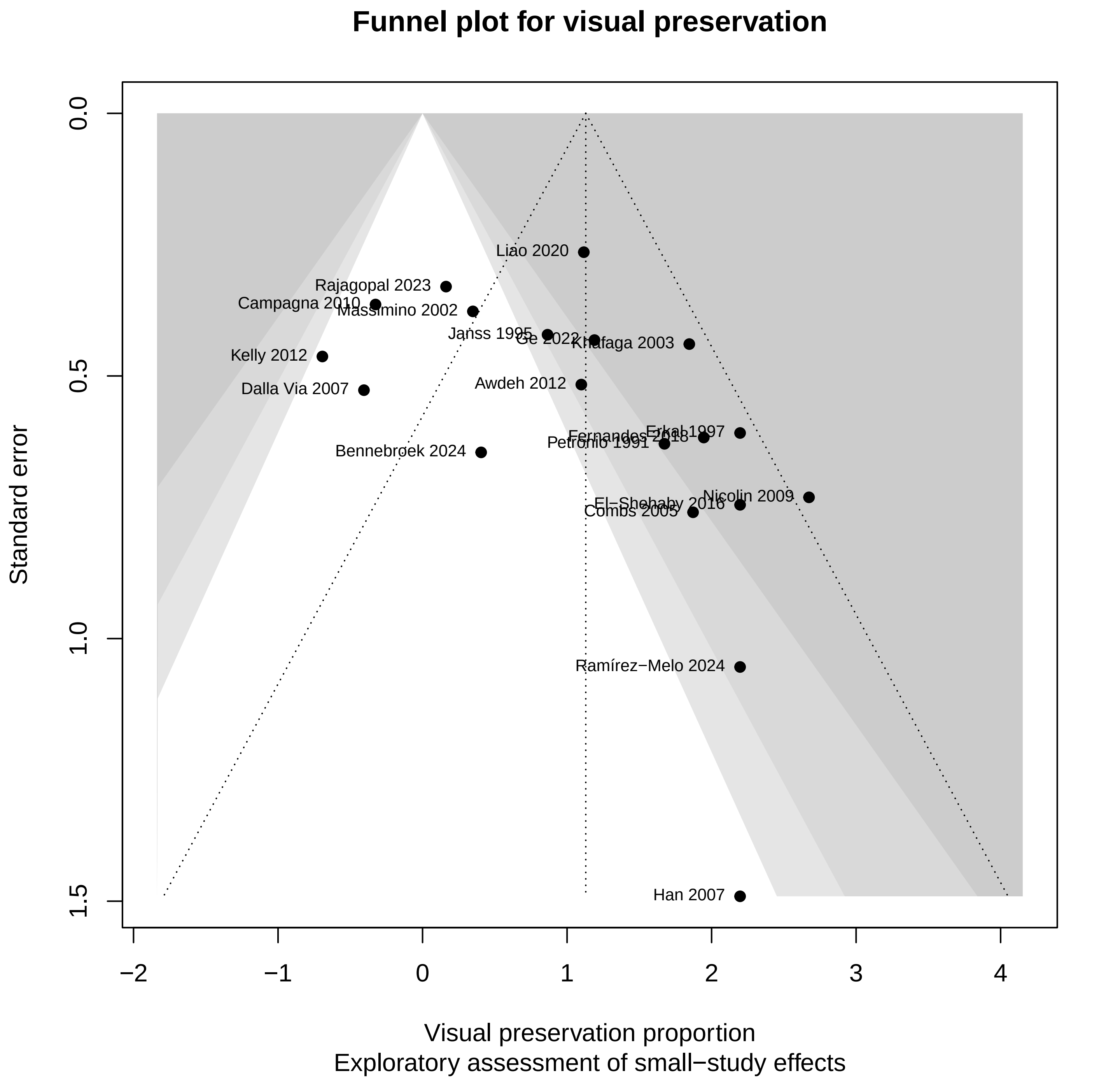

### Supplementary File 10, Panel E

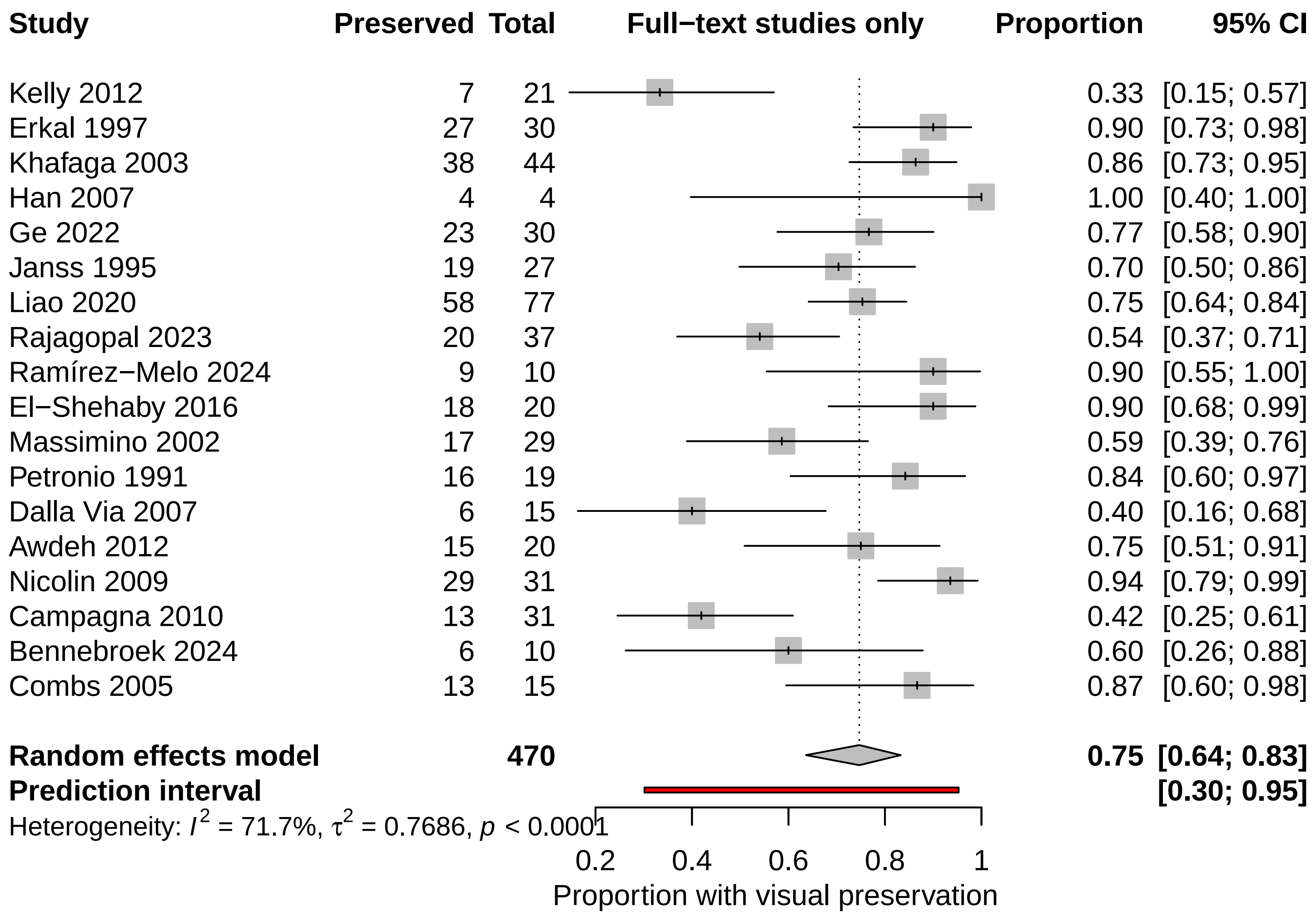

### Supplementary File 10, Panel F

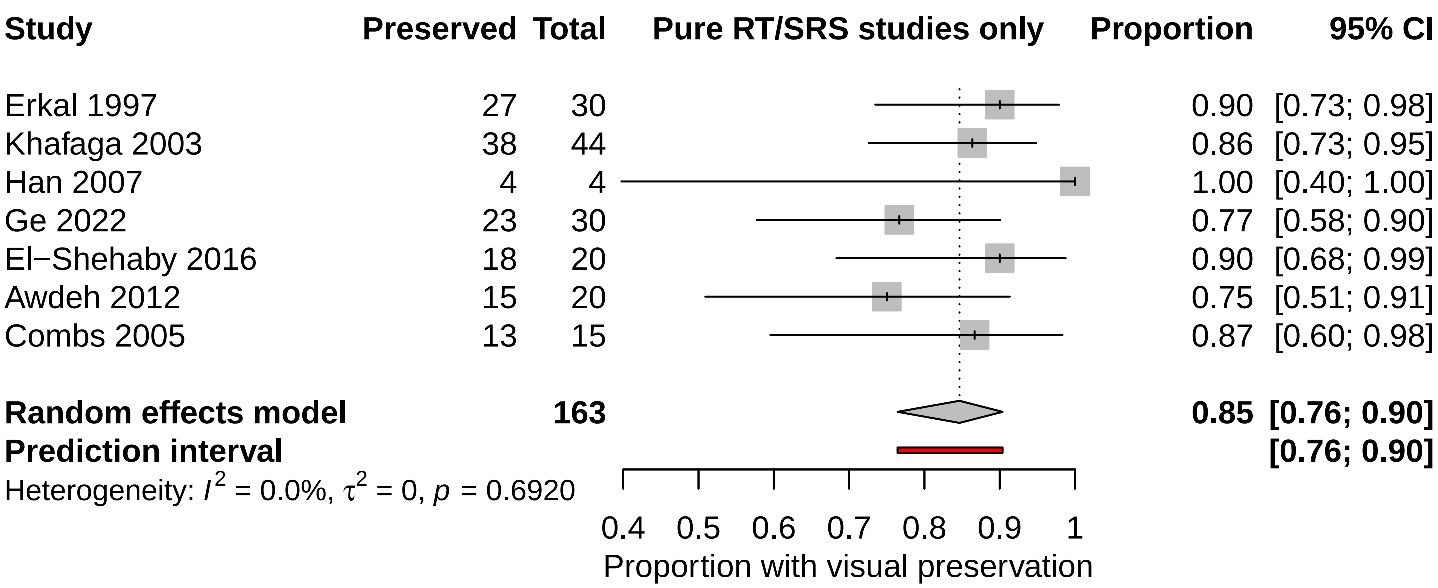
