## Supplementary File 9, Panel E for "Visual Outcomes After Radiotherapy and Radiosurgery for Optic Pathway Hypothalamic Glioma: A Systematic Review and Meta-analysis"

| Study | Proportion | 95% CI/PI | Tau2 | Tau | I2 |
|---|---|---|---|---|---|
| Omitting Kelly 2012 | 0.77 | [0.68; 0.85]<br>[0.38; 0.95] | 0.6075 | 0.7794 | 67.4% |
| Omitting Erkal 1997 | 0.74 | [0.63; 0.83]<br>[0.31; 0.95] | 0.7332 | 0.8563 | 70.8% |
| Omitting Khafaga 2003 | 0.75 | [0.64; 0.83]<br>[0.30; 0.95] | 0.7774 | 0.8817 | 70.5% |
| Omitting Han 2007 | 0.75 | [0.64; 0.83]<br>[0.31; 0.95] | 0.7471 | 0.8643 | 73.2% |
| Omitting Ge 2022 | 0.76 | [0.64; 0.84]<br>[0.29; 0.96] | 0.8348 | 0.9137 | 72.9% |
| Omitting Janss 1995 | 0.76 | [0.65; 0.84]<br>[0.30; 0.96] | 0.8367 | 0.9147 | 73.2% |
| Omitting Liao 2020 | 0.76 | [0.64; 0.84]<br>[0.29; 0.96] | 0.8473 | 0.9205 | 72.6% |
| Omitting Rajagopal 2023 | 0.77 | [0.66; 0.85]<br>[0.32; 0.96] | 0.7724 | 0.8789 | 71.2% |
| Omitting Ramírez−Melo 2024 | 0.75 | [0.64; 0.83]<br>[0.30; 0.95] | 0.7624 | 0.8732 | 72.5% |
| Omitting El−Shehaby 2016 | 0.74 | [0.64; 0.83]<br>[0.31; 0.95] | 0.7452 | 0.8633 | 71.7% |
| Omitting Massimino 2002 | 0.77 | [0.66; 0.85]<br>[0.31; 0.96] | 0.8016 | 0.8953 | 72.5% |
| Omitting Petronio 1991 | 0.75 | [0.64; 0.84]<br>[0.30; 0.96] | 0.7958 | 0.8921 | 72.4% |
| Omitting Dalla Via 2007 | 0.77 | [0.67; 0.85]<br>[0.35; 0.95] | 0.6985 | 0.8358 | 70.6% |
| Omitting Awdeh 2012 | 0.76 | [0.64; 0.84]<br>[0.30; 0.96] | 0.8308 | 0.9115 | 73.1% |
| Omitting Fernandes 2018 | 0.75 | [0.64; 0.83]<br>[0.30; 0.95] | 0.7686 | 0.8767 | 71.7% |
| Omitting Nicolin 2009 | 0.74 | [0.63; 0.82]<br>[0.32; 0.94] | 0.6689 | 0.8179 | 70.1% |
| Omitting Campagna 2010 | 0.77 | [0.67; 0.85]<br>[0.36; 0.95] | 0.6715 | 0.8195 | 67.6% |
| Omitting Bennebroek 2024 | 0.76 | [0.65; 0.85]<br>[0.31; 0.96] | 0.8061 | 0.8978 | 73.0% |
| Omitting Combs 2005 | 0.75 | [0.64; 0.83]<br>[0.30; 0.95] | 0.7792 | 0.8827 | 72.4% |
| **Random effects model** | **0.76** | **[0.65; 0.84]** | **0.7654** | **0.8749** | **71.6%** |
| **Prediction interval** | | **[0.31; 0.95]** | | | |

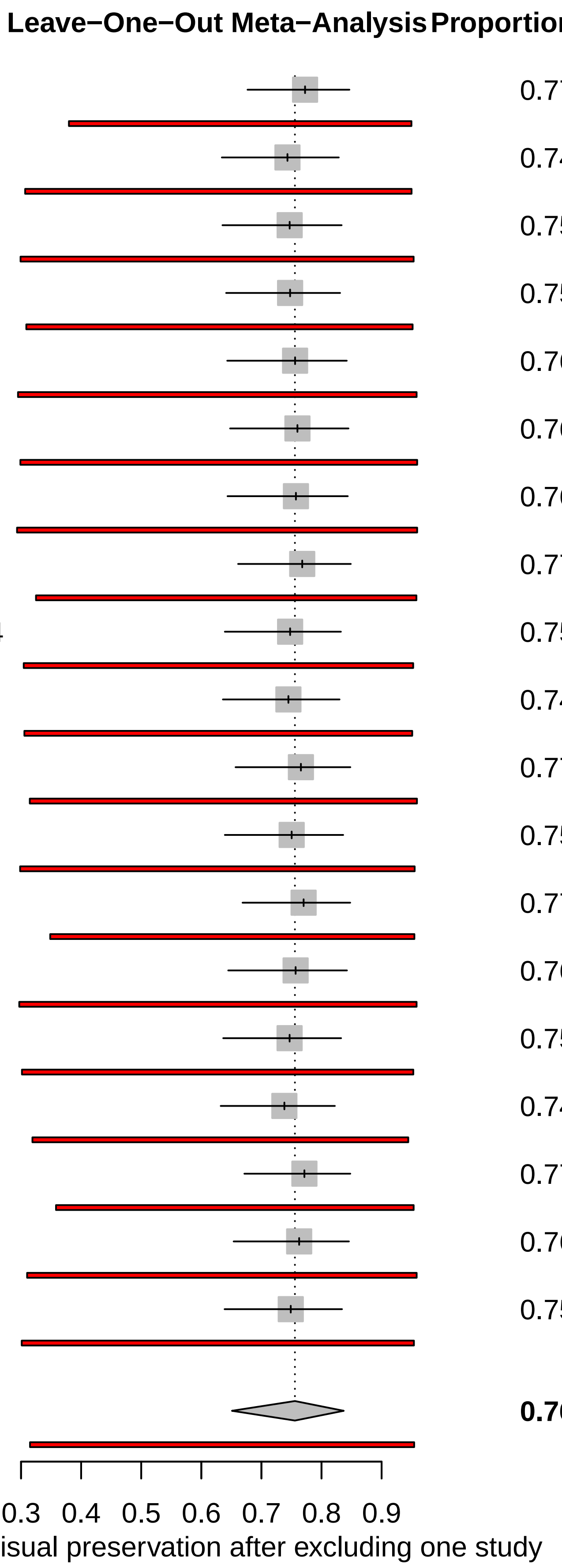
