## Supplementary File 10, Panel B for "Visual Outcomes After Radiotherapy and Radiosurgery for Optic Pathway Hypothalamic Glioma: A Systematic Review and Meta-analysis"

| Study | Preserved | Total | Subgroup by RT timing | Proportion | 95% CI |
|---|---|---|---|---|---|
| **Salvage RT** | | | | | |
| Kelly 2012 | 7 | 21 | | 0.33 | [0.15; 0.57] |
| Erkal 1997 | 27 | 30 | | 0.90 | [0.73; 0.98] |
| Han 2007 | 4 | 4 | | 1.00 | [0.40; 1.00] |
| Massimino 2002 | 17 | 29 | | 0.59 | [0.39; 0.76] |
| Petronio 1991 | 16 | 19 | | 0.84 | [0.60; 0.97] |
| Dalla Via 2007 | 6 | 15 | | 0.40 | [0.16; 0.68] |
| Nicolin 2009 | 29 | 31 | | 0.94 | [0.79; 0.99] |
| Combs 2005 | 13 | 15 | | 0.87 | [0.60; 0.98] |
| **Random effects model** | | **164** | | **0.77** | **[0.56; 0.90]** |
| Heterogeneity: $I^2$ = 78.4%, $\tau^2$ = 1.4492, $p$ < 0.0001 | | | | | |
| **Primary / upfront RT** | | | | | |
| Khafaga 2003 | 38 | 44 | | 0.86 | [0.73; 0.95] |
| Ge 2022 | 23 | 30 | | 0.77 | [0.58; 0.90] |
| Janss 1995 | 19 | 27 | | 0.70 | [0.50; 0.86] |
| **Random effects model** | | **101** | | **0.79** | **[0.70; 0.86]** |
| Heterogeneity: $I^2$ = 25.1%, $\tau^2$ = < 0.0001, $p$ = 0.2629 | | | | | |
| **RT after surgery** | | | | | |
| Rajagopal 2023 | 20 | 37 | | 0.54 | [0.37; 0.71] |
| **Adjuvant RT** | | | | | |
| Ramirez−Melo 2024 | 9 | 10 | | 0.90 | [0.55; 1.00] |
| **Mixed / unclear RT timing** | | | | | |
| Liao 2020 | 58 | 77 | | 0.75 | [0.64; 0.84] |
| El−Shehaby 2016 | 18 | 20 | | 0.90 | [0.68; 0.99] |
| Awdeh 2012 | 15 | 20 | | 0.75 | [0.51; 0.91] |
| Fernandes 2018 | 21 | 24 | | 0.88 | [0.68; 0.97] |
| Campagna 2010 | 13 | 31 | | 0.42 | [0.25; 0.61] |
| Bennebroek 2024 | 6 | 10 | | 0.60 | [0.26; 0.88] |
| **Random effects model** | | **182** | | **0.74** | **[0.58; 0.85]** |
| Heterogeneity: $I^2$ = 73.1%, $\tau^2$ = 0.5154, $p$ = 0.0023 | | | | | |
| **Random effects model** | | **494** | | **0.76** | **[0.66; 0.83]** |
| **Prediction interval** | | | | | **[0.31; 0.95]** |
| Heterogeneity: $I^2$ = 71.6%, $\tau^2$ = 0.7654, $p$ < 0.0001 | | | | | |
| Test for subgroup differences: $\chi^2_4$ = 9.86, df = 4 ($p$ = 0.0429) | | | | | |

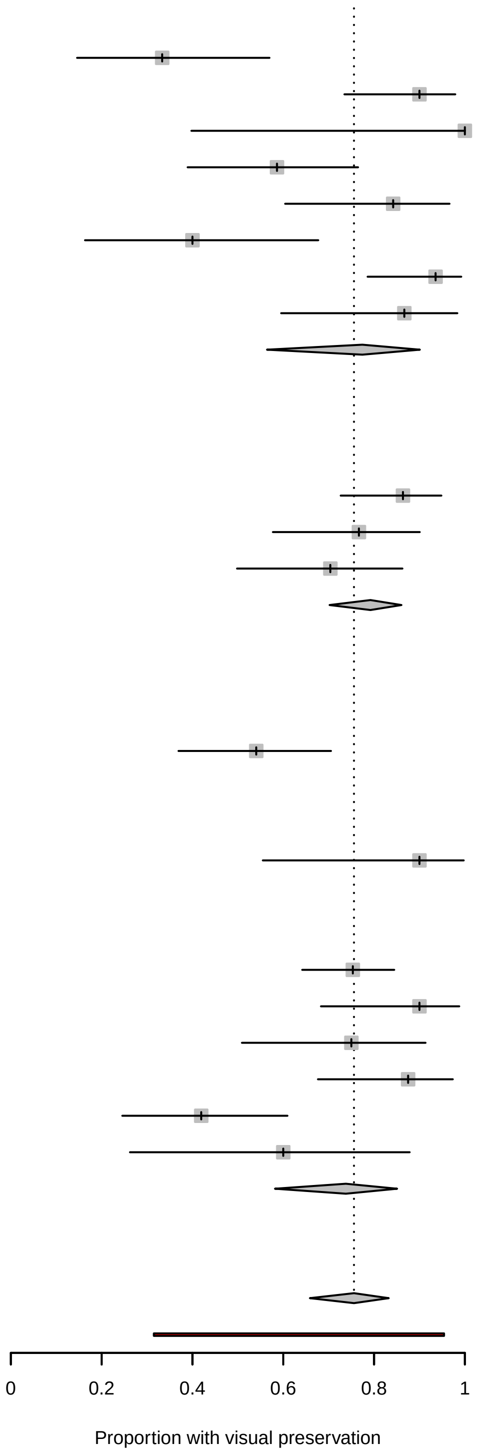
