## Supplementary material for "Visual Outcomes After Radiotherapy and Radiosurgery for Optic Pathway Hypothalamic Glioma: A Systematic Review and Meta-analysis": Tables 1,2,3,4

**Table 1. Study characteristics of included studies.**

| First_Author | Year | Study_Title | Country | Study_Design | Study_Period | Eligible_OPHG_N | RT_SRS_Subgroup_N | Age_Years | NF1_Status | Tumor_Location | Prior_Treatment_or_Management | RT_SRS_Modality | Dose_Fractionation | Follow_Up | Reported_Outcomes |
| --- | --- | --- | --- | --- | --- | --- | --- | --- | --- | --- | --- | --- | --- | --- | --- |
| Kovalic | 1990 | Radiation Therapy for Gliomas of the Optic Nerve and Chiasm | USA | Retrospective radiotherapy cohort / long-term outcome study | 1956-1986 | 33 | 33 | median 4; range 1-34 | NF1 11; sporadic 22; unknown 0 | Optic glioma cohort: 5 optic nerve only, 8 optic nerve plus chiasm, and 20 optic nerve plus chiasm plus contiguous structures; most had chiasmal or extrachiasmal extension | All analyzed patients received local-field radiation therapy; 22 had surgery before RT including biopsy or partial excision, and 11 received irradiation alone | Local-field radiation therapy using orthovoltage, cobalt teletherapy or megavoltage photon therapy | total 50.4 median; per fraction 0.76-2.0; median 1.6; range 15.22-61.0 including one incomplete course; otherwise 37.86-61.0; median 50.4 | median mo 147.6; range mo 24-372 among survivors | Visual; Endocrine; Progression; Survival; Adverse events |
| Kelly | 2012 | Longitudinal Measures of Visual Function, Tumor Volume, and Prediction of Visual Outcomes after Treatment of Optic Pathway Gliomas | USA | Retrospective longitudinal visual-function and tumor-volume cohort | July 1988-June 2007 | 21 | 11 | range 0.7-9 at treatment | NF1 6; sporadic 15; unknown 0 | Bilateral OPG cohort involving chiasm alone or with anterior/posterior extension; Table 1 locations included chiasm in most patients, optic nerve involvement in selected patients, hypothalamus in 6, optic tracts in 6 and visual radiations in 2 | Initial treatment was chemotherapy in 18 and external-beam radiotherapy in 3; radiotherapy was reserved for older non-NF1 children or after at least one failed chemotherapy regimen | External-beam radiation therapy | NR | mean mo 108; range mo 48-192 | Visual; Radiological; Progression; Survival |
| Erkal | 1997 | Management of optic pathway and chiasmatic-hypothalamic gliomas in children with radiation therapy | Turkey | Retrospective pediatric radiotherapy cohort | January 1973-October 1994 | 33 | 33 | median 7; range 2-18 | NF1 2; sporadic 31; unknown 0 | Optic pathway/chiasmatic-hypothalamic glioma cohort; optic nerve 2, chiasm 6, chiasm plus optic nerve 7, chiasm plus hypothalamus 11, chiasm plus hypothalamus plus optic nerve 7 | Radiotherapy for all patients; prior surgery in 14 including biopsy, subtotal resection or decompressive surgery; chemotherapy in 2 | External-beam radiotherapy using cobalt-60 or 6-MV photons | total 45-55; per fraction 1.5-2; range 45-55; median 50 | mean mo 163.2; range mo 6-193.2 | Visual; Radiological; Endocrine; Progression; Survival; Adverse events |
| Khafaga | 2003 | Optic Gliomas: A Retrospective Analysis of 50 Cases | Saudi Arabia | Retrospective optic pathway low-grade glioma cohort | January 1980-December 1995 | 50 | 28 initial RT; 35 any RT over course | median 4; range 2-36 | NF1 5; sporadic 45; unknown 0 | Optic pathway low-grade glioma cohort; optic nerve only 6, chiasm 3, optic nerve plus chiasm 15, chiasm plus hypothalamus 23, chiasm plus optic tract 3 | Initial management included surgery, radiotherapy, observation, and rare chemotherapy; 28 had initial RT and 35 received RT at some point | External-beam radiotherapy; photon technique not otherwise specified | total 45-54; per fraction 1.8; range 45-54; median 50.4 | median mo 84; range mo 28.8-198 | Visual; Radiological; Endocrine; Progression; Survival; Adverse events |
| Tsang | 2017 | Clinical outcomes in children with optic pathway glioma treated with radiation therapy | Canada | Retrospective radiotherapy cohort | 1986-2015 | 89 | 89 | median 7.9 at RT start; range 1.1-19.8 at RT start | NF1 19; sporadic 70; unknown 0 | Pediatric OPG treated with RT; optic chiasm involved in 71, hypothalamus in 64, optic nerve in 41, optic tract in 30, optic radiation in 11, and other intracranial extension in selected patients | RT used after observation, chemotherapy, surgery, or as part of multimodal management; 47 had prior chemotherapy, 39 prior surgery, and 7 prior RT | External-beam radiotherapy; photon/proton and conformal techniques across eras | total 50.4-54; per fraction 1.8; range 45-59.4; median 54 | median mo 105.6; range mo 5.4-351.6 | Visual; Radiological; Endocrine; Progression; Survival; Adverse events |
| Segal | 2010 | Optic pathway gliomas in patients with neurofibromatosis type 1: Follow-up of 9 years | USA | Retrospective NF1-associated OPG cohort | 1990-2007 | 42 | 3 | median 6 at OPG diagnosis | NF1 42; sporadic 0; unknown 0 | NF1-associated OPG; tumor location included optic nerve, chiasm, optic tract and hypothalamic involvement according to clinical and imaging records | Most patients observed; treatment included chemotherapy, surgery and rare radiotherapy for progressive disease | Radiotherapy; technique not specified | NR | mean mo 108 | Visual; Radiological; Progression |
| Liu | 2018 | Management of optic pathway gliomas in children: A 9-year single-center experience | China | Retrospective pediatric OPG management cohort | 2008-2016 | 37 | 4 | mean 6.7; range 0.75-17 | NF1 0; sporadic 37; unknown 0 | Pediatric OPG cohort; tumors involved optic nerve, chiasm, hypothalamus and optic tract in variable combinations; Dodge classification reported in study | Management included observation, surgery, chemotherapy and radiotherapy according to symptoms, age, tumor location and progression | Radiotherapy; technique not specified | NR | median mo 37; range mo 6-107 | Visual; Radiological; Endocrine; Progression; Survival |
| Han | 2007 | Optic pathway glioma treated with fractionated stereotactic radiotherapy | China | Retrospective fractionated stereotactic radiotherapy cohort | 1999-2005 | 14 | 14 | mean 12 at Novalis RS; 10.2 at initial diagnosis; range 5-16 at Novalis RS; 4-13 at initial diagnosis | NR | Optic pathway gliomas involving optic nerve, chiasm, optic tract or hypothalamus; detailed location distribution not fully extractable | Fractionated stereotactic radiotherapy for all patients; most had prior surgery and/or chemotherapy before radiation | Fractionated stereotactic radiotherapy with Novalis system | total 45-50; per fraction 1.8-2; range 45-50; median 50 | median mo 34; range mo 16-72 | Visual; Radiological; Progression; Survival; Adverse events |
| Mahoney | 2000 | Visual outcomes in children with optic pathway gliomas | USA | Retrospective visual outcome cohort | 1980-1997 | 42 | 10 | median 1.96; range 0.33-5.5 | NF1 26; sporadic 16; unknown 0 | Pediatric OPG cohort involving optic nerve, chiasm, tract, hypothalamus or radiations; anatomic subgroups reported by Dodge classification | Management included observation, chemotherapy, surgery and radiotherapy according to disease progression and visual status | Radiotherapy; technique not specified | NR | median mo 59; range mo 6-193 | Visual; Progression |
| Packer | 1988 | Treatment of chiasmatic/hypothalamic gliomas of childhood with chemotherapy | USA | Prospective chemotherapy cohort / clinical trial experience | 1977-1986 | 24 | 4 | median 2.4; range 0.25-15 | NF1 5; sporadic 19; unknown 0 | Chiasmatic/hypothalamic glioma cohort; tumors involved optic chiasm, hypothalamus and adjacent optic pathway structures | Primary chemotherapy strategy used to delay or avoid radiotherapy; radiotherapy used for progression in selected patients | Conventional external-beam radiotherapy | NR | median mo 51; range mo 3-120 | Visual; Radiological; Progression; Survival; Adverse events |
| Janss | 1995 | Optic pathway and hypothalamic/chiasmatic gliomas in children younger than age 5 years with a 6-year follow-up | Country NR | Retrospective pediatric cohort / treatment outcome study | NR | 44 | 12 | median 1.8; range 0.2-4.9 | NF1 12; sporadic 32; unknown 0 | Optic pathway and hypothalamic/chiasmatic gliomas in children younger than 5 years; detailed location included optic nerve, chiasm, hypothalamus and posterior pathway involvement | Management included chemotherapy, surgery, radiotherapy and combinations; RT generally delayed or used for progression | External-beam radiotherapy; technique not specified | NR | median mo 72; range mo 6-168 | Visual; Radiological; Endocrine; Progression; Survival; Adverse events |
| Fouladi | 2003 | Survival and functional outcome of children with hypothalamic/chiasmatic tumors treated on the Pediatric Oncology Group protocol | USA | Prospective Pediatric Oncology Group protocol cohort | 1986-1994 | 91 | NR | median 3.2; range 0.1-20.1 | NR | Hypothalamic/chiasmatic tumors including optic pathway gliomas; tumors involved optic chiasm, hypothalamus and adjacent structures | Protocol-based management with surgery, chemotherapy and/or radiotherapy according to age, diagnosis and protocol assignment | Radiotherapy; technique not specified | NR | median mo 60 | Visual; Endocrine; Progression; Survival; Adverse events |
| Liao | 2020 | The Visual Acuity Outcome and Relevant Factors Affecting Visual Improvement in Pediatric Sporadic Chiasmatic-Hypothalamic Glioma Patients Who Received Surgery | China | Retrospective surgical/prognostic cohort | 2007-2017 | 45 | 15 | median 7.7; range 0.9-18.8 | NF1 0; sporadic 45; unknown 0 | Sporadic pediatric chiasmatic-hypothalamic glioma; tumors involved chiasm and hypothalamic region; included prechiasmatic, chiasmatic and postchiasmatic extension | All patients received surgery; postoperative treatment included radiotherapy, chemotherapy, combined therapy or observation | Radiotherapy after surgery; technique not specified | NR | median mo 35; range mo 3-120 | Visual; Radiological; Endocrine; Progression; Survival; Adverse events |
| Hanania | 2021 | Long-term outcomes of pediatric optic pathway glioma | USA | Retrospective pediatric OPG cohort | 1990-2018 | 83 | 18 | median 4.1; range 0.3-17.8 | NF1 37; sporadic 46; unknown 0 | OPG cohort involving optic nerve, chiasm, tract, hypothalamus and posterior visual pathway in variable combinations | Treatment included observation, chemotherapy, surgery, radiation and targeted therapy according to progression, symptoms and era | Radiotherapy; technique not specified | NR | median mo 96; range mo 6-300 | Visual; Radiological; Endocrine; Progression; Survival; Adverse events |
| Green | 2022 | Bevacizumab for pediatric low-grade glioma: A national service evaluation | United Kingdom | Conference abstract / nationwide service evaluation | 2009-2020 | 88% OPG; exact count NR | 0 | NR | NF1 24% NF1; exact count NR | Progressive pediatric low-grade glioma cohort, predominantly optic pathway glioma | Bevacizumab-based therapy for progressive disease; radiotherapy not used in extracted subgroup | NA | NR | NR | Visual; Radiological; Progression; Survival; Adverse events |
| Ge | 2022 | Clinical outcomes of stereotactic radiosurgery for optic pathway gliomas | China | Retrospective Gamma Knife radiosurgery cohort | 2007-2018 | 20 | 20 | median 11; range 4-35 | NR | Optic pathway gliomas treated with Gamma Knife; lesions involved optic nerve, optic chiasm, optic tract or hypothalamic region | All patients received Gamma Knife radiosurgery; prior surgery or other treatment variably reported | Gamma Knife stereotactic radiosurgery | total 8-14; per fraction 8-14; fractions 1; range 8-14; median 11.5 | median mo 43; range mo 12-104 | Visual; Radiological; Endocrine; Progression; Survival; Adverse events |
| Lim | 2022 | Clinical features and treatment outcomes of pediatric optic pathway glioma | South Korea | Retrospective pediatric OPG cohort | 2000-2020 | 57 | 9 | median 4.8; range 0.2-17.9 | NF1 32; sporadic 25; unknown 0 | Pediatric OPG involving optic nerve, optic chiasm, optic tract, hypothalamus and posterior visual pathway according to imaging | Management included observation, chemotherapy, surgery and radiotherapy according to progression and symptoms | Radiotherapy; technique not specified | NR | median mo 78; range mo 6-221 | Visual; Radiological; Endocrine; Progression; Survival |
| Lohkamp | 2022 | Optic pathway gliomas: Long-term visual and endocrine outcome | Germany | Retrospective long-term OPG outcome cohort | 1990-2018 | 60 | 14 | median 4.6; range 0.1-17.2 | NF1 28; sporadic 32; unknown 0 | OPG cohort involving optic nerve, chiasm, hypothalamus and optic tract/posterior pathway structures | Treatment included observation, chemotherapy, surgery, radiotherapy and targeted therapy according to clinical course | Radiotherapy; technique not specified | NR | median mo 108; range mo 12-312 | Visual; Endocrine; Progression; Survival; Adverse events |
| Picariello | 2022 | Visual and endocrine outcomes in children with optic pathway glioma | United Kingdom | Retrospective pediatric OPG outcome cohort | 1995-2019 | 102 | 25 | median 4.3; range 0.2-16.4 | NF1 47; sporadic 55; unknown 0 | Pediatric OPG cohort with optic nerve, chiasmatic, hypothalamic and posterior pathway involvement | Management included observation, chemotherapy, surgery, radiotherapy and targeted therapy based on symptoms and progression | Radiotherapy; technique not specified | NR | median mo 84; range mo 12-240 | Visual; Endocrine; Progression; Survival; Adverse events |
| Rajagopal | 2023 | Visual outcomes in pediatric optic pathway glioma after treatment | USA | Retrospective visual outcome cohort | 2000-2020 | 59 | 13 | median 5.2; range 0.4-18 | NF1 29; sporadic 30; unknown 0 | Pediatric OPG cohort with optic nerve, chiasm, hypothalamus and posterior visual pathway involvement | Treatment included observation, chemotherapy, surgery, radiotherapy and targeted therapy | Radiotherapy; technique not specified | NR | median mo 72; range mo 6-216 | Visual; Radiological; Progression |
| Bennebroek | 2024 | Treatment of isolated pediatric optic nerve glioma: A nationwide retrospective cohort study and systematic literature review on visual and radiological outcome | Netherlands | Nationwide retrospective cohort study with systematic literature review | 1995-2020 | 21 | 3 | median 7.2; range 1.9-17.9 | NF1 11; sporadic 10; unknown 0 | Isolated optic nerve glioma only; modified Dodge stage 1A in 12 ONGs and 1A plus 1C cisternal segment in 9 ONGs; one patient had bilateral ONG | First-line treatment for progressive isolated ONG: SAT in 14, complete resection surgery in 4, and radiotherapy in 3 | Radiotherapy; technique not specified | total 52.2; range 52.2-54.0; median 52.2 | median mo 128.4; range mo 27.6-220.8 | Visual; Radiological; Endocrine; Progression |
| Ramírez-Melo | 2024 | Optic pathway gliomas: visual outcomes and prognostic factors | Mexico | Retrospective visual outcome cohort | 2005-2022 | 38 | 12 | median 5.5; range 0.5-17 | NF1 9; sporadic 29; unknown 0 | OPG cohort involving optic nerve, chiasm, hypothalamus and optic tract; detailed anatomical categories reported in study | Treatment included observation, chemotherapy, surgery and radiotherapy according to progression and visual status | Radiotherapy; technique not specified | NR | median mo 60; range mo 12-180 | Visual; Radiological; Progression |
| Goodden | 2014 | The role of surgery in optic pathway/hypothalamic gliomas in children | United Kingdom | Retrospective/prospective surgical cohort | 1998-2011 | 42 | 5 | median 5.58; mean 7.25; range 1.08-16.67 | NF1 19; sporadic 23; unknown 0 | OPHG cohort excluding pure optic nerve tumors; MRI location pie chart: optic nerves/chiasm/hypothalamus 45%, optic nerves/chiasm 33%, hypothalamus only 12%, chiasm only 7%, chiasm/hypothalamus 3%; non-NF1 tumors usually hypothalamic | Individualized observation, chemotherapy, surgery and radiotherapy; surgery used for diagnosis, tumor control or relief of mass effect | Radiotherapy; technique not specified | NR | median mo 77; range mo 21.8-142.3 | Visual; Radiological; Endocrine; Progression; Survival; Adverse events |
| Collett-Solberg | 1997 | Endocrine outcome in long-term survivors of low-grade hypothalamic/chiasmatic glioma | USA | Retrospective endocrine outcome cohort | 1973-1994 | 68 | 38 | mean 5; range 0.2-20 | NF1 22; sporadic 46; unknown 0 | All analyzed patients had hypothalamic/chiasmatic glioma located in the hypothalamic-chiasmatic region on MRI or CT | Management included radiotherapy, surgery, chemotherapy, combinations of therapies or conservative observation, with endocrine outcomes assessed retrospectively | Cranial field irradiation | range 45-60; median 55.8; mean 53.8 | median mo 43.2 | Endocrine; Adverse events |
| Gan | 2015 | Neuroendocrine Morbidity After Pediatric Optic Gliomas: A Longitudinal Analysis of 166 Children Over 30 Years | United Kingdom | Retrospective longitudinal OP/HSG morbidity cohort | 1980-2010 | 166 | 69 | median 4.9; range 0.2-15.4 | NF1 68; sporadic 98; unknown 0 | OP/HSG cohort including modified Dodge stage 1 in 29, stage 2 in 76, stage 3/4 in 34 and other midline tumors in 27; hypothalamic involvement 67, leptomeningeal metastases 6 and hydrocephalus 61 | Initial management included observation, surgery or decompression, radiotherapy, chemotherapy and combinations; final treatments included observation only 38, surgery only 21, RT only 15, chemotherapy only 20, multimodality groups and cumulative RT/chemotherapy exposure | Focal radiotherapy | total 48-55; fractions 25-30; range 48-55 | median mo 99.6; range mo 0.48-321.6 | Visual; Endocrine; Progression; Survival; Adverse events |
| Massimino | 2002 | High Response Rate to Cisplatin/Etoposide Regimen in Childhood Low-Grade Glioma | Italy | Prospective chemotherapy cohort / phase II-style treatment series | 1991-2000 | 29 visual pathway glioma | 1 | median 3.75; range 0.33-16.5 | NF1 8; sporadic 26; unknown 0 | Childhood LGG cohort predominantly visual pathway; tumor location chiasmatic/hypothalamic 29, frontal 2, temporal 2, spinal 1; metastatic disease 4 including subarachnoid 3 and spinal 1 | Cisplatin and etoposide chemotherapy for all patients to avoid radiotherapy; surgery or other treatment used only after progression or selected cases | Radiotherapy; technique not specified | NR | median mo 44; range mo 10-120 | Visual; Radiological; Endocrine; Progression; Survival; Adverse events |
| Laithier | 2003 | Progression-Free Survival in Children With Optic Pathway Tumors: Dependence on Age and the Quality of the Response to Chemotherapy—Results of the First French Prospective Study for the French Society of Pediatric Oncology | France and Belgium | Prospective chemotherapy-first trial | 1990-1998 | 85 | 28 | median 2.75 at chemotherapy start; 1.42 at diagnosis; range 0.33-13.67 at chemotherapy start; 0-10.25 at diagnosis | NF1 23; sporadic 62; unknown 0 | Progressive optic pathway tumors; Dodge A 2, Dodge A/B combined 27, Dodge C 58 with extension beyond chiasm; metastases or multicentric disease in 9 | Chemotherapy-first strategy with BBSFOP multiagent chemotherapy for 16 months; radiotherapy deferred unless progression despite chemotherapy or physician decision | Radiotherapy; technique not specified | total 50-55; range 50-55 | median mo 78; range mo 21.6-138 | Visual; Radiological; Progression; Survival; Adverse events |
| Sharif | 2006 | Second Primary Tumors in Neurofibromatosis 1 Patients Treated for Optic Glioma: Substantial Risks After Radiotherapy | United Kingdom | Retrospective long-term NF1 OPG safety cohort | NR | 58 | 18 | range 0.1-41 | NF1 58; sporadic 0; unknown 0 | NA | OPG diagnosis based on clinical assessment, neuroimaging features, pathologic examination of biopsy material or combination; visual testing methods not reported | NR | per fraction OPG radiation field; some second MPNSTs arose within radiation field; mean 25.5-50 | median mo 190.8; mean mo 223.2; range mo 0.36-700.8 | NR |
| Merchant | 2009 | Phase II Trial of Conformal Radiation Therapy for Pediatric Low-Grade Glioma | USA | Prospective phase II conformal radiotherapy trial | 1997-2006 | 58 diencephalic tumors | 78 | median 8.9; range 2.2-19.8 | NF1 13; sporadic 65; unknown 0 | Pediatric LGG cohort treated with CRT: diencephalon 58, cerebral hemisphere 3, cerebellum 17; 13 unbiopsied optic pathway gliomas included | Conformal radiotherapy for pediatric LGG indicated for symptoms, imaging progression or residual tumor progression risk at critical sites | Conformal radiation therapy with MRI registration; 75 CRT and 3 IMRT | total 54; per fraction 1.8; fractions 30; range 50.4-54; median 54 | median mo 89 | Visual; Endocrine; Progression; Survival; Adverse events |
| Petronio | 1991 | Management of chiasmal and hypothalamic gliomas of infancy and childhood with chemotherapy | USA | Retrospective chemotherapy cohort / nonrandomized clinical trial experience | 1983-1989 | 19 | 4 | median 3.2; range 0.29-15.6 | NF1 4; sporadic 14 non-NF1 including 1 tuberous sclerosis; unknown 1 lost to follow-up status NR | Chiasmal and hypothalamic gliomas involving optic chiasm, hypothalamus and retrochiasmal optic pathways; exact distribution not tabulated | Systemic chemotherapy for progressive symptomatic or radiographically enlarging chiasmal-hypothalamic glioma, with radiotherapy reserved for progression | Conventionally fractionated external-beam radiotherapy for salvage | total 50-60; range 50-60 | median mo 18.2; range mo 1.5-74.3 | Visual; Radiological; Endocrine; Progression; Adverse events |
| Kestle | 1993 | Moyamoya phenomenon after radiation for optic glioma | Canada | Retrospective cohort / radiation-associated vasculopathy case series | 1971-1990 | 47 | 28 | NR | NF1 10; sporadic 37; unknown 0 | Optic pathway astrocytoma cohort: 26 involved optic chiasm, 12 centered primarily in hypothalamus, 8 involved a single optic nerve, and 1 involved both optic nerves | Operative management with or without radiotherapy; study focused on later moyamoya phenomenon after RT | Radiotherapy; technique not specified | range 25-55; mean 50.16 | NR | Adverse events |
| Vaidya | 2024 | Magnetic Resonance Imaging Features of Sporadic Optic Chiasmatic-Hypothalamic Gliomas and Correlation with Histopathology and BRAF Gene Alterations | India | Retrospective imaging-histopathology-radiogenomic cohort | 2006-2019 | 26 | 15 | median 9.5; range 1-34 | NF1 0; sporadic 26; unknown 0 | Sporadic optic chiasmatic-hypothalamic gliomas; 24 epicentered in hypothalamic-chiasmatic region and 2 in optic tract; thalamic/gangliocapsular extension 10, temporal lobe extension 5, frontal lobe extension 2, leptomeningeal dissemination 1 | Standard treatment based on age, clinical presentation and disease extent; either surgical debulking followed by external-beam radiotherapy or primary chemoradiotherapy | External-beam radiotherapy; technique not specified | NR | median mo 30; range mo 11-120 | Radiological; Progression |
| Dalla Via | 2007 | Visual outcome of a cohort of children with neurofibromatosis type 1 and optic pathway glioma followed by a pediatric neuro-oncology program | Italy | Prospective protocol-based cohort / retrospective outcome analysis | 1994-2005 | 20 | 8 | median 2.42; mean 3.33; range 1.33-9.58 | NF1 20; sporadic 0; unknown 0 | NF1-associated OPG; Dodge 1 in 1, Dodge 2 in 7, Dodge 3 in 12; tumors involved optic nerve/chiasm/tract or hypothalamic pathway according to Dodge classification | Observation for stable disease; treatment only for progressive disease or clinical deterioration, with carboplatin/vincristine chemotherapy for younger children and fractionated stereotactic radiotherapy for older children | External conventional radiotherapy via fractionated guided stereotactic technique | total 54; per fraction 1.8; fractions 30; range 54; median 54; mean 54 | median mo 76; mean mo 81; range mo 5-216 | Visual; Radiological; Endocrine; Progression |
| Awdeh | 2012 | Visual Outcomes in Pediatric Optic Pathway Glioma After Conformal Radiation Therapy | USA | Prospective conformal radiation therapy visual-outcome cohort | 1997-2002 | 20 | 20 | median 9.3; range 3.2-14.6 | NF1 0; sporadic 20; unknown 0 | All tumors in hypothalamus, optic chiasm or optic tracts; 3 limited to optic chiasm, 2 involved hypothalamus, chiasm and proximal optic nerves, remainder largely hypothalamic/chiasmatic | Conformal radiation therapy 54 Gy for all evaluable children; some had chemotherapy, biopsy or subtotal resection before CRT | Conformal radiation therapy | total 54; per fraction 1.8; fractions 30; range 54; median 54; mean 54 | median mo 30; range mo 8-62 | Visual |
| Green | 2022 | A nationwide service evaluation of safety, radiologic and visual outcome refining bevacizumab-based treatments in children with progressive low-grade glioma | United Kingdom | Conference abstract / nationwide retrospective service evaluation | 2009-2020 | 88% OPG; exact count NR | 0 | NR | NF1 24% NF1; exact count NR | Progressive pediatric low-grade glioma cohort, 88% optic pathway glioma; detailed tumor-location distribution not reported in abstract | Bevacizumab-based treatment for progressive PLGG after radiological, visual or combined progression, mostly third-line or later | NA | NR | NR | Visual; Radiological; Progression; Survival; Adverse events |
| Morin | 2024 | Very long-term outcomes of pediatric patients treated for optic pathway gliomas: A longitudinal cohort study | France | Longitudinal retrospective cohort of 5-year childhood OPG survivors | 1980-2015 | 182 | 75 | median 3.4; range 0.33-16.37 | NF1 65; sporadic 117; unknown 0 | OPG cohort: optic nerve only 18, chiasmatic involvement 111, retrochiasmatic involvement 53, hypothalamic involvement 56, second location at diagnosis 26, leptomeningeal dissemination 7 | First-line treatment: systemic treatment alone 58, local treatment alone 51, combined systemic and local treatment 45, and monitoring 28; overall treatments included surgery, RT, systemic therapy and targeted therapy | Focal radiotherapy | range 50-60 in cerebrovascular-event subgroup; median 50 | median mo 206.4; range mo 64.8-488.4 | Visual; Endocrine; Progression; Survival; Adverse events |
| Fernandes | 2018 | Long term follow-up of optic pathway gliomas in children with neurofibromatosis type 1: An oncology hospital experience | Portugal | Conference abstract / retrospective cohort | 1991-2017 | 62 | 1 | median 4 | NF1 62; sporadic 0; unknown 0 | NF1-associated optic pathway gliomas in children; detailed optic nerve, chiasm, hypothalamus or tract distribution not reported in abstract | Careful surveillance for most patients; treatment applied for clinical or imaging progression | Radiotherapy; technique not specified | NR | mean mo 94.8; range mo Up to 264 | Visual; Radiological; Progression |
| Varan | 2013 | Optic Glioma in Children: A Retrospective Analysis of 101 Cases | Turkey | Retrospective clinical outcome cohort | 1975-2008 | 101 | 40 | median 6; range 0.08-18 | NF1 53; sporadic 48; unknown 0 | Optic glioma cohort including optic chiasm plus hypothalamus 32, intraorbital optic nerve 16, intraorbital optic nerve plus prechiasm 10, intraorbital optic nerve plus optic chiasm 10, optic chiasm plus hypothalamus plus optic tract 9, diffuse visual pathway glioma 8, intracranial optic nerve plus optic chiasm 6, intracranial optic nerve plus optic chiasm plus hypothalamus 4, prechiasm plus chiasm plus postchiasm 4, bilateral optic nerve 1, chiasm only 1 | Treatment consisted of observation, surgery, radiotherapy and chemotherapy according to symptoms, progression, age and tumor location | Conventional external-beam photon radiotherapy with Co60 or 6-MV photons | total 40.8-60; per fraction 1.7-2; range 40.8-60; mean 54 | median mo 96 | Progression; Survival; Adverse events |
| Nicolin | 2009 | Natural History and Outcome of Optic Pathway Gliomas in Children | Canada | Retrospective natural-history and treatment-outcome cohort | 1990-2004 | 133 | 16 | mean 5.89; range 0.34-16.8 | NF1 78; sporadic 55; unknown 0 | OPG locations: hypothalamic/chiasmatic 50, unilateral optic nerve 29, HC plus bilateral optic nerves 19, HC plus unilateral optic nerve 10, bilateral optic nerves 10, HC plus optic nerves plus optic radiations 9, HC plus thalamus 3, HC plus optic radiations 1, HC plus dissemination 2 | Observation initially in 87 and immediate treatment in 46; final first treatment among treated included chemotherapy, debulking plus chemotherapy, GTR, radiotherapy, debulking plus radiotherapy or debulking only | Radiotherapy; technique not specified | NR | median mo 103.2; mean mo 108; range mo 6.72-216 | Visual; Endocrine; Progression; Survival; Adverse events |
| Campagna | 2010 | Optic Pathway Glioma: Long-Term Visual Outcome in Children Without Neurofibromatosis Type-1 | Italy | Retrospective visual-outcome cohort | 1989-2008 | 32 | 17 | median 4.67; range 0.42-13 | NF1 0; sporadic 32; unknown 0 | Non-NF1 pediatric OPG cohort; Dodge I in 5 and Dodge III in 27; most tumors involved or extended to the hypothalamic-chiasmatic region | Observation, chemotherapy, radiotherapy, surgery when feasible, and subsequent treatment for clinical or radiological progression according to SIOP LGG strategies | External conventional radiotherapy | total 54; per fraction 1.8; fractions 30; range 54; median 54 | median mo 73; range mo 6-168 | Visual; Radiological; Endocrine; Progression |
| Shofty | 2015 | The Effect of Chemotherapy on Optic Pathway Gliomas and Their Sub-Components: A Volumetric MR Analysis Study | Israel | Retrospective chemotherapy-treated volumetric MRI cohort | 1990-2013 | 15 | 5 | mean 6.5; range 1-23 | NF1 10; sporadic 5; unknown 0 | Hypothalamic/chiasmatic OPG cohort; all patients Dodge II or III at diagnosis; tumors epicentered in hypothalamic/chiasmatic area | Chemotherapy-treated progressive hypothalamic/chiasmatic OPG with longitudinal volumetric MRI assessment | Radiotherapy; technique not specified | NR | mean mo 41; range mo 12-96 | Radiological; Progression |
| Bennebroek | 2024 | Treatment of isolated pediatric optic nerve glioma: A nationwide retrospective cohort study and systematic literature review on visual and radiological outcome | Netherlands | Nationwide retrospective cohort study with systematic literature review | 1995-2020 | 21 | 3 | median 7.2; range 1.9-17.9 | NF1 11; sporadic 10; unknown 0 | Isolated optic nerve glioma only; modified Dodge stage 1A in 12 ONGs and 1A plus 1C cisternal segment in 9 ONGs; one patient had bilateral ONG | First-line treatment for progressive isolated ONG: SAT in 14, complete resection surgery in 4, and radiotherapy in 3 | Radiotherapy; technique not specified | total 52.2; range 52.2-54.0; median 52.2 | median mo 128.4; range mo 27.6-220.8 | Visual; Radiological; Endocrine; Progression |
| Regueiro | 1995 | Radiotherapy in the management of optic pathway gliomas | Spain | Conference abstract / retrospective radiotherapy cohort | NR | 35 | 35 | NR | NR | Optic pathway glioma cohort including 7 optic nerve tumors and 28 chiasmal tumors; RT-alone group included 6 optic nerve and 19 chiasmal tumors; subtotal surgery plus RT group included 1 optic nerve and 9 chiasmal tumors | Radiotherapy alone in 25 patients and subtotal surgery followed by postoperative radiotherapy in 10 patients | Radiotherapy; technique not reported | NR | NR | Progression; Survival |
| Combs | 2005 | Fractionated Stereotactic Radiotherapy of Optic Pathway Gliomas: Tolerance and Long-Term Outcome | Germany | Retrospective radiotherapy cohort | 1990-2003 | 15 | 15 | median 6.9; range 0.67-33 | NF1 3; sporadic 12; unknown 0 | Optic pathway glioma cohort treated with FSRT; tumor confined to optic chiasm in 5, optic chiasm plus optic nerves in 3, optic nerve only in 3, and suprasellar/pituitary region in 4 | FSRT for all patients, mostly after prior neurosurgical intervention, without concomitant chemotherapy | Fractionated stereotactic radiotherapy | total 52.2; per fraction 1.8; range 45.2-57.6; median 52.2 | median mo 97; range mo 8-151 | Visual; Endocrine; Progression; Survival; Adverse events |
| Rakotonjanahary | 2015 | Mortality in Children with Optic Pathway Glioma Treated with Up-Front BB-SFOP Chemotherapy | France | Historical cohort study | June 1990-December 2004 | 180 | 55 subsequent RT after first-line chemotherapy | median 2.4 | NF1 60; sporadic 120; unknown 0 | Optic pathway glioma cohort; exact anatomic subgroup distribution not reported in extractable main text; included clinically/radiologically diagnosed OPG, including symptomatic/progressive tumors treated with chemotherapy | NA | NA | NR | median mo 163.2; range mo 73.2-283.2 | Survival; Adverse events |
| Liu | 2022 | The role of imaging features and resection status in the survival outcome of sporadic optic pathway glioma children receiving different adjuvant treatments | China | Retrospective surgical cohort | 2010-2019 | 165 | 92 | NR | NF1 0; sporadic 165; unknown 0 | Sporadic pediatric OPG cohort after primary intratumor debulking; optic chiasm involved 162/165, hypothalamus 131/165, optic nerve only 3/165, optic tract 24/165, leptomeningeal metastases 17/165 | Primary intratumor debulking surgery for all patients followed by first adjuvant treatment: radiotherapy, chemotherapy, or observation | Radiotherapy after surgery; technique not specified | NR | median mo 39; range mo 23-68 | Endocrine; Progression; Survival; Adverse events |
| Acharya | 2019 | Long-term visual acuity outcomes after radiation therapy for sporadic optic pathway glioma | USA | Retrospective RT cohort with prospective protocol subset | 1997-2017 | 41 | 41 | median 8; range 4.1-19.8 | NF1 0; sporadic 41; unknown 0 | Sporadic OPG treated with RT; posterior extent pre-chiasm 2, chiasm 1, post-chiasm/hypothalamus 38 | Definitive RT for sporadic OPG after radiographic progression or visual deterioration, with or without prior surgery or chemotherapy | 3D conformal photon therapy, intensity-modulated photon therapy, or intensity-modulated proton therapy | total 54 photon; 52.2 GyRBE proton; per fraction 1.8; fractions 30 photon; 29 proton; range 52.2-54 | mean mo 60; range mo 2.9-147.6 | Visual; Progression |
| Zhou | 2024 | Visual deterioration outcomes following optic pathway glioma treatment: a 12-year single institution retrospective study | China | Retrospective cohort | January 2011-December 2022 | 140 | 82 | mean 6.9; range 1-26 | unknown 140 | Pediatric OPG surgical cohort; Dodge I 2, Dodge II 29, Dodge III 109; anatomical type A 11, type M 31, type P 98; lateral extension 42; leptomeningeal dissemination 6 | Initial partial tumor resection for all patients, followed by adjuvant RT, chemotherapy, RT plus chemotherapy, or observation | Radiotherapy; modality not specified | NR | mean mo 71; range mo 12-137 | Visual; Progression |
| Quesada | 2019 | Visual Outcomes After Radiation Therapy for Optic Pathway Glioma | USA | Conference abstract / retrospective RT cohort | 1997-2017 | 40 | 40 | median 8.2; range 1-19 | NF1 3; sporadic 37; unknown 0 | OPG treated with RT; majority involved postchiasmatic optic tracts (90%); prechiasmatic location analyzed as risk factor for VA decline | Radiotherapy for all patients with serial ophthalmologic follow-up | Radiation therapy; modality not specified | NR | median mo 37.2 | Visual |

Characteristics of the 49 studies included in the systematic review. The table summarizes first author, publication year, study title, country, study design, study period, eligible OPHG population, radiotherapy/radiosurgery subgroup size, age, NF1 status, tumor location, prior treatment or management, radiotherapy/radiosurgery modality, dose and fractionation, follow-up duration, and reported outcomes.

**Table 2. Patient, tumor, and treatment characteristics.**

| Study_ID | Total_N | Eligible_OPHG_N | RT_SRS_N | Age_Years | Sex | Pediatric_Adult | NF1_Status | Tumor_Location | Baseline_Visual_Impairment | Prior_Surgery | Prior_Chemotherapy | RT_SRS_Modality | Dose_Fractionation | RT_Timing_Indication | Follow_Up |
| --- | --- | --- | --- | --- | --- | --- | --- | --- | --- | --- | --- | --- | --- | --- | --- |
| Kovalic_1990 | 35 treated; 33 analyzed | 33 | 33 | median 4; range 1-34 | M 12; F 21 | Pediatric 25 age <=15 years; text also states most were under 16; Adult 8 age >15 years | NF1 11; Sporadic 22; Unknown 0 | Optic glioma cohort: 5 optic nerve only, 8 optic nerve plus chiasm, and 20 optic nerve plus chiasm plus contiguous structures; most had chiasmal or extrachiasmal extension | visual symptoms 20 optic atrophy and 20 visual field defect; visual acuity recorded in 19; VA impairment 10 of 19 had visual acuity worse than 20/40 in affected eye; VF defect 20; blindness 8 total blindness; 6 light perception only; optic atrophy 20 | any 22; biopsy 14; debulking/partial 8; STR 8 partial excision | NR | Local-field radiation therapy using orthovoltage, cobalt teletherapy or megavoltage photon therapy | total 50.4 median Gy; 0.76-2.0; median 1.6 Gy/fraction; range 15.22-61.0 including one incomplete course; otherwise 37.86-61.0 Gy; median 50.4 Gy | first-line 33; adjuvant 22 postoperative after biopsy or partial excision; after surgery 22; indication Optic nerve or chiasmal glioma treated with local irradiation; authors concluded RT was beneficial for chiasmal involvement and incomplete resections | median 147.6 mo; range 24-372 among survivors mo |
| Kelly_2012 | 69 OPG cohort; 21 analyzed | 21 | 11 | range 0.7-9 at treatment | M 12; F 9 | Pediatric 21; Adult 0 | NF1 6; Sporadic 15; Unknown 0 | Bilateral OPG cohort involving chiasm alone or with anterior/posterior extension; Table 1 locations included chiasm in most patients, optic nerve involvement in selected patients, hypothalamus in 6, optic tracts in 6 and visual radiations in 2 | visual symptoms 17 reduced visual acuity; all had reduced VEP in one or both eyes; VA impairment 17; VF defect 11 of 11 tested; optic atrophy 17 optic nerve pallor | any 2 biopsy-confirmed diagnoses; exact surgery count NR; biopsy 2 biopsy-confirmed diagnoses | any 18; subgroup 18; regimen Vincristine/carboplatin most common; other regimens included TPCV, vincristine/actinomycin, temozolomide, vinblastine/carboplatin, vinblastine and lenalidomide | External-beam radiation therapy | NR | first-line 3; salvage 8; after chemotherapy 8; indication Primary treatment in older children without NF1 or after failed chemotherapy because of progressive tumor growth or vision loss | mean 108 mo; range 48-192 mo |
| Erkal_1997 | 33 | 33 | 33 | median 7; range 2-18 | M 11; F 22 | Pediatric 33; Adult 0 | NF1 6; Sporadic 27; Unknown 0 | Twenty-four optic pathway gliomas including 4 single optic nerve and 20 optic nerve plus chiasm; nine chiasmatic-hypothalamic gliomas involving chiasm, third ventricle and/or hypothalamus | visual symptoms 31; VA impairment 31; optic atrophy 10 | any 29; biopsy 7; debulking/partial 22 subtotal tumor resection; STR 22 | any 1; subgroup 1; regimen Combination chemotherapy regimen after progression following RT in one chiasmatic-hypothalamic glioma; regimen not specified | External-beam radiotherapy using cobalt-60 unit | total 40-60 Gy; 1.8-2 Gy/fraction; 20-30 fractions; range 40-60 Gy; median 50 Gy | first-line 31; adjuvant 29 after subtotal resection or biopsy before RT; salvage 2; after surgery 29; indication Radiotherapy at diagnosis for most patients; for NF1 or single optic nerve tumors RT started with progressive visual deterioration; two received RT for progression after subtotal resection | mean 163.2 mo; range 6-193.2 mo |
| Khafaga_2003 | 50 | 50 | 28 initial RT; 35 any RT over course | median 4; range 2-36 | M 22; F 28 | Pediatric 41 younger than 10 years; exact under-18 count NR; Adult 9 age >=10 includes some adults up to 36 | NF1 5; Sporadic 45; Unknown 0 | Optic pathway low-grade glioma cohort; optic nerve only 6, chiasm 3, optic nerve plus chiasm 15, chiasm plus hypothalamus 23, chiasm plus optic tract 3 | visual symptoms 39 decreased vision at presentation; blindness at presentation in 13; nystagmus in 7; VA impairment 39 decreased vision; blindness 13 | any 34; biopsy 22; debulking/partial 6; STR 3; GTR 3 | any 7; subgroup 7; regimen Chemotherapy regimens not specified; 7 patients received chemotherapy during disease course | Radiotherapy; technique not reported | total 45-54 Gy; 1.6-1.8 Gy/fraction; 25-30 fractions; range 45-54 Gy | first-line 28; salvage 7; after surgery 34; indication Initial or salvage treatment for optic pathway glioma; RT used in 28 initially and later in 7 additional patients | median 84 mo; range 28.8-198 mo |
| Tsang_2017 | 89 | 89 | 89 | median 7.9 at RT start; median 4.8 at diagnosis; range 1.1-19.8 at RT start | M 41; F 48 | Pediatric 89; Adult 0 | NF1 19; Sporadic 70; Unknown 0 | Pediatric OPG treated with RT; optic chiasm involved in 71, hypothalamus in 64, optic nerve in 41, optic tract in 30, optic radiation in 11, and other intracranial extension in selected patients | visual symptoms 75 visual acuity loss; 37 visual field deficit; VA impairment 75; VF defect 37; blindness 11 | any 39; biopsy 29 biopsy only; debulking/partial 10 | any 47; subgroup 47; regimen Chemotherapy regimens varied across era; not specified in extract | Radiotherapy; external-beam photon or proton, including conformal techniques in later era | total 50.4-54 Gy; 1.8 Gy/fraction; range 45-59.4 Gy; median 54 Gy | first-line 25; salvage 64; after surgery 39; after chemotherapy 47; indication Progressive or symptomatic OPG treated with RT; included upfront and salvage RT after chemotherapy, surgery or observation | median 105.6 mo; range 5.4-351.6 mo |
| Segal_2010 | 57 NF1-OPG cohort; 42 with follow-up data | 42 | 3 | median 6 at OPG diagnosis | M NR; F NR | Pediatric 42; Adult 0 | NF1 42; Sporadic 0; Unknown 0 | NF1-associated OPG; tumor location included optic nerve, chiasm, optic tract and hypothalamic involvement according to clinical and imaging records | visual symptoms 16 symptomatic at diagnosis; VA impairment 8 decreased acuity; VF defect 5 field defects | any NR | any 12; subgroup 12; regimen Carboplatin/vincristine and other chemotherapy regimens not detailed in extract | Radiotherapy; technique not specified | NR | salvage 3; indication Used rarely for progressive NF1-associated OPG | mean 108 mo |
| Liu_2018 | 37 | 37 | 4 | mean 6.7; range 0.75-17 | M 19; F 18 | Pediatric 37; Adult 0 | NF1 0; Sporadic 37; Unknown 0 | Pediatric OPG cohort; tumors involved optic nerve, chiasm, hypothalamus and optic tract in variable combinations; Dodge classification reported in study | visual symptoms 34 decreased vision; VA impairment 34; VF defect NR | any 29; biopsy 6; debulking/partial 23 | any 12; subgroup 12; regimen Chemotherapy regimens not specified | Radiotherapy; technique not specified | NR | adjuvant 4; after surgery 4; indication Adjuvant treatment in selected children after surgery or progression | median 37 mo; range 6-107 mo |
| Han_2007 | 14 | 14 | 14 | mean 12 at Novalis RS; 10.2 at initial diagnosis; range 5-16 at Novalis RS; 4-13 at initial diagnosis | M 7; F 7 | Pediatric 14; Adult 0 | NF1 NR; Sporadic NR; Unknown 14 | Optic pathway gliomas involving optic nerve, chiasm, optic tract or hypothalamus; detailed location distribution not fully extractable | VA impairment 14; VF defect NR | any 12; debulking/partial 12 | any 7; subgroup 7; regimen Chemotherapy regimens before FSRT not specified | Fractionated stereotactic radiotherapy with Novalis system | total 45-50 Gy; 1.8-2 Gy/fraction; 25 fractions; range 45-50 Gy; median 50 Gy | salvage 14; after surgery 12; after chemotherapy 7; indication FSRT for residual, recurrent or progressive OPG after prior surgery and/or chemotherapy | median 34 mo; range 16-72 mo |
| Mahoney_2000 | 42 | 42 | 10 | median 1.96; range 0.33-5.5 | M 19; F 23 | Pediatric 42; Adult 0 | NF1 26; Sporadic 16; Unknown 0 | Pediatric OPG cohort involving optic nerve, chiasm, tract, hypothalamus or radiations; anatomic subgroups reported by Dodge classification | VA impairment 42 ophthalmologic follow-up cohort; VF defect NR | any 10; biopsy NR; debulking/partial NR | any 15; subgroup 15; regimen Chemotherapy regimens not specified | Radiotherapy; technique not specified | NR | salvage 10; indication Used in selected patients with progressive visual or tumor findings after observation or chemotherapy | median 59 mo; range 6-193 mo |
| Packer_1988 | 24 | 24 | 4 | median 2.4; range 0.2-5.5 | M 11; F 13 | Pediatric 24; Adult 0 | NF1 5; Sporadic 19; Unknown 0 | Chiasmatic/hypothalamic glioma cohort; tumors involved optic chiasm, hypothalamus and adjacent optic pathway structures | visual symptoms 24 visual symptoms or signs part of eligibility; VA impairment NR; VF defect NR | any 19; biopsy 5; debulking/partial 14 | any 24; subgroup 24; regimen Actinomycin D, vincristine, procarbazine and other agents in protocol-era chemotherapy | Conventional external-beam radiotherapy | NR | salvage 4; after chemotherapy 4; indication Used for tumor progression after chemotherapy | median 51 mo; range 3-120 mo |
| Ge_2022 | 20 | 20 | 20 | median 11; range 2-53 | M 7; F 13 | Pediatric 14 under 18; Adult 6 | NF1 NR; Sporadic NR; Unknown 20 | Optic pathway gliomas treated with Gamma Knife; lesions involved optic nerve, optic chiasm, optic tract or hypothalamic region | visual symptoms 20; VA impairment 20; VF defect NR | GTR 2 prior total resection; other surgery details NR | NR | Gamma Knife stereotactic radiosurgery | total 8-14 Gy; 8-14 Gy/fraction; 1 fractions; range 8-14 Gy; median 11.5 Gy | salvage 20; indication SRS for optic pathway glioma; generally selected lesions suitable for Gamma Knife treatment | median 43 mo; range 12-104 mo |
| Janss_1995 | 44 | 44 | 12 | median 1.8; range 0.08-5.0 | M 22; F 22 | Pediatric 44; Adult 0 | NF1 12; Sporadic 32; Unknown 0 | Optic pathway and hypothalamic/chiasmatic gliomas in children younger than 5 years; detailed location included optic nerve, chiasm, hypothalamus and posterior pathway involvement | visual symptoms 36; VA impairment NR; VF defect NR | any 27; biopsy 14; debulking/partial 13 | any 34; subgroup 34; regimen Chemotherapy regimens not specified | Radiotherapy; technique not specified | NR | salvage 12; after chemotherapy 12; indication RT generally delayed or used for progressive disease in young children | median 72 mo; range 6-168 mo |
| Fouladi_2003 | 91 | 91 | NR | median 3.2; range 0.25-15.0 | M 47; F 44 | Pediatric 91; Adult 0 | NF1 NR; Sporadic NR; Unknown 91 | Hypothalamic/chiasmatic tumors including optic pathway gliomas; tumors involved optic chiasm, hypothalamus and adjacent structures | visual symptoms NR; VA impairment NR; VF defect NR | any NR | any NR; regimen Protocol-based chemotherapy according to Pediatric Oncology Group treatment assignment | Radiotherapy; technique not specified | NR | NR | median 60 mo |
| Hanania_2021 | 83 | 83 | 18 | median 4.1 | M 39; F 44 | Pediatric 83; Adult 0 | NF1 37; Sporadic 46; Unknown 0 | OPG cohort involving optic nerve, chiasm, tract, hypothalamus and posterior visual pathway in variable combinations | visual symptoms NR; VA impairment NR; VF defect NR | any NR | any NR; regimen Chemotherapy regimens varied by era; carboplatin/vincristine and other regimens reported in institution | Radiotherapy; technique not specified | NR | salvage 18; indication RT used in selected patients with progressive or refractory disease | median 96 mo; range 6-300 mo |
| Picariello_2022 | 102 | 102 | 25 | median 4.3; range 0.06-3.00 | M 50; F 52 | Pediatric 102; Adult 0 | NF1 47; Sporadic 55; Unknown 0 | Pediatric OPG cohort with optic nerve, chiasmatic, hypothalamic and posterior pathway involvement | visual symptoms NR; VA impairment NR; VF defect NR | any NR | any NR; regimen Chemotherapy regimens varied; not fully extractable | Radiotherapy; technique not specified | NR | salvage 25; indication RT used in selected progressive or refractory cases | median 84 mo; range 12-240 mo |
| Bennebroek_2025 | 198 | 198 | 4 | median 4.4; range 0.3-16.5 | M 91; F 107 | Pediatric 198; Adult 0 | NF1 113; Sporadic 85; Unknown 0 | OPG cohort excluding isolated optic nerve glioma; tumors included optic chiasm, hypothalamus, optic tract and posterior pathway involvement | visual symptoms NR; VA impairment NR; VF defect NR | any NR | any NR; regimen Chemotherapy regimens varied, including vincristine/carboplatin and other systemic therapies | Radiotherapy; technique not specified | NR | salvage 4; indication RT rarely used in modern pediatric OPG cohort | median 90 mo; range 12-300 mo |
| Liao_2020 | 45 | 45 | 15 | median 7.7 | M 24; F 21 | Pediatric 45; Adult 0 | NF1 0; Sporadic 45; Unknown 0 | Sporadic pediatric chiasmatic-hypothalamic glioma; tumors involved chiasm and hypothalamic region; included prechiasmatic, chiasmatic and postchiasmatic extension | visual symptoms 45; VA impairment 45; VF defect NR; blindness NR | any 45; debulking/partial 45 | any 19; subgroup 19; regimen Chemotherapy regimens not specified | Radiotherapy after surgery; technique not specified | NR | adjuvant 15; after surgery 15; indication Postoperative radiotherapy in selected surgically treated sporadic chiasmatic-hypothalamic glioma patients | median 35 mo; range 3-120 mo |
| Rajagopal_2023 | 59 | 59 | 13 | median 5.2; range 0.5-14 | M 28; F 31 | Pediatric 59; Adult 0 | NF1 29; Sporadic 30; Unknown 0 | Pediatric OPG cohort with optic nerve, chiasm, hypothalamus and posterior visual pathway involvement | visual symptoms NR; VA impairment NR; VF defect NR | any NR | any NR; regimen Chemotherapy regimens varied; not fully extractable | Radiotherapy; technique not specified | NR | salvage 13; indication RT used for progressive or refractory disease | median 72 mo; range 6-216 mo |
| RamirezMelo_2024 | 38 | 38 | 12 | median 5.5; range 0.25-17 approx | M 18; F 20 | Pediatric 38; Adult 0 | NF1 9; Sporadic 29; Unknown 0 | OPG cohort involving optic nerve, chiasm, hypothalamus and optic tract; detailed anatomical categories reported in study | visual symptoms NR; VA impairment NR; VF defect NR | any NR | any NR; regimen Chemotherapy regimens not specified | Radiotherapy; technique not specified | NR | salvage 12; indication RT used in selected patients with progression or visual decline | median 60 mo; range 12-180 mo |
| Lim_2022 | 57 | 57 | 9 | median 4.8 | M 30; F 27 | Pediatric 57; Adult 0 | NF1 32; Sporadic 25; Unknown 0 | Pediatric OPG involving optic nerve, optic chiasm, optic tract, hypothalamus and posterior visual pathway according to imaging | visual symptoms NR; VA impairment NR; VF defect NR | any NR | any NR; regimen Chemotherapy regimens varied; not fully extractable | Radiotherapy; technique not specified | NR | salvage 9; indication RT used for selected progressive or refractory OPG | median 78 mo; range 6-221 mo |
| Lohkamp_2022 | 60 | 60 | 14 | median 4.6; range 0.8-17.04 | M 29; F 31 | Pediatric 60; Adult 0 | NF1 28; Sporadic 32; Unknown 0 | OPG cohort involving optic nerve, chiasm, hypothalamus and optic tract/posterior pathway structures | visual symptoms NR; VA impairment NR; VF defect NR | any NR | any NR; regimen Chemotherapy regimens varied; not fully extractable | Radiotherapy; technique not specified | NR | salvage 14; indication RT used in selected patients during long-term management | median 108 mo; range 12-312 mo |
| ElShehaby_2016 | 35 | 35 | 35 | median 16; range 5-43 | M 18; F 17 | Pediatric 23 under 18; Adult 12 | NF1 NR; Sporadic NR; Unknown 35 | OPG treated with Gamma Knife; locations included optic nerve, optic chiasm, optic tract and hypothalamic region in selected patients | visual symptoms 35; VA impairment 35; VF defect NR | any NR | NR | Gamma Knife radiosurgery | total 8-14 Gy; 8-14 Gy/fraction; 1 fractions; range 8-14 Gy; median 10 Gy | salvage 35; indication Gamma Knife radiosurgery for optic pathway glioma selected for radiosurgical management | median 60 mo; range 12-180 mo |
| Goodden_2014 | 42 | 42 | 5 | median 5.58; mean 7.25; range 1.08-16.67 | M 24; F 18 | Pediatric 42; Adult 0 | NF1 19; Sporadic 23; Unknown 0 | OPHG cohort excluding pure optic nerve tumors; MRI location pie chart: optic nerves/chiasm/hypothalamus 45%, optic nerves/chiasm 33%, hypothalamus only 12%, chiasm only 7%, chiasm/hypothalamus 3%; non-NF1 tumors usually hypothalamic | VA impairment NR; VF defect NR | any 28; biopsy 12; debulking/partial 16 | any 20; subgroup 20; regimen Chemotherapy regimens based on institutional pediatric LGG protocols; not fully specified | Radiotherapy; technique not specified | NR | salvage 5; after surgery 5; indication RT used in selected patients after surgery or progression | median 77 mo; range 21.8-142.3 mo |
| CollettSolberg_1997 | 68 | 68 | 38 | mean 5; range 0.2-20 | M 36; F 32 | Pediatric NR; Adult NR | NF1 22; Sporadic 46; Unknown 0 | All analyzed patients had hypothalamic/chiasmatic glioma located in the hypothalamic-chiasmatic region on MRI or CT | visual symptoms NR; VA impairment NR; VF defect NR | any NR | any NR; regimen Chemotherapy in selected patients; details not central to endocrine cohort | Cranial field irradiation | range 45-60 Gy; median 55.8 Gy; mean 53.8 Gy | first-line NR; salvage NR; indication Cranial irradiation as part of management of hypothalamic/chiasmatic glioma | median 43.2 mo |
| Gan_2015 | 166 | 166 | 69 | median 4.9; range 0.2-15.4 | M 85; F 81 | Pediatric 166; Adult 0 | NF1 68; Sporadic 98; Unknown 0 | OP/HSG cohort including modified Dodge stage 1 in 29, stage 2 in 76, stage 3/4 in 34 and other midline tumors in 27; hypothalamic involvement 67, leptomeningeal metastases 6 and hydrocephalus 61 | visual symptoms NR; VA impairment NR; VF defect NR | any 75; biopsy 23; debulking/partial NR | any 85; subgroup 85; regimen Chemotherapy regimens varied over 30 years; not fully specified | Focal radiotherapy | total 48-55 Gy; 25-30 fractions; range 48-55 Gy | first-line 15 RT only and additional multimodality first-line/overall exposure; after surgery NR; after chemotherapy NR; indication RT used as treatment modality in selected children across 30-year cohort | median 99.6 mo; range 0.48-321.6 mo |
| Massimino_2002 | 34 LGG; 29 visual pathway glioma | 29 visual pathway glioma | 1 | median 3.75; range 0.33-16.5 | M 17; F 17 | Pediatric 34; Adult 0 | NF1 8; Sporadic 26; Unknown 0 | Childhood LGG cohort predominantly visual pathway; tumor location chiasmatic/hypothalamic 29, frontal 2, temporal 2, spinal 1; metastatic disease 4 including subarachnoid 3 and spinal 1 | visual symptoms NR; VA impairment NR; VF defect NR | any 10; biopsy 7; debulking/partial 3 | any 34; subgroup 34; regimen Cisplatin plus etoposide | Radiotherapy; technique not specified | NR | salvage 1; after chemotherapy 1; indication RT reserved for selected progression after chemotherapy | median 44 mo; range 10-120 mo |
| Laithier_2003 | 85 | 85 | 28 | median 2.75 at chemotherapy start; 1.42 at diagnosis; range 0.33-13.67 at chemotherapy start; 0-10.25 at diagnosis | M 39; F 46 | Pediatric 85; Adult 0 | NF1 23; Sporadic 62; Unknown 0 | Progressive optic pathway tumors; Dodge A 2, Dodge A/B combined 27, Dodge C 58 with extension beyond chiasm; metastases or multicentric disease in 9 | visual symptoms 85 progressive or symptomatic by trial inclusion; VA impairment NR; VF defect NR | any 43; biopsy 35; debulking/partial 8 | any 85; subgroup 85; regimen BB-SFOP multiagent chemotherapy | Radiotherapy; technique not specified | total 50-55 Gy; range 50-55 Gy | salvage 28; after chemotherapy 28; indication RT delivered after progression or physician decision after chemotherapy-first strategy | median 78 mo; range 21.6-138 mo |
| Sharif_2006 | 58 | 58 | 18 | range 0.1-41 | M NR; F NR | Pediatric NR; Adult NR | NF1 58; Sporadic 0; Unknown 0 | NA | NR | NR | NR | NR | 25.5-50 Gy/fraction | NR | median 190.8 mo; mean 223.2 mo; range 0.36-700.8 mo |
| Merchant_2009 | 78 | 58 diencephalic tumors | 78 | median 8.9; range 2.2-19.8 | M 37; F 41 | Pediatric 78; Adult 0 | NF1 13; Sporadic 65; Unknown 0 | Pediatric LGG cohort treated with CRT: diencephalon 58, cerebral hemisphere 3, cerebellum 17; 13 unbiopsied optic pathway gliomas included | visual symptoms NR; VA impairment NR; VF defect NR | any 46; biopsy 18; debulking/partial 28 | any 20; subgroup 20; regimen Chemotherapy regimens before CRT included carboplatin/vincristine and others; not fully specified | Conformal radiation therapy with MRI registration; 75 CRT and 3 IMRT | total 54 Gy; 1.8 Gy/fraction; 30 fractions; range 50.4-54 Gy; median 54 Gy | first-line NR; salvage NR; indication CRT for symptomatic, progressive, residual or unresectable pediatric low-grade glioma at critical sites | median 89 mo |
| Petronio_1991 | 19 | 19 | 4 | median 3.2; range 0.29-15.6 | M 11; F 8 | Pediatric 19; Adult 0 | NF1 4; Sporadic 14 non-NF1 including 1 tuberous sclerosis; Unknown 1 lost to follow-up status NR | Chiasmal and hypothalamic gliomas involving optic chiasm, hypothalamus and retrochiasmal optic pathways; exact distribution not tabulated | visual symptoms 19; VA impairment NR; VF defect NR | any 12; biopsy 6; debulking/partial 6 | any 19; subgroup 19; regimen Vincristine and actinomycin D with or without other agents; exact regimens varied | Conventionally fractionated external-beam radiotherapy for salvage | total 50-60 Gy; range 50-60 Gy | salvage 4; after chemotherapy 4; indication RT used after progressive disease despite chemotherapy | median 18.2 mo; range 1.5-74.3 mo |
| Kestle_1993 | 47 | 47 | 28 | NR | M NR; F NR | Pediatric NR; Adult NR | NF1 10; Sporadic 37; Unknown 0 | Optic pathway astrocytoma cohort: 26 involved optic chiasm, 12 centered primarily in hypothalamus, 8 involved a single optic nerve, and 1 involved both optic nerves | NR | any 39; biopsy/resection details not fully extractable | NR | Radiotherapy; technique not specified | range 25-55 Gy; mean 50.16 Gy | after surgery NR; indication RT for optic pathway astrocytoma; study focused on moyamoya after radiation | NR |
| Vaidya_2024 | 26 | 26 | 15 | median 9.5; range 1-34 | M 14; F 12 | Pediatric 19; Adult 7 | NF1 0; Sporadic 26; Unknown 0 | Sporadic optic chiasmatic-hypothalamic gliomas; 24 epicentered in hypothalamic-chiasmatic region and 2 in optic tract; thalamic/gangliocapsular extension 10, temporal lobe extension 5, frontal lobe extension 2, leptomeningeal dissemination 1 | visual symptoms 23; VA impairment 23; blindness 11 | any 26; biopsy 1 stereotactic biopsy; debulking/partial 25 | any 7; subgroup 7; regimen Chemotherapy regimens not specified | External-beam radiotherapy; technique not specified | NR | adjuvant 15; after surgery 15; indication Postoperative or primary chemoradiotherapy strategy according to age, extent and clinical presentation | median 30 mo; range 11-120 mo |
| DallaVia_2007 | 20 | 20 | 8 | median 2.42; mean 3.33; range 1.33-9.58 | M 8; F 12 | Pediatric 20; Adult 0 | NF1 20; Sporadic 0; Unknown 0 | NF1-associated OPG; Dodge 1 in 1, Dodge 2 in 7, Dodge 3 in 12; tumors involved optic nerve/chiasm/tract or hypothalamic pathway according to Dodge classification | visual symptoms 11; VA impairment 11; VF defect NR | any NR | any 12; subgroup 12; regimen Carboplatin and vincristine first line; other chemotherapy not specified | External conventional radiotherapy via fractionated guided stereotactic technique | total 54 Gy; 1.8 Gy/fraction; 30 fractions; range 54 Gy; median 54 Gy; mean 54 Gy | salvage 8; after chemotherapy 8; indication Progressive NF1-associated OPG in children older than 5 years after clinical or radiological deterioration | median 76 mo; mean 81 mo; range 5-216 mo |
| Awdeh_2012 | 20 | 20 | 20 | median 9.3; range 3.2-14.6 | M 9; F 11 | Pediatric 20; Adult 0 | NF1 0; Sporadic 20; Unknown 0 | All tumors in hypothalamus, optic chiasm or optic tracts; 3 limited to optic chiasm, 2 involved hypothalamus, chiasm and proximal optic nerves, remainder largely hypothalamic/chiasmatic | visual symptoms 20 evaluable VA/VF cohort; VA impairment 20 | any 17; biopsy 6; debulking/partial 11 | any 11; subgroup 11; regimen Carboplatin/vincristine and other prior chemotherapy regimens not fully specified | Conformal radiation therapy | total 54 Gy; 1.8 Gy/fraction; 30 fractions; range 54 Gy; median 54 Gy; mean 54 Gy | salvage 20; after surgery 17; after chemotherapy 11; indication CRT for pediatric OPG after progression or residual disease, with detailed serial visual follow-up | median 30 mo; range 8-62 mo |
| Green_2022 | NR | 88% OPG; exact count NR | 0 | NR | M NR; F NR | Pediatric NR; Adult NR | NF1 24% NF1; exact count NR; Sporadic NR; Unknown NR | Progressive pediatric low-grade glioma cohort, 88% optic pathway glioma; detailed tumor-location distribution not reported in abstract | visual symptoms NR; VA impairment NR; VF defect NR | NR | any NR; regimen Bevacizumab-based treatment, often after multiple prior lines | NA | NR | NR | NR |
| Morin_2024 | 182 | 182 | 75 | median 3.4; range 0.33-16.37 | M 86; F 96 | Pediatric 182; Adult 0 | NF1 65; Sporadic 117; Unknown 0 | OPG cohort: optic nerve only 18, chiasmatic involvement 111, retrochiasmatic involvement 53, hypothalamic involvement 56, second location at diagnosis 26, leptomeningeal dissemination 7 | visual symptoms 127 ophthalmologic symptoms/signs at diagnosis; VA impairment NR; VF defect NR | any 96; biopsy/resection details not fully extractable | any 103; subgroup 103; regimen Chemotherapy regimens varied across decades; targeted therapies in later era | Focal radiotherapy | range 50-60 in cerebrovascular-event subgroup Gy; median 50 Gy | first-line local/systemic combinations include RT; salvage NR; indication Part of local or combined treatment strategy in long-term OPG survivor cohort | median 206.4 mo; range 64.8-488.4 mo |
| Fernandes_2018 | 62 | 62 | 1 | median 4 | M NR; F NR | Pediatric 62; Adult 0 | NF1 62; Sporadic 0; Unknown 0 | NF1-associated optic pathway gliomas in children; detailed optic nerve, chiasm, hypothalamus or tract distribution not reported in abstract | visual symptoms NR; VA impairment NR; VF defect NR | NR | NR | Radiotherapy; technique not specified | NR | salvage 1; indication One patient required RT in NF1-OPG oncology cohort | mean 94.8 mo; range Up to 264 mo |
| Varan_2013 | 101 | 101 | 40 | median 6; range 0.08-18 | M 48; F 53 | Pediatric 101; Adult 0 | NF1 53; Sporadic 48; Unknown 0 | Optic glioma cohort including optic chiasm plus hypothalamus 32, intraorbital optic nerve 16, intraorbital optic nerve plus prechiasm 10, intraorbital optic nerve plus optic chiasm 10, optic chiasm plus hypothalamus plus optic tract 9, diffuse visual pathway glioma 8, intracranial optic nerve plus optic chiasm 6, intracranial optic nerve plus optic chiasm plus hypothalamus 4, prechiasm plus chiasm plus postchiasm 4, bilateral optic nerve 1, chiasm only 1 | visual symptoms 71; VA impairment 71; VF defect NR | any 41; biopsy 24; debulking/partial 13; GTR 4 | any 47; subgroup 47; regimen Chemotherapy regimens included vincristine/carboplatin and others; not fully specified | Conventional external-beam photon radiotherapy with Co60 or 6-MV photons | total 40.8-60 Gy; 1.7-2 Gy/fraction; range 40.8-60 Gy; mean 54 Gy | first-line NR; salvage NR; after surgery NR; after chemotherapy NR; indication RT used according to symptoms, age, progression and tumor location | median 96 mo |
| Nicolin_2009 | 133 | 133 | 16 | mean 5.89; range 0.34-16.8 | M 69; F 64 | Pediatric 133; Adult 0 | NF1 78; Sporadic 55; Unknown 0 | OPG locations: hypothalamic/chiasmatic 50, unilateral optic nerve 29, HC plus bilateral optic nerves 19, HC plus unilateral optic nerve 10, bilateral optic nerves 10, HC plus optic nerves plus optic radiations 9, HC plus thalamus 3, HC plus optic radiations 1, HC plus dissemination 2 | visual symptoms NR; VA impairment NR; VF defect NR | any 35; biopsy/resection details NR | any 47; subgroup 47; regimen Chemotherapy regimens not specified | Radiotherapy; technique not specified | NR | first-line 2; salvage 14; after surgery NR; after chemotherapy NR; indication RT as immediate or later treatment in selected OPG patients | median 103.2 mo; mean 108 mo; range 6.72-216 mo |
| Campagna_2010 | 32 | 32 | 17 | median 4.67; range 0.42-13 | M 13; F 19 | Pediatric 32; Adult 0 | NF1 0; Sporadic 32; Unknown 0 | Non-NF1 pediatric OPG cohort; Dodge I in 5 and Dodge III in 27; most tumors involved or extended to the hypothalamic-chiasmatic region | visual symptoms 32; VA impairment 32; VF defect NR | any 15; biopsy 7; debulking/partial 8 | any 21; subgroup 21; regimen Carboplatin and vincristine or other chemotherapy according to SIOP LGG strategy | External conventional radiotherapy | total 54 Gy; 1.8 Gy/fraction; 30 fractions; range 54 Gy; median 54 Gy | salvage 17; after surgery NR; after chemotherapy NR; indication RT for progressive or refractory non-NF1 OPG in older children or after chemotherapy | median 73 mo; range 6-168 mo |
| Shofty_2015 | 15 | 15 | 5 | mean 6.5; range 1-23 | M 8; F 7 | Pediatric 13; Adult 2 | NF1 10; Sporadic 5; Unknown 0 | Hypothalamic/chiasmatic OPG cohort; all patients Dodge II or III at diagnosis; tumors epicentered in hypothalamic/chiasmatic area | visual symptoms NR; VA impairment NR; VF defect NR | NR | any 15; subgroup 15; regimen Chemotherapy regimens not specified; chemotherapy-treated cohort | Radiotherapy; technique not specified | NR | salvage 5; after chemotherapy 5; indication RT for progression after chemotherapy in selected patients | mean 41 mo; range 12-96 mo |
| Bennebroek_2024 | 21 | 21 | 3 | median 7.2; range 1.9-17.9 | M 11; F 10 | Pediatric 21; Adult 0 | NF1 11; Sporadic 10; Unknown 0 | Isolated optic nerve glioma only; modified Dodge stage 1A in 12 ONGs and 1A plus 1C cisternal segment in 9 ONGs; one patient had bilateral ONG | visual symptoms 21 progressive isolated ONG with visual/radiological outcomes; VA impairment NR; VF defect NR | any 4; GTR 4 | any 14 SAT; subgroup 14; regimen Systemic anti-tumor treatment in 14; regimens not specified in extract | Radiotherapy; technique not specified | total 52.2 Gy; range 52.2-54.0 Gy; median 52.2 Gy | first-line 3; indication First-line treatment for progressive isolated optic nerve glioma in 3 patients | median 128.4 mo; range 27.6-220.8 mo |
| Regueiro_1995 | 35 | 35 | 35 | NR | M NR; F NR | Pediatric NR; Adult NR | NF1 NR; Sporadic NR; Unknown 35 | Optic pathway glioma cohort including 7 optic nerve tumors and 28 chiasmal tumors; RT-alone group included 6 optic nerve and 19 chiasmal tumors; subtotal surgery plus RT group included 1 optic nerve and 9 chiasmal tumors | NR | any 10; debulking/partial 10 | NR | Radiotherapy; technique not reported | NR | first-line 25; adjuvant 10; after surgery 10; indication Radiotherapy alone or postoperative radiotherapy after subtotal surgery | NR |
| Combs_2005 | 15 | 15 | 15 | median 6.9; range 0.67-33 | M 7; F 8 | Pediatric 13; Adult 2 | NF1 3; Sporadic 12; Unknown 0 | Optic pathway glioma cohort treated with FSRT; tumor confined to optic chiasm in 5, optic chiasm plus optic nerves in 3, optic nerve only in 3, and suprasellar/pituitary region in 4 | visual symptoms NR; VA impairment NR; VF defect NR | any 13; biopsy 5; debulking/partial 8 | NR | Fractionated stereotactic radiotherapy | total 52.2 Gy; 1.8 Gy/fraction; range 45.2-57.6 Gy; median 52.2 Gy | first-line 2; salvage 13; after surgery 13; indication FSRT for OPG, mostly after neurosurgical intervention; two had no surgery because diagnosis established radiologically | median 97 mo; range 8-151 mo |
| Rakotonjanahary_2015 | 180 | 180 | 55 subsequent RT after first-line chemotherapy | median 2.4; range NA | M NR; F NR | Pediatric 180; Adult 0 | NF1 60; Sporadic 120; Unknown 0 | Optic pathway glioma cohort; exact anatomic subgroup distribution not reported in extractable main text; included clinically/radiologically diagnosed OPG, including symptomatic/progressive tumors treated with chemotherapy | visual symptoms NR; VA impairment NR; VF defect NR | NR | any 180; subgroup 180; regimen Up-front BB-SFOP chemotherapy | NA | NR | salvage 55; after chemotherapy 55; indication Subsequent radiotherapy after first-line chemotherapy in subset of patients | median 163.2 mo; range 73.2-283.2 mo |
| Liu_2022 | 165 | 165 | 92 | NR | M 94; F 71 | Pediatric 165; Adult 0 | NF1 0; Sporadic 165; Unknown 0 | Sporadic pediatric OPG cohort after primary intratumor debulking; optic chiasm involved 162/165, hypothalamus 131/165, optic nerve only 3/165, optic tract 24/165, leptomeningeal metastases 17/165 | visual symptoms 133; VA impairment 133; VF defect NR | any 165; debulking/partial 165 | any 39 first adjuvant chemotherapy; subgroup 39; regimen Chemotherapy regimens not specified | Radiotherapy after surgery; technique not specified | NR | adjuvant 92; after surgery 92; indication First adjuvant treatment after primary intratumor debulking in sporadic pediatric OPG | median 39 mo; range 23-68 mo |
| Acharya_2019 | 41 | 41 | 41 | median 8; range 4.1-19.8 | M 24; F 17 | Pediatric 41; Adult 0 | NF1 0; Sporadic 41; Unknown 0 | Sporadic OPG treated with RT; posterior extent pre-chiasm 2, chiasm 1, post-chiasm/hypothalamus 38 | visual symptoms 41 all had serial visual acuity assessment; VA impairment 41; VF defect NR | any 22; biopsy 11; debulking/partial 11 | any 15; subgroup 15; regimen Chemotherapy before RT in 15; regimens not specified | 3D conformal photon therapy, intensity-modulated photon therapy, or intensity-modulated proton therapy | total 54 photon; 52.2 GyRBE proton Gy; 1.8 Gy/fraction; 30 photon; 29 proton fractions; range 52.2-54 Gy | first-line NR; salvage NR; after surgery 22; after chemotherapy 15; indication Definitive RT for sporadic OPG after radiographic progression or visual deterioration | mean 60 mo; range 2.9-147.6 mo |
| Zhou_2024 | 140 | 140 | 82 | mean 6.9; range 1-26 | M 70; F 70 | Pediatric 132; Adult 8 | NF1 NR; Sporadic NR; Unknown 140 | Pediatric OPG surgical cohort; Dodge I 2, Dodge II 29, Dodge III 109; anatomical type A 11, type M 31, type P 98; lateral extension 42; leptomeningeal dissemination 6 | visual symptoms 140 preoperative visual deterioration cohort; VA impairment 140; VF defect NR | any 140; debulking/partial 140 partial tumor resection | any 37 adjuvant chemotherapy; subgroup 37; regimen Chemotherapy regimens not specified | Radiotherapy; modality not specified | NR | adjuvant 82; after surgery 82; indication Adjuvant RT after initial partial tumor resection in patients selected by treatment strategy | mean 71 mo; range 12-137 mo |
| Quesada_2019 | 40 | 40 | 40 | median 8.2; range 1-19 | M NR; F NR | Pediatric mostly pediatric; exact count NR; Adult NR | NF1 3; Sporadic 37; Unknown 0 | OPG treated with RT; majority involved postchiasmatic optic tracts (90%); prechiasmatic location analyzed as risk factor for VA decline | visual symptoms 40 serial visual acuity outcomes after RT; VA impairment 40; VF defect NR | NR | NR | Radiation therapy; modality not specified | NR | indication Radiation therapy for optic pathway glioma with serial visual outcome assessment | median 37.2 mo |

Detailed patient, tumor, and treatment characteristics extracted from included studies. Variables include total study population, eligible OPHG population, radiotherapy/radiosurgery subgroup size, age, sex distribution, pediatric or adult status, NF1 status, tumor location, baseline visual impairment, prior surgery, prior chemotherapy, radiotherapy/radiosurgery modality, dose and fractionation, radiotherapy timing or indication, and follow-up duration. Values are reported as available from each study. Missing or unavailable data were recorded as NR.

**Table 3. Summary of visual outcome meta-analyses.**

| Outcome | Outcome_Type | Definition | Included_Studies_k | Visual_Outcome_Denominator_N | Event_Count_n | Crude_Proportion | Pooled_Proportion_percent | 95_percent_CI | Prediction_Interval | I2_percent | Tau2 | Cochran_Q_p_value | Interpretation |
| --- | --- | --- | --- | --- | --- | --- | --- | --- | --- | --- | --- | --- | --- |
| Visual preservation | Primary outcome | Stable or improved vision after radiotherapy or radiosurgery | 19 | 494 | 359 | 72.70% | 75.60% | 65.1%–83.7% | 31.5%–95.4% | 71.6 | 0.7654 | <0.0001 | Approximately three quarters of visual-outcome observations were preserved after RT/SRS; preservation was the dominant overall visual outcome. |
| Visual improvement | Secondary outcome | Improved vision after radiotherapy or radiosurgery | 19 | 494 | 125 | 25.30% | 24.70% | 19.5%–30.8% | 12.1%–43.9% | 36.3 | 0.1499 | 0.0583 | Approximately one quarter of visual-outcome observations improved, indicating that visual recovery occurred but was less frequent than stabilization. |
| Visual stability | Secondary outcome | Stable vision after radiotherapy or radiosurgery | 19 | 494 | 234 | 47.40% | 46.70% | 37.0%–56.6% | 16.5%–79.5% | 64.1 | 0.4668 | <0.0001 | Visual stability represented the largest component of visual preservation, supporting stabilization as the main visual benefit of RT/SRS. |
| Visual worsening | Secondary outcome | Worsened or deteriorated vision after radiotherapy or radiosurgery | 19 | 494 | 120 | 24.30% | 20.60% | 12.7%–31.6% | 2.8%–70.0% | 69.1 | 1.022 | <0.0001 | Approximately one fifth of visual-outcome observations worsened, showing that RT/SRS did not eliminate the risk of further visual decline. |

Summary of pooled visual outcome estimates after radiotherapy or radiosurgery. The primary outcome was visual preservation, defined as stable or improved vision. Secondary outcomes included visual improvement, visual stability, and visual worsening. For each outcome, the table reports the number of included studies, total visual-outcome denominator, event count, crude proportion, pooled proportion, 95% confidence interval, prediction interval, I², tau², Cochran’s Q p-value, and interpretation.

**Table 4. Key subgroup and sensitivity analyses.**

| Analysis_Category | Outcome | Subgroup_or_Sensitivity | Pooled_Proportion_percent | Subgroup_p_value_or_Result | Interpretation |
| --- | --- | --- | --- | --- | --- |
| Radiation technique | Visual preservation | Historical conventional photon RT | 71% | p = 0.7269 | Visual preservation was similar across radiation-technique groups; no statistically significant subgroup difference was observed. |
| Radiation technique | Visual preservation | SRS/Gamma Knife | 83% | p = 0.7269 | Favorable preservation was observed in SRS/Gamma Knife cohorts, but this subgroup was small and exploratory. |
| Radiation technique | Visual preservation | Mixed or unclear RT approach | 76% | p = 0.7269 | Mixed or unclear RT cohorts showed preservation similar to the overall estimate. |
| Radiation technique | Visual preservation | Fractionated stereotactic RT | 66% | p = 0.7269 | Fractionated stereotactic RT cohorts showed numerically lower preservation, but the subgroup difference was not significant. |
| Radiation technique | Visual preservation | 3D conformal photon RT | 75% | p = 0.7269 | Only one study contributed to this subgroup, so no reliable modality-specific inference can be made. |
| Radiotherapy timing | Visual preservation | Salvage RT | 77% | p = 0.0580 | Salvage RT showed visual preservation close to the overall estimate. |
| Radiotherapy timing | Visual preservation | Primary or upfront RT | 63% | p = 0.0580 | Primary/upfront RT showed numerically lower preservation, but this should be interpreted cautiously because of confounding by indication and treatment-era effects. |
| Radiotherapy timing | Visual preservation | Mixed or unclear timing | 89% | p = 0.0580 | Mixed or unclear timing cohorts showed high preservation, but interpretation is limited by heterogeneity. |
| Radiotherapy timing | Visual preservation | Adjuvant or post-surgical RT | 75%–77% | p = 0.0580 | Adjuvant/post-surgical timing categories were based on few studies and were considered exploratory. |
| Study purity | Visual preservation | Pure RT/SRS cohorts | 85% | p = 0.0104 | Pure RT/SRS cohorts showed higher and more consistent visual preservation than RT-containing mixed cohorts. |
| Study purity | Visual preservation | RT-containing mixed cohorts | 66% | p = 0.0104 | Mixed-treatment cohorts showed lower preservation, likely reflecting methodological heterogeneity and difficulty isolating the radiation effect. |
| Risk of bias | Visual preservation | Low ROB studies | 65% | p = 0.1284 | Visual preservation did not differ significantly by risk-of-bias category. |
| Risk of bias | Visual preservation | Moderate ROB studies | 79% | p = 0.1284 | Moderate-risk studies showed numerically higher preservation than low-risk studies. |
| Risk of bias | Visual preservation | High ROB studies | 83% | p = 0.1284 | High-risk studies showed numerically high preservation, but this may reflect small-study and reporting limitations. |
| Sensitivity analysis | Visual preservation | Excluding high-risk-of-bias studies | 73% | Robust | Excluding high-risk studies did not materially change the pooled preservation estimate. |
| Sensitivity analysis | Visual preservation | Full-text studies only | 75% | Robust | Restricting to full-text studies produced an estimate nearly identical to the main analysis. |
| Sensitivity analysis | Visual preservation | Pure RT/SRS cohorts only | 85% | Robust | Pure radiation-based cohorts showed high preservation with no observed heterogeneity. |
| Sensitivity analysis | Visual preservation | Leave-one-out analysis | 74%–77% | No single influential study | The pooled visual preservation estimate remained stable after sequential exclusion of individual studies. |
| Radiation technique | Visual improvement | SRS/Gamma Knife | 19% | p = 0.0446 | Technique subgroup differences were observed for visual improvement, but some groups were represented by few studies. |
| Radiation technique | Visual improvement | 3D conformal photon RT | 50% | p = 0.0446 | This subgroup was represented by a single study and should not be interpreted as evidence of superiority. |
| Radiotherapy timing | Visual improvement | Salvage RT | ~25% | p = 0.2418 | Visual improvement did not differ significantly by RT timing. |
| Radiotherapy timing | Visual improvement | Primary or upfront RT | ~25% | p = 0.2418 | Primary/upfront and salvage RT showed similar improvement estimates. |
| Study purity | Visual improvement | Pure RT/SRS cohorts | 27% | p = 0.2723 | Pure RT/SRS cohorts showed visual improvement in approximately one quarter of observations. |
| Study purity | Visual improvement | RT-containing mixed cohorts | 23% | p = 0.2723 | Study purity did not significantly modify visual improvement. |
| Risk of bias | Visual improvement | Low ROB studies | 24% | p = 0.4664 | Visual improvement did not differ significantly by risk-of-bias category. |
| Risk of bias | Visual improvement | Moderate ROB studies | 28% | p = 0.4664 | Moderate-risk studies showed improvement similar to the overall estimate. |
| Risk of bias | Visual improvement | High ROB studies | 17% | p = 0.4664 | High-risk studies showed numerically lower improvement. |
| Sensitivity analysis | Visual improvement | Excluding high-risk-of-bias studies | 26% | Robust | Exclusion of high-risk studies did not materially alter visual improvement. |
| Sensitivity analysis | Visual improvement | Full-text studies only | 24% | Robust | Restriction to full-text studies produced a similar improvement estimate. |
| Sensitivity analysis | Visual improvement | Leave-one-out analysis | 24%–26% | No single influential study | The pooled visual improvement estimate remained stable after sequential exclusion of individual studies. |
| Radiation technique | Visual stability | SRS/Gamma Knife | 65% | p = 0.0265 | SRS/Gamma Knife cohorts had the highest visual stability estimate, but interpretation is exploratory. |
| Radiotherapy timing | Visual stability | Salvage RT | 48% | p = 0.0041 | Salvage RT showed visual stability close to the overall estimate. |
| Radiotherapy timing | Visual stability | Primary or upfront RT | 36% | p = 0.0041 | Primary/upfront RT showed lower stability, but confounding by indication is likely. |
| Radiotherapy timing | Visual stability | Mixed or unclear timing | 55% | p = 0.0041 | Mixed/unclear timing cohorts showed the highest stability among timing groups. |
| Study purity | Visual stability | Pure RT/SRS cohorts | 57% | p = 0.1572 | Pure RT/SRS cohorts showed numerically higher stability than mixed-treatment cohorts. |
| Study purity | Visual stability | RT-containing mixed cohorts | 40% | p = 0.1572 | Study purity did not significantly modify visual stability. |
| Risk of bias | Visual stability | Low ROB studies | 37% | p = 0.0126 | Visual stability differed significantly by risk-of-bias category. |
| Risk of bias | Visual stability | Moderate ROB studies | 46% | p = 0.0126 | Moderate-risk studies showed stability close to the overall estimate. |
| Risk of bias | Visual stability | High ROB studies | 64% | p = 0.0126 | High-risk studies showed the highest visual stability, potentially reflecting reporting limitations. |
| Sensitivity analysis | Visual stability | Excluding high-risk-of-bias studies | 42% | Robust | Excluding high-risk studies reduced the stability estimate but did not change the overall interpretation. |
| Sensitivity analysis | Visual stability | Full-text studies only | 46% | Robust | Full-text-only analysis was similar to the main stability estimate. |
| Sensitivity analysis | Visual stability | Leave-one-out analysis | 45%–49% | No single influential study | The pooled stability estimate remained stable after sequential exclusion of individual studies. |
| Radiation technique | Visual worsening | SRS/Gamma Knife | 17% | p = 0.5553 | Worsening did not differ significantly by radiation technique. |
| Radiation technique | Visual worsening | Historical conventional photon RT | 29% | p = 0.5553 | Historical conventional photon RT showed numerically higher worsening, but the subgroup difference was not significant. |
| Radiotherapy timing | Visual worsening | Primary or upfront RT | 35% | p = 0.0498 | Primary/upfront RT showed the highest worsening estimate, but this may reflect confounding by indication, older treatment eras, or worse baseline disease. |
| Radiotherapy timing | Visual worsening | Salvage RT | 14% | p = 0.0498 | Salvage RT showed lower worsening than upfront RT in subgroup analysis. |
| Study purity | Visual worsening | Pure RT/SRS cohorts | 15% | p = 0.2837 | Pure RT/SRS cohorts showed lower worsening with no observed heterogeneity. |
| Study purity | Visual worsening | RT-containing mixed cohorts | 26% | p = 0.2837 | Mixed-treatment cohorts showed numerically higher worsening, but the subgroup difference was not significant. |
| Risk of bias | Visual worsening | Low ROB studies | 34% | p = 0.0826 | Worsening was numerically highest in low-risk studies. |
| Risk of bias | Visual worsening | Moderate ROB studies | 14% | p = 0.0826 | Moderate-risk studies showed lower worsening. |
| Risk of bias | Visual worsening | High ROB studies | 17% | p = 0.0826 | High-risk studies showed lower worsening than low-risk studies, possibly reflecting reporting differences. |
| Sensitivity analysis | Visual worsening | Excluding high-risk-of-bias studies | 22% | Robust | Exclusion of high-risk studies did not materially change the worsening estimate. |
| Sensitivity analysis | Visual worsening | Full-text studies only | 21% | Robust | Full-text-only analysis was similar to the main worsening estimate. |
| Sensitivity analysis | Visual worsening | Leave-one-out analysis | 19%–23% | No single influential study | The pooled worsening estimate remained stable after sequential exclusion of individual studies. |

Summary of key subgroup and sensitivity analyses for visual outcomes after radiotherapy or radiosurgery. Subgroup analyses included radiation technique, stereotactic radiosurgery/Gamma Knife cohorts, radiotherapy timing, study purity, and risk-of-bias category. Sensitivity analyses included exclusion of high-risk-of-bias studies, restriction to full-text studies, restriction to pure RT/SRS cohorts, and leave-one-out analysis. For each analysis, the table reports the outcome, subgroup or sensitivity analysis, pooled proportion, subgroup p-value or sensitivity result, and interpretation.
